# Persistent Pain in Wales: Prevalence and Healthcare Utilisation from a Population-Scale Retrospective Cohort Study

**DOI:** 10.1101/2025.06.28.25330404

**Authors:** Helen Daniels, Tim Osborne, Athena McBride, Owen Hughes, Natalie Joseph-Williams, Adrian Edwards, Ashley Akbari, Rowena Bailey, Rhiannon K Owen

## Abstract

Persistent pain has a significant impact on quality of life and places considerable demand on the NHS (National Health Service). In 2023, Welsh Government published their guidance ‘Living with Persistent Pain’, marking persistent pain as a national priority. The research aimed to provide evidence to help inform NHS leads in Wales around current and future services, capacity planning, and to aid in implementing health policies.

We conducted a retrospective cohort study covering the period 2010-2023. Anonymised, individual-level, population-scale linked routinely-collected electronic health records and administrative data from the SAIL Databank was used. The population included people living in Wales and registered with a General Practice (GP) that provides data to SAIL. Three cohorts of people were created. These were i) people with diagnostic codes relevant to persistent pain, ii) people prescribed opioids or gabapentinoids for 3 months or more, and iii) people referred to specialist outpatient pain services. A comparator group included people who did not meet any of these criteria. We measured the proportion of the population living with persistent pain, demographic details, and health status. Healthcare use data included GP appointments and prescriptions, hospital admissions, emergency department visits, and outpatient appointments.

The results showed that 15% of the population of Wales are living with persistent pain. Persistent pain was more common among older adults, women, and individuals living in more deprived areas. A higher burden of frailty and comorbid conditions was observed in the persistent pain cohort compared to the general population. People living with persistent pain had 63% more GP events than those without. Those referred to pain services had more healthcare interactions overall and were younger and less frail compared to those not referred. Trends over time showed small but statistically significant decline was observed in the prevalence of persistent pain over the study period. After adjusting for demographics and seasonal changes—there were small but significant monthly increases in GP events, hospital admissions, emergency attendances, for the persistent pain cohort. Outpatient attendances showed small but statistically significant month-on-month decreases.

Findings may point to unmet need in accessing specialist care, particularly among older adults and individuals from more deprived areas. Patterns of pain prevalence and service use reflect existing inequalities across age, sex, and socioeconomic status. There is a need for further research, service improvements and policy development.

**Funding statement:** The authors and their Institutions were funded for this work by the Health and Care Research Wales Evidence Centre, itself funded by Health and Care Research Wales on behalf of Welsh Government.

## 1 Introduction

### 1.1 Background and rationale

Persistent pain is a major public health concern, and as many as half of the population of Wales are thought to be affected (1). Persistent pain not only limits physical function, but there are often significant psychological and social consequences associated with the condition (1). Persistent pain is defined as pain that persists or recurs for more than three months (2). The shift from the term ‘chronic pain’ arose from an appetite to avoid potential stigma, to aid in management, and to help improve others’ understanding of the condition (3). Multiple demographic and socioeconomic factors— such as deprivation, unemployment, and ageing are associated with an increased risk of persistent pain. However, significant research gaps remain, particularly around the effectiveness of interventions and the cultural and demographic factors that influence access to services, which are crucial to improving prevention and treatment (4).

In 2023, the Welsh Government published its guidance ‘Living with Persistent Pain’, marking it as a clear national priority (3). Eluned Morgan, the Minister for Health and Social Care, explained that it is:

> Imperative for the NHS and social care to focus on making the best use of resources to make a real difference. Continuing concentration on prudent healthcare principles and value-based healthcare is essential to allow the development and delivery of efficient and effective services (3).Given the extent of the issue, and not least due to the current financial climate, the Welsh Government is keen to address the evidence gap regarding the management of individuals experiencing persistent pain (3). This report is a request from NHS Practice Leads for the Health and Care Research Wales Evidence Centre to provide evidence to inform their decision-making around current and future services, capacity planning and to aid in implementing health policies—all of which will expand on their 2023 guidance.

### 1.2 Objectives

Our objectives for this report were as follows:

1. Develop a methodology to identify all individuals resident in Wales experiencing persistent pain using routinely-collected electronic health record (EHR) data sources.
2. Estimate the size of the prevalent population of individuals living with persistent pain.
3. Compare their demographic, socioeconomic and socio-geographic characteristics with the comparator group (all individuals not identified as experiencing persistent pain) on the earliest date of their cohort entry:

a. Compare differences (characteristics, health resource utilisation (HRU)) between those with persistent pain who access persistent pain outpatient services and those who do not.
4. Quantify and describe the HRU patterns for people with persistent pain and those referred to pain services and compare them with the comparator group (all individuals not identified as experiencing persistent pain).

## 2 Methods

### 2.1 Study design

This is a retrospective cohort study using anonymised, individual-level, population-scale linked routinely-collected EHR and administrative data sources.

2.2 Data sources and management

Our study used data held within the SAIL Databank (5), the national trusted research environment (TRE) for Wales. SAIL contains anonymised, population-scale, individual-level, routinely-collected administrative and EHR data sources for the population of Wales and for those accessing services in Wales. These are collected from interactions with the National Health Service (NHS) (for all services provided in Wales and all services received by Welsh residents occurring across the UK) since 1998 (approximately 9 million individuals, 3.2 million active residents) (6). The SAIL Databank currently receives complete population coverage data from secondary care (emergency department, hospital admissions and outpatient appointments) and data from 86% of general practices (7). A unique individual anonymised person identifier, Anonymised Linking Field (ALF), facilitates consistent linkage across data sources at the individual-level. A unique property reference number (UPRN) (which is encrypted to a Residential Anonymous Linking Field (RALF)) enables linkage of household variables, along with Lower-layer Super Output Area (LSOA) of residence for area-level linkage (8).

The Welsh Demographic Service Dataset (WDSD) is one of the many individual-level linkable population-scale anonymised data sources held within the SAIL Databank. It provides demographic characteristics (e.g. week of birth, sex) and residence history of people registered with general practices in Wales. The Welsh Longitudinal General Practice (WLGP) data contains records relating to activity in primary care services, including individuals’ symptoms, investigations, diagnoses and prescribed medication. We used this data source to identify individuals with persistent pain through their prescriptions and diagnoses. Secondary care data within the SAIL Databank include the Patient Episode Database for Wales (PEDW), which holds records of hospital admissions and associated administrative, diagnostic and operative information, and the Emergency Department Data Set (EDDS), which records administrative and clinical information relating to emergency department attendances. The Outpatient Database for Wales (OPDW) and the Outpatient Referrals Dataset (OPRD) contains data on outpatient attendance and referrals.

### 2.3 Participants

This study included individuals living in Wales registered to a SAIL-providing GP during the study period from 1^st^ January 2010 to 31^st^ December 2023. We created three cohorts of individuals experiencing persistent pain using three definitions based on i) diagnoses, ii) prescriptions, and iii) referrals to pain services. Individuals could be included in one or more of these cohorts if they met the inclusion criteria.

#### 2.3.1 Diagnoses

In collaboration with project team members and stakeholders, we compiled a list of diagnostic Read version 2 codes that were likely to refer to persistent pain (see Appendix 1), such as chronic primary headaches, chronic postsurgical pain, and chronic posttraumatic pain. In addition, we converted the International Classification of Diseases (ICD) 11^th^ revision chronic pain codes as described in an academic paper by (9) into their 10^th^ revision equivalents (ICD-10), since secondary care data sources within SAIL use ICD-10 codes for diagnoses. Project team members and stakeholders reviewed these codes before finalising them (Appendix 2). If an individual had one or more of these Read or ICD-10 codes recorded in their primary or secondary care EHR (data sources: WLGP, PEDW, EDDS, OPDW), were living in Wales and were registered with a SAIL-providing general practice at that time, then those individuals entered the cohort on the earliest date these conditions were met. These individuals remained in the cohort until the study period ended or until death, whichever occurred first.

### 2.3.2 Pain services

We identified individuals referred to specialist outpatient pain services using the 191 referral code in the OPDW and OPRD data sources. If individuals were living in Wales and were registered with a SAIL- providing general practice at the time of the referral, then individuals entered this cohort on their earliest referral date. These individuals remained in the cohort until the study period ended or until death, whichever occurred first.

#### 2.3.3 Prescriptions

In collaboration with project team members and stakeholders, we compiled a list of medications commonly prescribed to individuals experiencing persistent pain. These included opioids and gabapentinoids (Appendix 3). Individuals entered this cohort if they had an active prescription period for opioids and/or gabapentinoids for three months or more during the study period, were living in Wales and registered with a SAIL providing general practice. We defined an active prescription period as a period where the individual received a repeat prescription for pain medication within eight weeks of the previous prescription. If the repeat prescription occurred more than eight weeks after the previous prescription, then the active prescription period ended 28 days after the previous prescription date. The subsequent prescription date became the new index date for the next period. Individuals only remained in this cohort for the duration of their active prescription period but were able to re-enter the cohort at subsequent active prescription periods.

#### 2.3.4 Comparator cohort

Any individuals not meeting the criteria set out in Sections 2.3.1, 2.3.2 or 2.3.3 entered a comparator cohort. Their cohort entry date was the earliest date during the study period where the individual was living in Wales and was registered with a SAIL-providing general practice. Individuals remained in this cohort until they met one or more of the criteria for persistent pain (as described in Sections 2.31, 2.3.2 or 2.3.3), migrated out of Wales, died, or the end of the study period was reached, whichever occurred first. Individuals could re-enter the comparator cohort if they no longer met the criteria for persistent pain at a later date and/or they migrated back to Wales.

### 2.4 Variables

#### 2.4.1 Exposures

We extracted sex (male or female), week of birth—used to calculate the age in years of individuals on their earliest cohort entry dates—and Lower-layer Super Output Areas (LSOAs) version 2011 from the WDSD at cohort entry. Wales is divided into 1,909 LSOAs containing an average of 1,600 people (10). We used LSOAs to identify whether individuals lived in rural or urban areas using the Office for National Statistics (ONS) rural-urban classification (11). In addition, we used LSOA codes to map to the Welsh Index of Multiple Deprivation (WIMD) version 2019 (12). The overall WIMD scores were calculated, ranked and divided into fifths to create a quintile from 1 to 5, where 1 represents the ‘most deprived’, and 5 the ‘least deprived’. We recorded the WIMD quintile for individuals at cohort entry. We also used LSOA codes to assign one of the seven Welsh Health Boards to each individual (13).

The electronic Frailty Index (eFI) assigns a frailty score to an individual using 36 variables from primary care data, including falls, symptoms, signs, diseases, disabilities, polypharmacy, and abnormal laboratory values, referred to as deficits. Baseline eFI was calculated using WLGP data using a 10-year look back window from the earliest data of an individual’s cohort entry. The eFI scores were categorised as: fit (eFI value of 0–0.12), mild (>0.12–0.24), moderate (>0.24–0.36) or severe frailty (>0.36), or missing if there were no data available to calculate a score (14).

The Charlson Comorbidity Index is a weighted score used in medical and research practice to predict mortality risk and healthcare outcomes based on an individual’s comorbid conditions. Scores of 0-1 indicate a low risk of mortality and/or morbidity, 2-3 indicate moderate risk, and scores of 4 or more indicate a high risk (15). We calculated the Charlson Comorbidity Index for each individual at cohort entry and calculated the median score and IQR for each cohort. This is a modified version of the Charlson Index; the original includes reference to human immunodeficiency virus (HIV), however, SAIL does not have access to these data due to current Welsh legislation around the use of sensitive data items.

#### 2.4.2 Outcomes

Our main outcome measures were the counts of all individuals experiencing persistent pain and the proportion of the total population that these individuals represent. We also calculated the counts and proportions separately for each of the three persistent pain cohorts, using each of the three criteria set out in Sections 2.31, 2.32 and 2.33. Furthermore, we assessed the health utilisation of each persistent pain cohort per month using counts of the number of patient interactions with healthcare services, including:

- Counts of primary care events (appointments and prescriptions) using WLGP records. Events recorded on the same day were counted as a single event.
- Counts of secondary care hospital admissions and secondary care emergency admissions (defined using admission method codes 1-28 (16)). These were derived from PEDW using

person spell identifiers, which link together multiple episodes of care across hospital spells and providers to count a continuous admission as a single event.

- Counts of emergency department visits, derived from EDDS. Visits recorded on the same attendance date were counted as a single event.
- Counts of outpatient appointments, derived from OPDW.

### 2.5 Statistical Analysis

We produced a cohort profile for the persistent pain and comparator cohorts by calculating descriptive summary statistics for the included population, summarising key demographic, socioeconomic, and socio-geographic characteristics. Categorical variables were summarised using counts and percentages, with the denominator based on the total individuals in the respective cohort. Continuous variables were assessed for normality and summarised using means and standard deviations where normality held, or medians and interquartile ranges where the distributions were skewed. Chi-squared tests were used to examine associations between categorical variables, both within a single group and between independent groups. T-tests were used to assess differences in means between two independent groups when data were normally distributed, while Wilcoxon rank-sum tests (non-parametric alternatives) were used when normality assumptions were not met.

Trends in persistent pain were expressed as the rate per 100,000 person-years and were assessed using negative binomial regression models using a log link function with an offset for the population at risk. Trends in healthcare utilisation were expressed per 1,000 person-years. Predicted healthcare utilisation rates were calculated to compare observed versus expected counts. Negative binomial regression unadjusted models, using an offset for population at risk, were fitted to monthly aggregated data to predict expected healthcare utilisation. Trends were modelled using epidemiological month and seasonal effects were captured using Fourier terms. Outputs were visualised by plotting the observed vs. expected counts each month, together with the 95% prediction intervals obtained from the regression model. Decomposition plots from this analysis for each of the healthcare utilisation measures were produced.

Models were adjusted for potential confounders, including WIMD, age, and sex, and included an offset term for the population at risk. Time was modelled using epidemiological month and seasonal effects were captured using Fourier terms. Analyses were first conducted across all pain cohorts combined to assess overall trends in event rates over time, with rate ratios (RR) and 5% significance levels reported. Further comparisons were made between the combined pain and comparator groups, and between individuals who accessed pain services and those who did not, to determine percentage differences in event rates.

## 3 Results

### 3.1 Overview

We analysed 37.6 million patient-years of data spanning from 1^st^ January 2010 to 31^st^ December 2023. 4,216,980 people were registered with a SAIL-providing GP during our study period. Of these, 3,984,768 individuals were living in Wales and were included in our study. 603,697 were identified as living with persistent pain, and 3,381,071 were in the general population comparator cohort who did not meet any of our inclusion criteria for persistent pain. This means that approximately 15%, or 15,150 persons per 100,000, for the entire study period, met the criteria for persistent pain during cohort follow up. Figure 1 illustrates the flow of participants^1^ through the study selection process, including the formation of the comparator cohort and the various pain cohorts.

**Figure 1.**
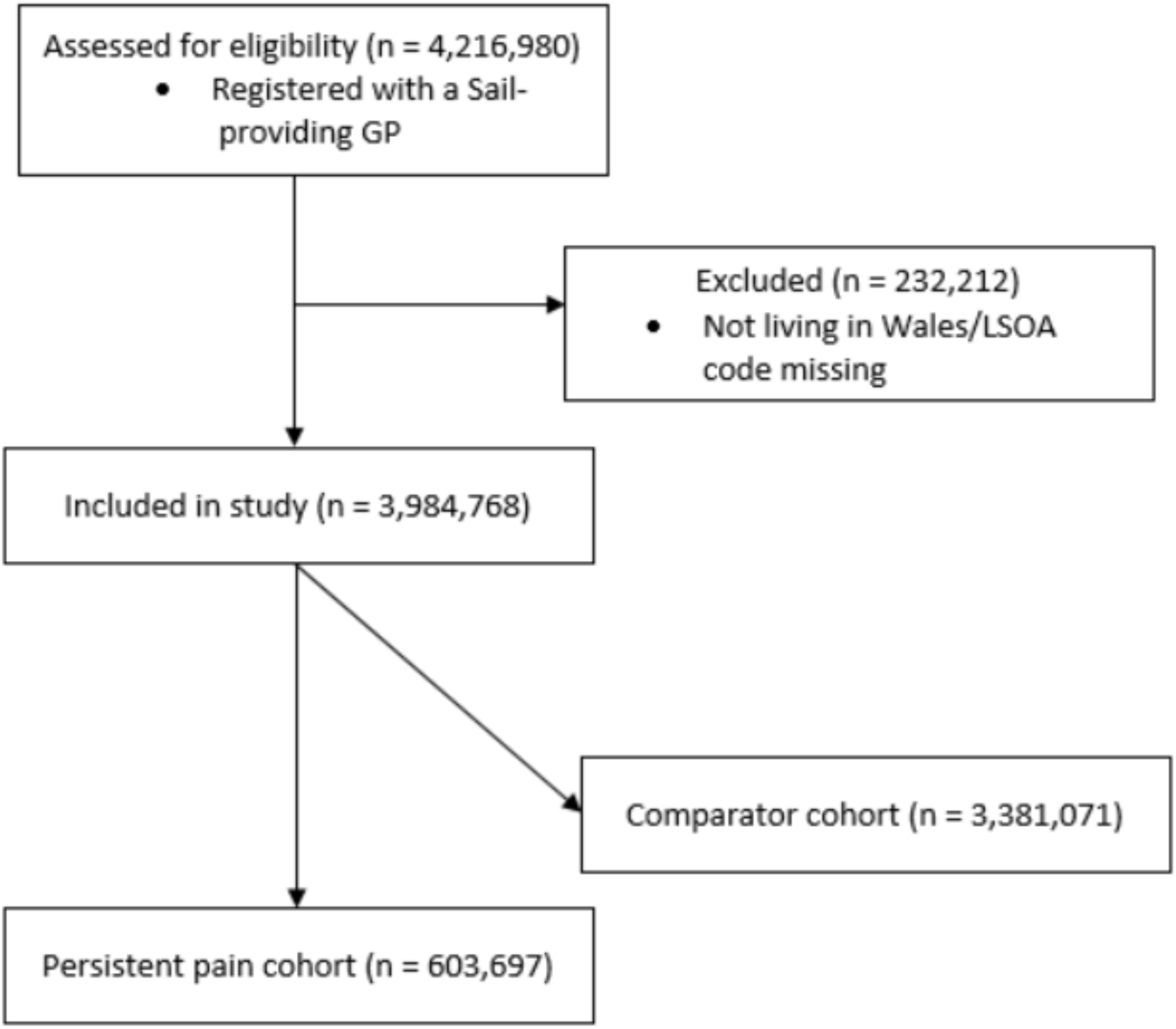
Study flow diagram.

The largest pain cohort consisted of 403,705 individuals with a diagnostic code indicating persistent pain in their EHR, representing 10% of the general population (10,131 persons per 100,000) for the entire study period. Individuals in receipt of a prescription for pain medication for three months or more accounted for 291,350 individuals, 7.3% or 7,312 individuals per 100,000 of the general population. The smallest pain cohort, those referred to specialist outpatient services, included 73,782 individuals, representing 1.85% of the general population (1,851 persons per 100,000) (see Table 1 for all yearly counts). Figure 2 shows the Venn diagram of the distribution of individuals between all three pain cohorts for January 2010. See Appendix 4 for the graphical representation of Table 1. Table 2 gives the distribution of each cohort across Wales’ seven healthboards.

**Figure 2.**
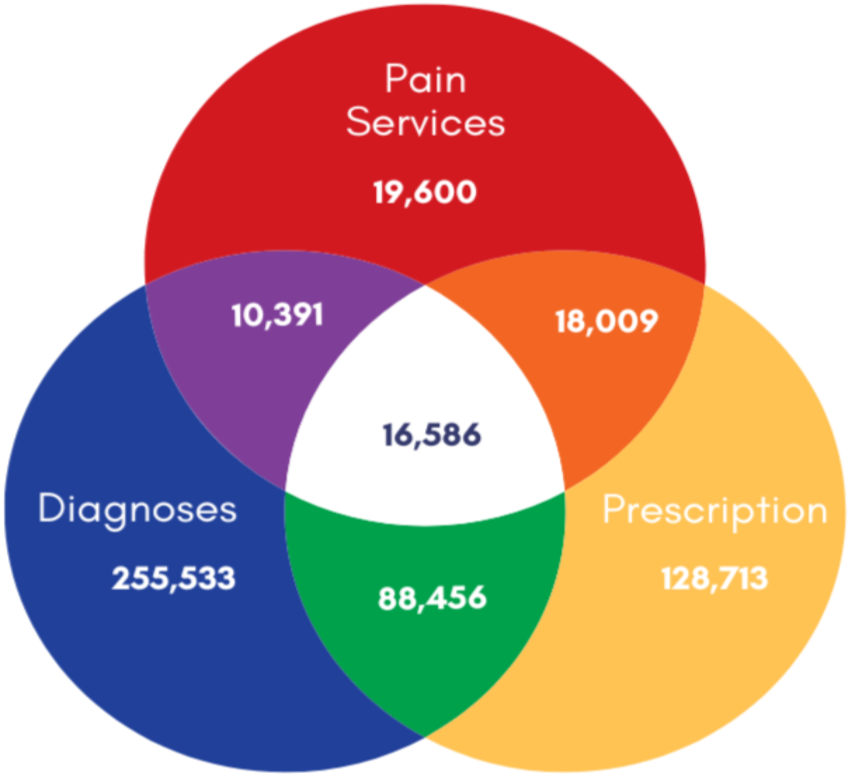
Venn diagram showing the overlap between the three pain cohorts for January 2010.

**Table 1.**
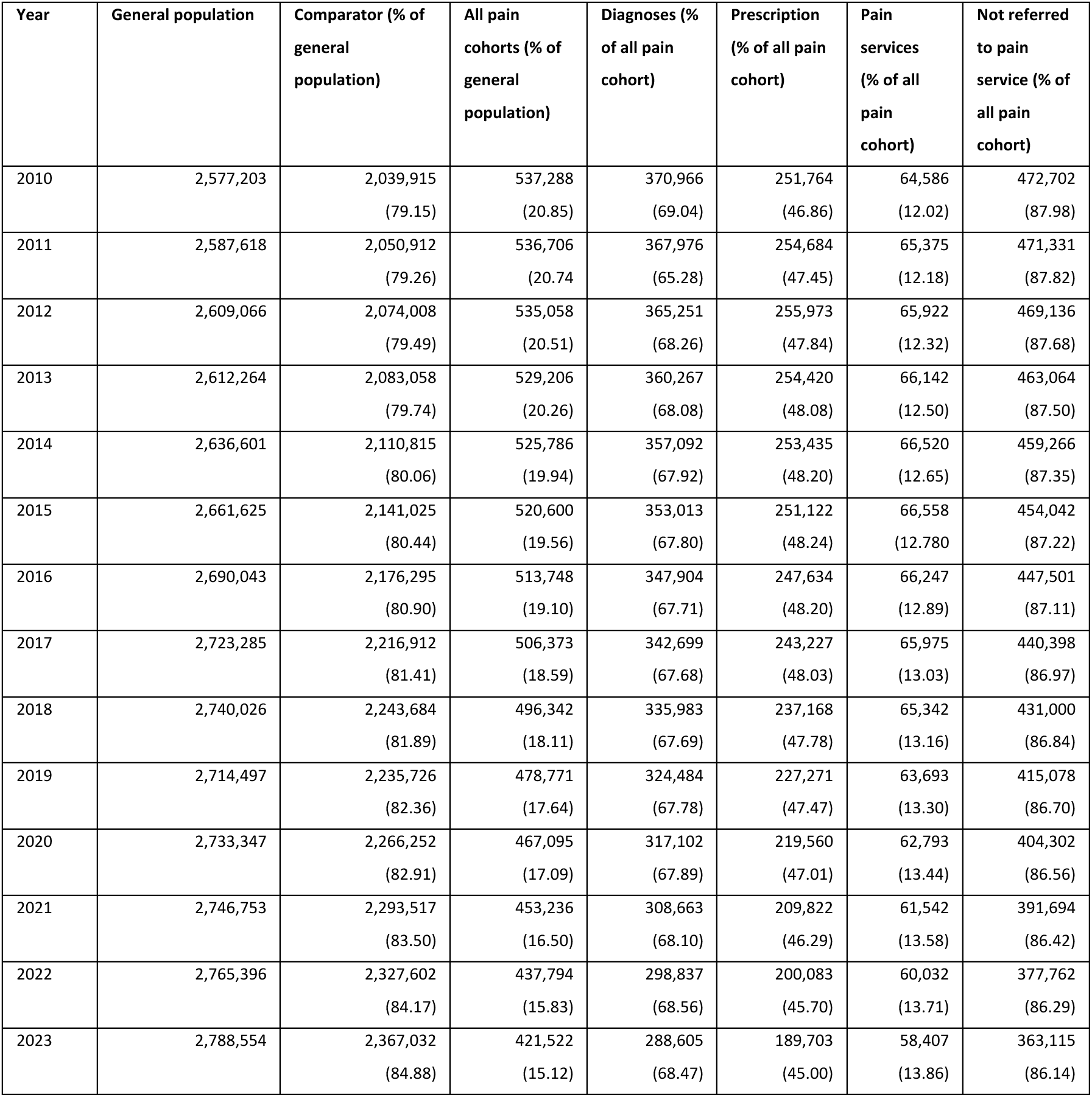
Yearly counts taken on 1st January of every study year for each cohort.

**Table 2.** Overall cohort counts by health board.

| Health Board | No pain (% of cohort) | All pain (% of cohort) | Diagnoses (% of cohort) | Prescription (% of cohort) | Service (% of cohort) | Non-pain service (% of cohort) |
| --- | --- | --- | --- | --- | --- | --- |
| Aneurin Bevan University | 571,845<br>(16.91) | 115,776<br>(19.18) | 74,477<br>(18.45) | 59,630<br>(20.47) | 14,713<br>(19.94) | 101,063<br>(19.07) |
| Betsi Cadwaladr University | 717,694<br>(21.23) | 134,731<br>(22.32) | 90,428<br>(22.40) | 61,503<br>(21.11) | 21,767<br>(29.50) | 112,964<br>(21.32) |
| Cardiff and Vale University | 681,727<br>(20.16) | 79,424<br>(13.16) | 51,822<br>(12.84) | 39,629<br>(13.60) | 7,390<br>(10.02) | 72,034<br>(13.59) |
| Cwm Taf Morgannwg University | 485,611<br>(14.36) | 92,824<br>(15.38) | 59,645<br>(14.77) | 50,272<br>(17.25) | 5,563<br>(7.54) | 87,261<br>(16.47) |
| Hywel Dda University | 401,702<br>(11.88) | 73,507<br>(12.18) | 49,761<br>(12.33) | 33,727<br>(11.58) | 10,268<br>(13.92) | 63,239<br>(11.93) |
| Powys Teaching | 69,200<br>(2.05) | 13,564<br>(2.25) | 9,379<br>(2.32) | 5,882<br>(2.02) | 1,726<br>(2.34) | 11,838<br>(2.23) |
| Swansea Bay University | 449,530<br>(13.30) | 93,698<br>(15.52) | 68,105<br>(16.87) | 40,617<br>(13.94) | 12,321<br>(16.70) | 81,377<br>(15.36) |
| Missing | 3,762<br>(0.11) | 173<br>(0.03) | 86<br>(0.02) | 90<br>(0.03) | 34<br>(0.05) | 139 (0.03) |

All pain cohorts are all individuals who meet one of the three criterion (diagnoses, prescription, pain services) for persistent pain. Individuals can meet one, two, or all criteria and therefore can be a member of more than one cohort concurrently. Individuals not referred to pain service are all those who meet the criteria for diagnoses and/or prescriptions but who have not beon referred to specialist outpatient pain services. Percentages are the percentage of the all pain cohorts column, i.e. all individuals who have experienced persistent pain.

Figure 3 shows the rate of individuals experiencing persistent pain per 100,000 of the general population from 2010 to 2024. The negative binomial regression models indicate a small, but statistically significant decrease in the number of individuals living with persistent pain, with a percentage decrease ranging from 0.003 to 0.008 (p < 0.0001) (see coefficients for this model in Appendix 5).

**Figure 3.**
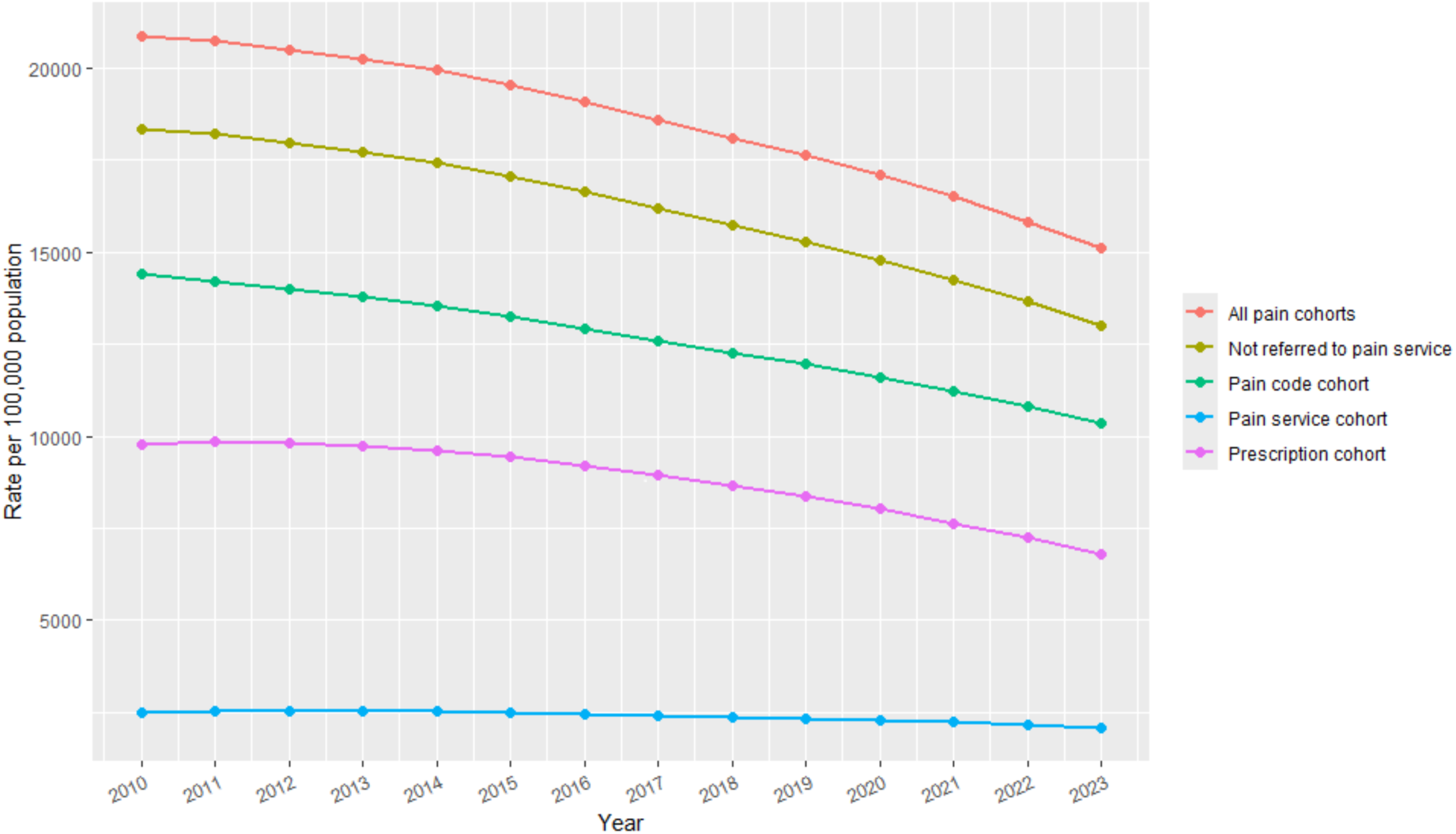
Plot of pain cohorts per Figure 3 100,000 person years, 2010-2023.

### 3.2 Demographics

Table 3 shows the distribution of deprivation acorss each of the seven health boards. Table 4 gives the demographic information for each of the cohorts in this study.

| Health Board | WIMD Quintile | Non-pain (% of cohort) | All pain (% of cohort) | Diagnoses (% of cohort) | Prescription (% of cohort) | Service (% of cohort) | Non service (% of cohort) |
| --- | --- | --- | --- | --- | --- | --- | --- |
| Aneurin Bevan University | 1 | 175,867 (5.2) | 34,201 (5.67) | 20,061 (4.97) | 19,718 (6.77) | 4,906 (6.65) | 34,201 (5.53) |
| Aneurin Bevan University | 2 | 119,915 (3.55) | 26,355 (4.37) | 16,269 (4.03) | 14,343 (4.92) | 3,509 (4.76) | 26,355 (4.31) |
| Aneurin Bevan University | 3 | 113,810 (3.37) | 23,251 (3.85) | 15,565 (3.86) | 11,495 (3.95) | 2,794 (3.79) | 23,251 (3.86) |
| Aneurin Bevan University | 4 | 83,692 (2.48) | 16,914 (2.8) | 11,669 (2.89) | 7,831 (2.69) | 1,951 (2.64) | 16,914 (2.82) |
| Aneurin Bevan University | 5 | 78,561 (2.32) | 15,055 (2.49) | 10,913 (2.7) | 6,243 (2.14) | 1,553 (2.1) | 15,055 (2.55) |
| Betsi Cadwaladr University | 1 | 82,259 (2.43) | 14,693 (2.43) | 8,351 (2.07) | 8,149 (2.8) | 2,913 (3.95) | 14,693 (2.22) |
| Betsi Cadwaladr University | 2 | 131,180 (3.88) | 24,608 (4.08) | 15,754 (3.9) | 11,998 (4.12) | 4,318 (5.85) | 24,608 (3.83) |
| Betsi Cadwaladr University | 3 | 173,949<br>(5.14) | 33,682 (5.58) | 23,405 (5.8) | 14,883 (5.11) | 5,385 (7.3) | 33,682 (5.34) |
| Betsi Cadwaladr University | 4 | 177,909<br>(5.26) | 34,694 (5.75) | 24,162 (5.99) | 14,959 (5.13) | 5,107 (6.92) | 34,694 (5.58) |
| Betsi Cadwaladr University | 5 | 152,397<br>(4.51) | 27,054 (4.48) | 18,756 (4.65) | 11,514 (3.95) | 4,044 (5.48) | 27,054 (4.34) |
| Cardiff and Vale University | 1 | 157,086<br>(4.65) | 20,509 (3.4) | 11,716 (2.9) | 12,301 (4.22) | 2,269 (3.08) | 20,509 (3.44) |
| Cardiff and Vale University | 2 | 79,573<br>(2.35) | 9,674 (1.6) | 5,867 (1.45) | 5,432 (1.86) | 1,053 (1.43) | 9,674 (1.63) |
| Cardiff and Vale University | 3 | 145,155<br>(4.29) | 9,346 (1.55) | 5,838 (1.45) | 4,792 (1.64) | 928 (1.26) | 9,346 (1.59) |
| Cardiff and Vale University | 4 | 85,007<br>(2.51) | 10,507 (1.74) | 7,270 (1.8) | 4,871 (1.67) | 914 (1.24) | 10,507 (1.81) |
| Cardiff and Vale University | 5 | 214,906<br>(6.36) | 29,388 (4.87) | 21,131 (5.23) | 12,233 (4.2) | 2,226 (3.02) | 29,388 (5.13) |
| Cwm Taf Morgannwg | 1 | 122,502<br>(3.62) | 24,678 (4.09) | 14,549 (3.6) | 14,884 (5.11) | 1,427 (1.93) | 24,678 (4.39) |
| Cwm Taf Morgannwg | 2 | 151,544<br>(4.48) | 30,238 (5.01) | 18,944 (4.69) | 17,100 (5.87) | 1,645 (2.23) | 30,238 (5.4) |
| Cwm Taf Morgannwg | 3 | 77,081<br>(2.28) | 12,372 (2.05) | 8,038 (1.99) | 6,653 (2.28) | 693 (0.94) | 12,372 (2.2) |
| Cwm Taf Morgannwg | 4 | 61,015 (1.8) | 12,350 (2.05) | 8,711 (2.16) | 5,812 (1.99) | 944 (1.28) | 12,350 (2.15) |
| Cwm Taf Morgannwg | 5 | 73,469<br>(2.17) | 13,186 (2.18) | 9,403 (2.33) | 5,823 (2) | 854 (1.16) | 13,186 (2.33) |
| Hywel Dda University | 1 | 32,424<br>(0.96) | 6,486 (1.07) | 3,810 (0.94) | 3,581 (1.23) | 1,171 (1.59) | 6,486 (1) |
| Hywel Dda University | 2 | 76,280<br>(2.26) | 14,883 (2.47) | 9,559 (2.37) | 7,400 (2.54) | 2,205 (2.99) | 14,883 (2.39) |
| Hywel Dda University | 3 | 133,041<br>(3.93) | 26,132 (4.33) | 17,970 (4.45) | 11,766 (4.04) | 3,469 (4.7) | 26,132 (4.28) |
| Hywel Dda University | 4 | 112,845<br>(3.34) | 22,137 (3.67) | 15,721 (3.89) | 9,347 (3.21) | 2,915 (3.95) | 22,137 (3.63) |
| Hywel Dda University | 5 | 47,112<br>(1.39) | 3,869 (0.64) | 2,701 (0.67) | 1,633 (0.56) | 508 (0.69) | 3,869 (0.63) |
| Powys Teaching | 1 | 7,126 (0.21) | 1,351 (0.22) | 865 (0.21) | 659 (0.23) | 212 (0.29) | 1,351 (0.21) |
| Powys Teaching | 2 | 6,325 (0.19) | 1,390 (0.23) | 956 (0.24) | 642 (0.22) | 200 (0.27) | 1,390 (0.22) |
| Powys Teaching | 3 | 20,504<br>(0.61) | 4,235 (0.7) | 2,969 (0.74) | 1,803 (0.62) | 509 (0.69) | 4,235 (0.7) |
| Powys Teaching | 4 | 31,762<br>(0.94) | 5,957 (0.99) | 4,145 (1.03) | 2,538 (0.87) | 718 (0.97) | 5,957 (0.99) |
| Powys Teaching | 5 | 3,483 (0.1) | 631 (0.1) | 444 (0.11) | 240 (0.08) | 87 (0.12) | 631 (0.1) |
| Swansea Bay University | 1 | 118,215 (3.5) | 26,537 (4.4) | 18,147 (4.5) | 13,123 (4.5) | 3,788 (5.13) | 26,537 (4.29) |
| Swansea Bay University | 2 | 90,292<br>(2.67) | 19,668 (3.26) | 13,953 (3.46) | 9,014 (3.09) | 2,654 (3.6) | 19,668 (3.21) |
| Swansea Bay University | 3 | 63,092<br>(1.87) | 13,337 (2.21) | 9,866 (2.44) | 5,741 (1.97) | 1,707 (2.31) | 13,337 (2.19) |
| Swansea Bay University | 4 | 57,348 (1.7) | 11,837 (1.96) | 8,788 (2.18) | 4,848 (1.66) | 1,574 (2.13) | 11,837 (1.94) |
| Swansea Bay University | 5 | 120,583<br>(3.57) | 22,319 (3.7) | 17,351 (4.3) | 7,891 (2.71) | 2,598 (3.52) | 22,319 (3.72) |
| Missing |  | 3,762 (0.11) | 173 (0.03) | 86 (0.02) | 90 (0.03) | 34 (0.05) | 173 (0.03) |

**Table 4.** Demographic information for study cohorts. Variable distributions are reported as n (%) unless otherwise specified. Percentages may not total 100 due to rounding.

|  |  | Pain cohorts |  |  |  |  |
| --- | --- | --- | --- | --- | --- | --- |
|  | General population comparator (non pain) group (% of cohort) | All pain cohorts (% of cohort) | Diagnosis code (% of cohort) | Prescription (% of cohort) | Pain service (% of cohort) | Not referred to pain services (% of cohort) |
| Total, n | 3,381,071 | 603,697 | 403,703 | 291,350 | 73,782 | 529,915 |
| **Age groups** |  |  |  |  |  |  |
| 0-10 | 792,792 (23.45) | 1,856 (0.31) | 1,684 (0.42) | 184 (0.06) | 55 (0.07) | 1,801 (0.34) |
| 11-20 | 513,214 (15.18) | 9,145 (1.51) | 6,353 (1.57) | 2,800 (0.96) | 1,487 (2.02) | 7,658 (1.45) |
| 21-30 | 564,726 (16.70) | 27,234 (4.51) | 13,050 (3.23) | 13,798 (4.74) | 6,203 (8.41) | 21,031 (3.97) |
| 31-40 | 415,413 (12.29) | 48,815 (8.09) | 23,894 (5.92) | 27,012 (9.27) | 11,012 (14.93) | 37,803 (7.13) |
| 41-50 | 370,644 (10.96) | 88,387 (14.64) | 53,318 (13.21) | 46,630 (16.00) | 16,384 (22.21) | 72,003 (13.59) |
| 51-60 | 285,739 (8.45) | 133,992 (22.20) | 96,776 (23.97) | 61,319 (21.05) | 16,246 (22.02) | 117,746 (22.22) |
| 61-70 | 222,049 (6.57) | 137,296 (22.74) | 102,686 (25.44) | 62,330 (21.39) | 12,387 (16.79) | 124,909 (23.57) |
| 71-80 | 128,430 (3.80) | 106,119 (17.58) | 75,902 (18.80) | 49,934 (17.14) | 7,579 (10.27) | 98,540 (18.60) |
| 81-90 | 73,283 (2.17) | 45,038 (7.46) | 27,317 (6.77) | 23,810 (8.17) | 2,275 (3.08) | 42,763 (8.07) |
| 91+ | 14,781 (0.44) | 5,815 (0.96) | 2,723 (0.67) | 3,533 (1.21) | 154 (0.21) | 5,661 (1.07) |
| **Age, median (IQR)** | 27.4 (20.09) | 60.47 (22.76) | 61.62 (20.2) | 60.07 (24.04) | 51.98 (23.08) | 61.53 (22.57) |
| **WIMD** |  |  |  |  |  |  |
| 1 (Most deprived) | 695,479 (20.57) | 128,455 (21.28) | 77,499 (19.20) | 72,415 (24.85) | 16,686 (22.62) | 111,769 (21.09) |
| 2 | 655,109 (19.38) | 126,816 (21.01) | 81,302 (20.14) | 65,929 (22.63) | 15,584 (21.12) | 111,232 (20.99) |
| 3 | 726,632 (21.49) | 122,355 (20.27) | 83,651 (20.72) | 57,133 (19.61) | 15,485 (20.99) | 106,870 (20.17) |
| 4 | 609,578 (18.03) | 114,396 (18.95) | 80,466 (19.93) | 50,206 (17.23) | 14,123 (19.14) | 100,273 (18.92) |
| 5 (Least deprived) | 690,511 (20.42) | 111,502 (18.47) | 80,699 (19.99) | 45,577 (15.64) | 11,870 (16.09) | 99,632 (18.80) |
| Missing | 3,762 (0.11) | 173 (0.03) | 86 (0.02) | 90 (0.03) | 34 (0.05) | 139 (0.03) |
| **Area** |  |  |  |  |  |  |
| Rural | 923,187 (27.30) | 186,701 (30.93) | 128,783 (31.90) | 84,225 (28.91) | 23,828 (32.30) | 366,903 (69.24) |
| Urban | 2,454,122 (72.58) | 416,823 (69.05) | 274,834 (68.08) | 207,035 (71.06) | 49,920 (67.66) | 162,873 (30.74) |
| Missing | 3,762 (0.11) | 173 (0.03) | 86 (0.02) | 90 (0.03) | 34 (0.05) | 139 (0.03) |
| **Sex** |  |  |  |  |  |  |
| Male | 1,746,528 (51.66) | 239,587 (39.69) | 155,817 (38.60) | 112,993 (38.78) | 26,064 (35.33) | 213,523 (36.01) |
| Female | 1,634,490 (48.34) | 364,110 (60.31) | 247,886 (61.40) | 178,357 (61.22) | 47,718 (64.67) | 316,392 (53.36) |
| Missing | 53 (0.00) |  |  |  |  |  |
| **Charlson score** |  |  |  |  |  |  |
| Min | -1 | -1 | -1 | -1 | -1 | -1 |
| Max | 78 | 84 | 78 | 84 | 72 | 84 |
| Median | 0 | 0 | 0 | 0 | 0 | 0 |
| IQR | 0 | 4 | 0 | 4 | 4 | 0 |
| **eFI score** |  |  |  |  |  |  |
| Fit | 3,155,855 (93.34) | 357,645 (59.24) | 244,066 (60.46) | 158,796 (54.50) | 44,712 (60.60) | 312,933 (59.05) |
| Mild | 167,995 (4.97) | 189,550 (31.40) | 126,278 (31.28) | 99,149 (34.03) | 23,114 (31.33) | 166,436 (31.41) |
| Moderate | 36,029 (1.07) | 47,240 (7.83) | 28,508 (7.06) | 27,619 (9.48) | 5,104 (6.92) | 42,136 (7.95) |
| Severe | 7,353 (0.22) | 9,262 (1.53) | 4,851 (1.20) | 5,786 (1.99) | 852 (1.15) | 8,410 (1.59) |
| Missing | 13,839 (0.41) |  |  |  |  |  |

#### 3.2.1 Age

Within-group analysis showed that there was a statistically significant difference between the age group categories (p<0.0001) across all cohorts. Additionally, statistically significant differences in age were observed between the combined pain group and the comparator group (p<0.0001), as well as between the pain service group and those who were not (p < 0.0001). Individuals in the pain cohorts were older than the comparator cohort: in the diagnoses and prescription pain cohorts, the largest proportion of individuals was within the 61-70 age group (25.44% and 21.39%, respectively), whereas the largest percentage of individuals in the pain services group was in the 51-60 age range (22.02%).

#### 3.2.2 Deprivation

Within-group analyses showed statistically significant differences in WIMD quintile distribution across all cohorts (p < 0.0001). Between-group comparisons also revealed significant differences in WIMD between the combined pain group and the comparator group, and between individuals referred to pain services and those not referred (both p < 0.0001). Most pain cohorts had a higher proportion of individuals from more deprived areas, particularly WIMD quintile 1 (most deprived). For example, in the combined pain cohort, 21.28% of individuals were from the most deprived quintile, and the lowest percentage of individuals were in the least deprived quintile (18.47%). The exception was the diagnosis cohort, where WIMD distribution was more evenly spread across quintiles.

#### 3.2.3 Geographical area

Within-group analyses showed statistically significant differences in area of residence (urban/rural) across all cohorts (p < 0.0001). Between-group comparisons also indicated significant differences in area of residence between the combined pain group and the comparator group, and between those referred to pain services and those not referred (both p < 0.0001). Across all cohorts, the majority of individuals lived in urban areas. For example, 69.05% of individuals in the combined pain cohort lived in urban areas; this was 72.58% in the comparator group, while the proportion living in rural areas was comparatively lower across all cohorts.

#### 3.2.4 Sex

Within-group analyses showed statistically significant differences in sex distribution for all cohorts (p < 0.0001). Between-group comparisons also revealed significant differences in sex between the combined pain group and the comparator group, and between those referred to pain services and those not referred (p < 0.0001). Across all pain cohorts, a higher proportion of individuals were female. For example, in the combined pain cohort, 60.31% were female compared to 48.34% in the comparator group.

#### 3.2.5 Charlson Comorbidity Index Score

Between-group comparison showed a significant difference between the combined pain group and the comparator group (p < 0.0001), with higher comorbidity observed in the combined pain group (maximum score 84) compared with the comparator group (maximum score 78). There was no difference in Charlson scores between those referred to pain services and those not referred (p = 0.1055). Median scores were the same for all cohorts (score of 0). The interquartile range (IQR) was 4 for the combined pain, prescription, and the pain service cohort, while it was 0 for the remaining cohorts, indicating a slightly greater variability in these groups. The pain service cohort had the lowest maximum score, with a value of 72.

#### 3.2.6 eFI

Within-group analyses showed statistically significant differences across all categories of eFI in all cohorts (p < 0.0001). Between-group comparisons also showed significant differences in eFI between the combined pain group and the comparator group, and between those referred to pain services and those not referred (p < 0.0001). The majority of the comparator group (93.94%) was classed as ‘fit’ according to their eFI score, while approximately between 50-60% of individuals in the pain cohorts were categorised similarly (diagnoses cohort: 60.46%, prescription: 54.50%, pain service: 60.60%). Between 31.28% and 34.03% of pain cohorts were classified with mild frailty compared with only 4.97% of the comparator cohort. Only 1.07% of the general comparator cohort were classified as moderately frail, while this percentage in the pain cohorts varied from 7.06% (pain service) to 9.48% (diagnoses). The percentage of individuals classed as having severe frailty was highest in the prescription cohort (1.99%) and the lowest was in the general population comparator cohort (0.22%). Among the pain cohorts, the pain service cohort appeared to be least frail.

#### 3.2.7 Healthcare utilisation

We calculated five different healthcare utilisation measures: GP events, hospital admissions, emergency hospital admissions, emergency attendances, and outpatient attendances. Table 5 gives the summary statistics for the healthcare utilisation for each cohort for the study duration. The counts for the healthcare utilisation taken on the 1^st^ of January of each study year can be found in Appendix 6. Note that individuals in the prescription cohort are removed from the cohort when they have no active prescription period, whereas individuals in the diagnosis and pain service cohorts remain until they leave Wales, a SAIL-providing GP, or at death.

**Table 5.** Summary statistics for healthcare utilisation for all cohorts.

|  |  | Pain cohorts |  |  |  |  |
| --- | --- | --- | --- | --- | --- | --- |
|  | General population comparator (non pain) group | All pain cohorts | Diagnosis code | Prescription | Pain service | Non-pain service |
| Total cohort n = | 3,381,071 | 603,697 | 403,703 | 291,350 | 73,782 | 529,915 |
| **Total HCU for the study period** |  |  |  |  |  |  |
| GP events | 366,468,782 | 158,301,483 | 110,828,339 | 27,950,561 | 24,525,932 | 133,775,551 |
| Hospital admissions | 6,890,068 | 3,132,374 | 2,118,541 | 579,622 | 502,283 | 2,630,091 |
| Hospital emergency admissions | 2,668,455 | 1,158,496 | 763,365 | 210,990 | 185,894 | 972,602 |
| ED attendances | 7,968,627 | 2,068,638 | 1,385,097 | 352,715 | 356,731 | 1,711,907 |
| Outpatient attendances | 30,171,711 | 13,969,031 | 9,846,640 | 2,573,979 | 2,471,667 | 11,497,364 |
| **Median monthly HCU** |  |  |  |  |  |  |
| GP events | 2,091,332 | 957,399 | 668,900 | 1,767,389 | 155,050 | 809,310 |
| Hospital admissions | 41,265 | 18,980 | 127,90 | 3,780 | 3,112 | 15,852 |
| Hospital emergency admissions | 15,773 | 7,126 | 4,642 | 1,344 | 1,200 | 5,945 |
| ED attendances | 48,365 | 13,111 | 8,545 | 2,216 | 2,309 | 10,771 |
| Outpatient attendances | 178,338 | 85,070 | 59,546 | 16,415 | 15,388 | 69,942 |
| **Monthly HCU IQR** |  |  |  |  |  |  |
| GP events | 512,142 | 279,808 | 161,288 | 368,689 | 57,166 | 222,517 |
| Hospital admissions | 4,841 | 5,085 | 3,035 | 1,266 | 976 | 4,092 |
| Hospital emergency admissions | 2,153 | 1,926 | 1,087 | 295 | 425 | 1,486 |
| ED attendances | 7,219 | 3,689 | 2,134 | 600 | 842 | 2,952 |
| Outpatient attendances | 28,175 | 22,046 | 11,963 | 5,434 | 5,207 | 16,492 |
| **Total HCU per 1000 people for the study period** |  |  |  |  |  |  |
| GP events | 108,388 | 262,220 | 274,529 | 95,934 | 332,411 | 252,447 |
| Hospital admissions | 2,038 | 5,189 | 5,248 | 1,989 | 6,808 | 4,963 |
| Hospital emergency admissions | 789 | 1,919 | 1,891 | 724 | 2,520 | 1,835 |
| ED attendances | 2,357 | 3,427 | 3,431 | 1,211 | 4,834 | 3,231 |
| Outpatient attendances | 8,924 | 23,139 | 24,391 | 6,577 | 33,500 | 21,697 |

After controlling for age, sex, deprivation level, and seasonality, plus offsetting for changes in the size of the population at risk, the monthly trends in rates of healthcare utilisation for the combined pain cohort were as follows:

- GP event rates were increasing by 0.0052% (RR 1.000052, 95% CI: 1.000050 to 1.000053, p < 0.0001).
- Hospital admission rates were increasing by 0.0012% (RR 1.000012, 95% CI: 1.0000097 to 1.000015, p < 0.0001).
- Emergency hospital admission rates showed no statistically significant change over time, decreasing by -0.00030% (RR 0.999997, 95% CI: 0.9999956 to 1.0000039, p = 0.89).
- Emergency department attendance rates were increasing by 0.00059% (RR 1.0000059, 95% CI: 1.0000026 to 1.0000091, p < 0.0001).
- Outpatient attendance rates were decreasing by 0.0018% (RR 0.999982, 95% CI: 0.999981 to 0.999984, p < 0.0001).

See coefficients from this model in Appendix 7.

We further assessed the difference in healthcare utilisation between the combined pain and the comparator cohorts adjusted for age, sex, deprivation level, seasonality, and changes in the size of the population at risk. Results from the regression models showed that:

- There was a 62.60% increased risk of GP events in the combined pain cohort (all three groups) compared to the comparator group (RR 1.626, 95% CI 1.625 to 1.627, p < 0.0001).
- There was a 2.8% decreased risk of hospital admission in the combined pain cohort compared to the comparator group (RR 0.972, 95% CI: 0.971 to 0.973, p < 0.0001).
- There was a 0.9% increased risk of emergency hospital admission in the combined pain cohort compared to the comparator group (RR 1.009 95% CI: 1.006 to 1.011, p < 0.0001).
- There was a 1.9% increased risk of emergency department attendance in the combined pain cohort compared to the comparator group (RR 1.019, 95% CI: 1.018 to 1.021, p < 0.0001).
- There was a 2.7% increased risk of outpatient attendance rates in the combined pain cohort compared to the comparator group (RR 1.027, 95% CI: 1.026 to 1.028, p < 0.0001).

See coefficients from this model in Appendix 8.

For all those who were included in the persistent pain cohort, we also compared healthcare utilisation between individuals who accessed outpatient pain services and those who did not, adjusting for age, sex, deprivation level, seasonality, and population at risk. We found that:

- There was a 32.8% increased risk of a GP event in the pain services group compared to non-pain services group (RR 1.328 95% CI: 1.325 to 1.330, p < 0.0001).
- There was a 4.41% decreased risk of hospital admission in the pain services group compared to the non-pain services group (RR 0.956 95% CI: 0.953 to 0.959 p <0.0001).
- There was a 1.00% increase in the risk of emergency hospital admission in the pain services group compared to non-pain services group (RR 1.01 95% CI: 1.005 to 1.015, p = 0.0001).
- There was a 2.02% increased risk of emergency department attendance in the pain services group compared to the non-pain services group (RR 1.020 95% CI: 1.016 to 1.024, p = 0.001).
- There was a 2.33% increased risk of outpatient department attendance in the pain services group compared to the non-pain services group (RR 1.023 95% CI: 1.022 to 1.025, p < 0.0001).

See coefficients from this model in Appendix 9.

Figure 4 shows the monthly rates of healthcare utilisation per 1000 people for all pain cohorts (in black) and the general comparator cohort (in red). From the graphs, we can see that the rates per 1000 people for pain cohorts are generally higher and are increasing at a greater rate than for the comparator cohort for each of the healthcare utilisation measures.

**Figure 4.**
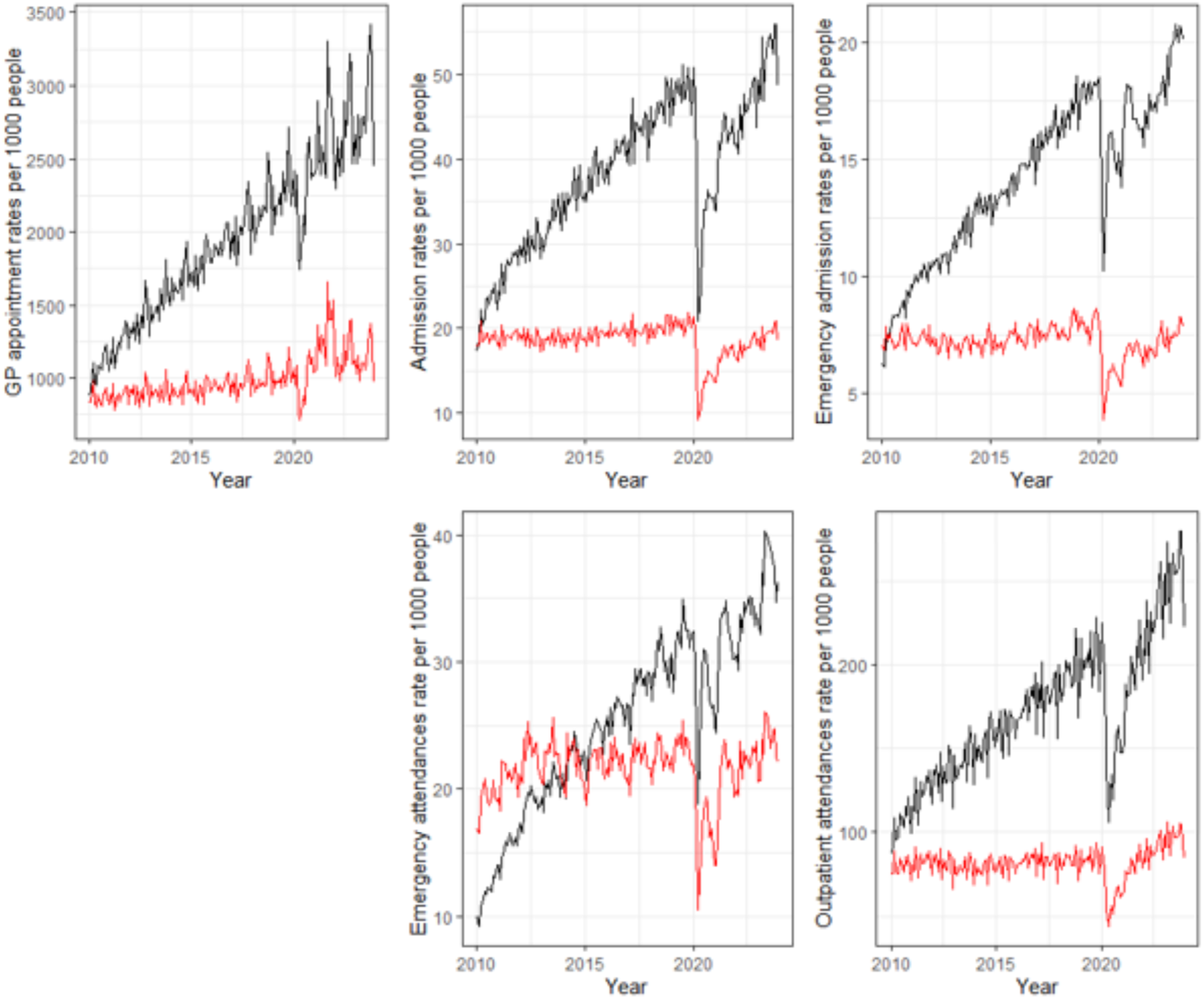
Graphs of the rates per 1000 people for healthcare utilisation 2010-2023. People who have experienced persistent pain are in black, while the red represents the comparator cohort, i.e. those who have not experienced persistent pain.

Figure 4 shows the negative binomial model estimate (in red) and associated prediction intervals (in grey) for the healthcare utilisation of all those with persistent pain. Observed counts are represented in black. Aside from the noticeable drop in healthcare utilisation during the COVID-19 pandemic, all healthcare resource utilisation lies within the expected range as represented by the prediction intervals, and illustrates an upward trend in healthcare utilisation over time.

**Figure 5.**
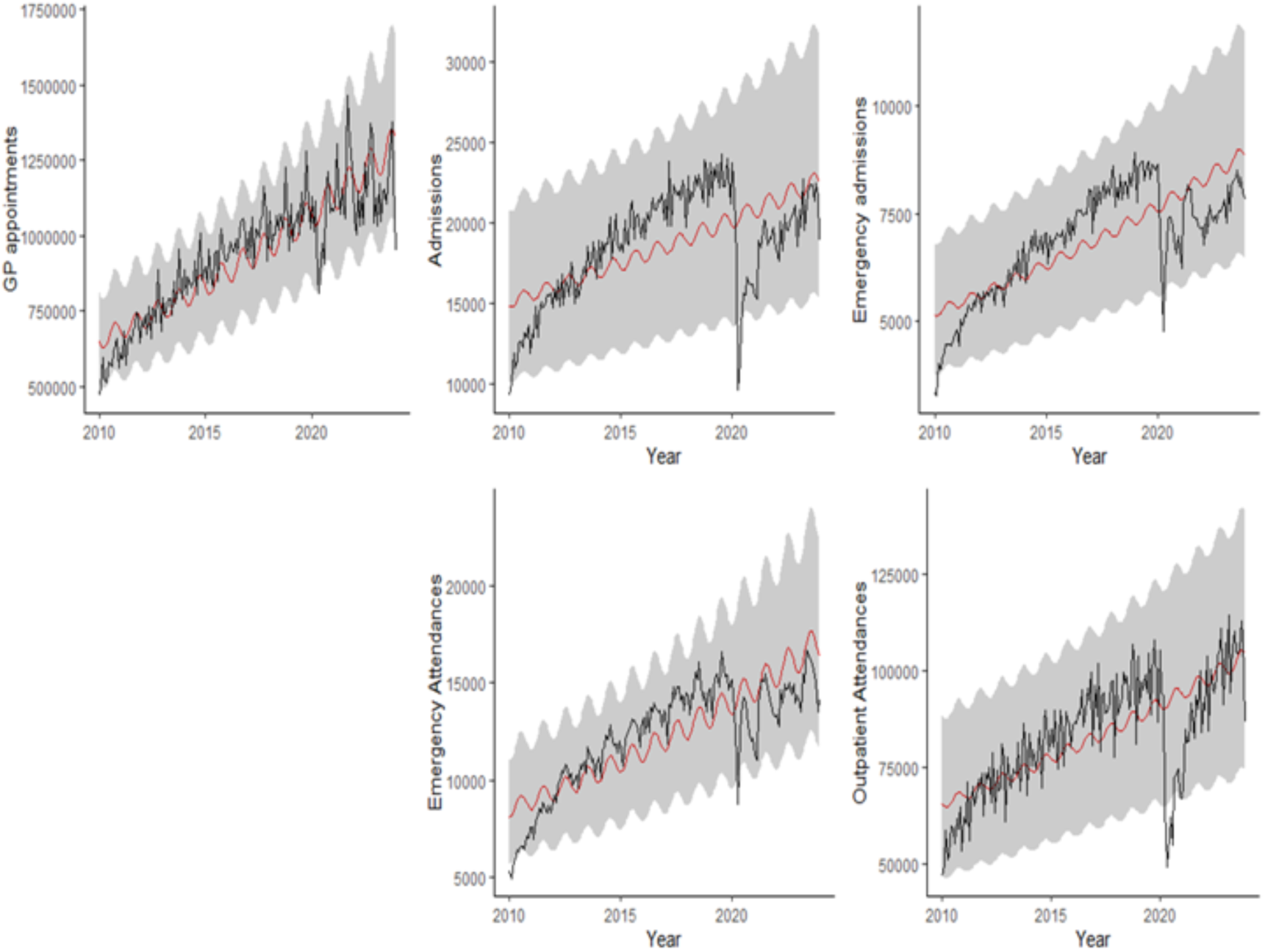
Negative binomial model estimate (in red) and associated prediction intervals (in grey) for the healthcare utilisation of all those with persistent pain. Black represents actual counts.

Appendix 10 gives the time decomposition plots for each of the healthcare utilisation measures. After removing random and seasonal components from the time series, each trend plot shows an increase in healthcare utilisation during the study period (aside from during the COVID-19 pandemic). Each plot shows a clear upward trend for each of the healthcare utilisation measurements with a noticeable seasonal effect for GP events and emergency attendances.

## 4.1 Discussion

This retrospective linked-data study aimed to identify individuals in Wales living with persistent pain and describe associated epidemiological trends. Our study included 3,984,768 individuals, with 603,697 in the three pain cohorts combined (diagnoses, prescriptions, pain services) and 3,381,071 in the general population comparator cohort. Approximately 15% of the general population met the criteria for persistent pain, with the largest cohort defined by diagnostic codes (403,705 individuals), followed by those prescribed pain medication for three months or more (291,350 individuals) and those referred to specialist outpatient pain services (73,782 individuals). Given the absence of a dedicated persistent pain register, this approach of using multiple definitions of persistent pain sought to capture as many individuals living with persistent pain as possible.

Our analysis showed a small but statistically significant decrease in the yearly rate of individuals defined as living with persistent pain during our study period. Individuals living with persistent pain tended to be in the older age groups, with the diagnosis and prescription group primarily aged 61-70. The pain service group, on the other hand, was younger, with the majority falling within the 51-60 age group. Individuals living with persistent pain tended to live in more deprived areas, and the majority of both the persistent pain group and the general population resided in urban, as opposed to rural, areas. The persistent pain group was less urban than the general population comparator group.

There was an over-representation of women identified as living with persistent pain, with the difference being more pronounced in the pain service group. Individuals with persistent pain had more comorbid conditions than the general population without persistent pain. While no statistically significant difference was found between the Charlson scores of those referred to pain services and those who were not (median scores of 72 vs. 84, respectively), this difference may be clinically significant. With regards to eFI, individuals living with persistent pain were more frail than the general population. In addition, the pain services group had a higher proportion of individuals in the ’fit’ category and fewer individuals in the ’moderate’ and ’severe’ categories compared to the other two pain cohorts.

We calculated five different healthcare utilisation measures: General Practitioner (GP) appointments, hospital admissions, emergency hospital admissions, emergency attendances, and outpatient attendances. After controlling for age, sex, deprivation level, seasonality, and the population at risk, we found statistically significant increases in the monthly rates of GP appointments, hospital admissions, and emergency department attendances, while outpatient attendances showed a statistically significant decrease. There was a substantial and statistically significant difference in the rate of GP appointments for individuals with persistent pain compared to the general population without persistent pain (62.60% greater for the persistent pain group). Emergency hospital attendances were 0.9% higher, emergency department attendances were 1.9% higher, outpatient department attendances were 2.7% higher. Hospital admissions were 2.8% lower. All differences were statistically significant.

When comparing individuals in the persistent pain cohort who were referred to specialist outpatient services to those who were not, rates of GP appointments were 32.80% higher in the pain service group, emergency hospital admissions were 1% higher, emergency department attendances were 2.05% higher, and outpatient attendances were 2.31% higher. However, hospital admissions were 4.41% lower in the pain service group. These differences were statistically significant.

The prevalence of persistent pain in Wales, as calculated in this study, is considerably lower than that reported in other UK studies. A systematic review by Fayaz et al (1) found that between one-third and one-half of the UK population are affected by persistent pain. The studies included in this review, however, gathered data on pain status using self-reported surveys. Self-reported studies are often found to identify a higher prevalence of health conditions compared to EHRs (17). This suggests that not all individuals who experience persistent pain are seeking medical assistance—potentially because their pain is less severe, occurs less consistently or is less specific in nature. Versus Arthritis (18) estimated in their report that 12% of the population experiences high-impact persistent pain (where people are unable to carry out their daily activities). The prevalence of 15% identified in our study may be capturing those with more severe or disabling forms of persistent pain.

Conversely, given that older people are more likely to experience persistent pain (as found by our study), the disparity between EHR and self-reported prevalence data may reflect issues that disproportionately affect older populations when accessing healthcare. These could include barriers caused by comorbid medical conditions (19), extrinsic factors such as cost, transportation difficulties, and reliance on care givers (20), or digital exclusion (21). If so, this may have led to an underestimation of persistent pain prevalence in older populations in our data.

Our analysis found that the rate of individuals experiencing persistent pain decreased each year, which may seem contrary to expectations given the ageing population. Several factors could explain this trend. Firstly, this could be a result of GP coding practices changing over time. A qualitative study involving healthcare professionals in Wales highlighted the complexity of coding in primary care.

Participants reported that systems lacked intuitiveness, and that multimorbidities and time pressures—issues particularly prominent in deprived areas—negatively affected coding practices and often led to underreporting of diseases in these areas (22). Wales especially has experienced many shifts in how GPs record data on health conditions. The Quality and Outcomes Framework (QoF) has undergone significant changes in the last decade (23). With the removal of financial incentives for recording diagnostic data, coding practices have become less consistent, particularly in areas such as multimorbidity (24). In addition, if persistent pain is caused by another long-term condition, then only the primary condition may be recorded with persistent pain being viewed instead as a symptom. Given that persistent pain is strongly associated with multimorbidity and long-term conditions (25), coding for persistent pain in general practices may have declined.

In secondary care, ICD-10 codes do not capture persistent pain well, hence the much-needed development of ICD-11 codes in this area (26). Adoption of this system should improve diagnosis and data recording efficiency; however, the UK has not yet mandated its use in secondary care (27). There is currently no publically available information indicating that Wales has begun implementing IVD-11 in secondary care settings. It will likely be decades before these codes can be used for longitudinal research in this country.

The decline in persistent pain diagnoses over time observed in our data could also be linked to a reduced reliance on long-term opioid prescription, especially post COVID-19. Despite efforts by primary care health professionals to reduce opioid prescribing in Wales, there was a 30% increase in 2018/2019 compared with 2008/2009 (27). Long-term opioid prescribing rates increased again during the pandemic (28,29). This rise promoted efforts by national bodies to take action, due to the increased risk of adverse events associated with prolonged use of these drugs. The Medicines and Healthcare products Regulatory Agency (MHRA) (30) and National Institute for Health and Care Excellence (NICE) (31) issued recommendations highlighting the risk of addiction from opioid use. These efforts showed positive results: in 2023, NHS England reported a reduction in opioid prescriptions by 450,000 (32). Consequently, more emphasis has been placed on non-pharmaceutical interventions like physiotherapy and cognitive behavioural therapy (CBT) (33), which may not be fully reflected in our data.

The decline in the rates of individuals meeting our definition for persistent pain may also be partially attributed to reduced access to general practitioners. Limited availability of appointments, long waiting times, and other barriers to healthcare access, including those mentioned above, may have contributed to fewer referrals to specialist outpatient services, as individuals may have been unable to see their GP or sought care less frequently (34). However, limited access resulted from a year-on-year rise in appointments, meaning referrrals have increased proportionaly due to demand (35). Thus, the decrease in the pain service cohort may not be explained by a reduction in referrals. Rather, it is possible that, due to limited access to public services, people are opting to pay for private pain services instead (36).

More optimistically, this reduction in rates could be reflecting real changes in population health. People may be managing pain better due to interventions and lifestyle changes—many interventions for pain have been found to be effective that are accessible outside of the healthcare service, such as exercise (37), mindfulness and meditation (38), acupuncture (39) and online CBT programmes (39).

Our analysis found that individuals living with persistent pain were older, more likely to be female, and more commonly live in higher levels of deprivation compared to the general population without persistent pain. They also have higher levels of comorbidity, as indicated by a greater maximum Charlson score, and demonstrate increased levels of frailty, with far fewer individuals classified as ‘fit’ according to the eFI score.

These findings are consistent with existing literature. A review by Fayaz et al (1) found that persistent pain increases with age, ranging from 14.3% in individuals aged 18-25 years to 62% in those over 75 years. The Health Survey for England reported persistent pain prevalence of 16% among those aged 16-24 years rising to 53% in those aged 75 years and over (41). Older adults are more likely to experience comorbid conditions associated with chronic pain, such as diabetes and osteoarthritis (42–44). Ageing is also strongly associated with polypharmacy (45), which may exacerbate persistent pain, or contribute to conditions that are linked with pain such as frailty and poor physical functioning (46). Despite being more likely to experience persistent pain, older adults are less likely to receive adequate pain management (47). This under-treatment may be contributing to the increasing prevalence of persistent pain with age, as poorly managed pain can worsen over time.

Socioeconomic deprivation also plays a significant role in the experience and prevalence of pain, supporting the findings in our report. People living in deprived areas are more likely to experience chronic health conditions such as musculoskeletal disorders and diabetes, which are closely linked to persistent pain (48). Barriers to timely and effective health services are common in more deprived areas and can result in delays in diagnosis and treatment of health conditions, and thus, their exacerbation (49). Furthermore, greater exposure to manual labour, psychosocial stress, and mental health challenges in deprived areas can increase both the risk and severity of persistent pain (1). These interacting factors potentially contribute to the higher burden of persistent pain in deprived areas.

In line with our findings, which showed a higher proportion of women in our persistent pain cohort, existing evidence consistently demonstrates an overrepresentation of women among those living with persistent pain. Women are more likely to experience certain types of painful conditions, such as fibromyalgia, migraines, and musculoskeletal pain (50), and reproductive conditions such as endometriosis, menstrual pain, and pelvic inflammatory disease (51). In addition, women are generally more likely to seek medical help than men, and potentially also to receive prescription treatment for pain, which may contribute to the overrepresentation of women in our persistent pain cohort. As such, the gender disparity observed in our findings could be, at least in part, influenced by differences in healthcare-seeking behaviour, and the medical management of women’s problems by healthcare professionals, rather than true differences in pain prevalence (52).

Higher levels of frailty and comorbid conditions in our persistent pain cohort are likely due to the complex interaction between persistent pain and overall health status. This relationship is supported by findings in previous studies (53,54). Persistent pain can contribute to reduced mobility, physical inactivity, poor sleep, and psychological distress, all of which can accelerate frailty and worsen existing health conditions. As mentioned above, individuals with multiple long-term conditions—such as diabetes, cardiovascular disease, or osteoarthritis—may be more likely to develop persistent pain as a symptom or complication of those conditions (4,44,48). This bidirectional relationship means that people living with persistent pain often experience a decline in their functional capacity and this can contribute to frailty (55).

GP events were substantially higher in the persistent pain cohort compared to the general population without persistent pain. In the UK, GPs are typically the first point of contact for individuals experiencing persistent pain. Given the complex, evolving, and ongoing nature of persistent pain, it is unlikely that issues are identified or resolved in a single consultation. As a result, repeat visits are often necessary to manage symptoms effectively (33). The majority of people living with persistent pain in the UK (69%) had to visit multiple GPs and wait more than a year before receiving an official diagnosis, with more than one quarter of people (28%) waiting three years or more (56).

Despite the decrease in the number and rate of people living with persistent pain during the study period, the rate of GP events of those living with persistent pain increased steeply compared to the general population without persistent pain. This could reflect a worsening of long-term conditions over time, leading to more frequent GP appointments and prescriptions as people manage additional symptoms and comorbidities. For instance, in their report Versus Arthirits (18), it was found that the proportion of young people experiencing high-impact persistent pain increased from 21% to 32% between 2011 and 2017. Furthermore, NICE (33) advised healthcare professionals to schedule regular reviews for those under pain management plans to evaluate their effectiveness and to adjust plans if necessary, which would also result in an increase in GP service utilisation.

The pain service group consisted of a small percentage (12.22%) of the total pain cohort. Interestingly, GP appointments, emergency department attendance, and outpatient appointments were significantly higher in the pain service group compared to those in the persistent pain cohort who were not referred. Several factors may explain this. Long wait times to access specialist pain services may have led people to seek care elsewhere within the NHS. Also, the pain service group may experience more severe forms of pain, requiring the need for specialist services and thus more frequent use of healthcare resources. The pain service group was significantly younger, had a greater overrepresentation of women, and was less frail. Individuals in the pain service group may be more proactive in managing their health and have greater awareness of red flag symptoms—being referred to outpatient services and attendance to the emergency department could reflect a pattern of more frequent healthcare engagement, GP visits included. It is well documented that women are more likely to seek medical help than men (57,58), therefore, the higher rate of healthcare utilisation in the pain service group may, in part, be explained by the greater proportion women in this group. Conversely, the older age and poorer health of the non-pain service group, may be an indication of barriers in accessing care (19,20,21), which may suggest that they are not receiving the specialist care that they need. The non-pain services group did however have an increased rate of hospital admissions compared to those accessing pain management services, which may reflect the poorer health of this group and/or poorer management of their pain outside on inpatient settings.

## 4.2 Limitations

In the absence of a national pain register, it was not possible to confirm whether we have captured all individuals who are living with persistent pain in Wales. There is potential for misclassification or under-recording given the data sources, as not all individuals living with persistent pain in Wales will seek treatment from health services. It was also not possible to capture data on pain severity. Given that this is a retrospective observational study, no causal relationships could be determined. However, this study is strengthened by the use of three distinct and clinically relevant definitions of persistent pain, which allowed for a more comprehensive identification of individuals. In addition, the large sample size—covering over 86% of the population— enhances the reliability and generalisability of the findings within the study region.

## 4.3 Conclusion

This study provides valuable insight into the prevalence and healthcare utilisation patterns of individuals living with persistent pain in Wales. The findings also underscore the complexity of factors such as age, sex, deprivation, and comorbidities in the experience of persistent pain. It also highlights the increased healthcare burden this population faces, which has implications for NHS capacity and service planning. However, to gain a more accurate and complete understanding of the current situation in Wales, linked data needs to be supplemented with additional data collection methods, such as patient-reported outcomes (e.g. functional abilities, activities of daily living), over the counter medications, and data on non-pharmaceutical interventions. Future research should also explore the potential barriers to healthcare access, particularly in older populations and those living in deprived areas, to ensure that all individuals with persistent pain receive the necessary care and support.

## Data Availability

The data used in this study are available in the SAIL Databank at Swansea University, Swansea, UK, but as restrictions apply, they are not publicly available. All proposals to use SAIL data are subject to review by an independent Information Governance Review Panel (IGRP). Before any data can be accessed, approval must be given by the IGRP.  

## Additional Information

### Availability of data and materials

The data used in this study are available in the SAIL Databank at Swansea University, Swansea, UK, but as restrictions apply, they are not publicly available. All proposals to use SAIL data are subject to review by an independent Information Governance Review Panel (IGRP). Before any data can be accessed, approval must be given by the IGRP. The IGRP carefully considers each project to ensure the proper and appropriate use of SAIL data. When access has been granted, it is gained through a privacy-protecting trusted research environment (TRE) and remote access system referred to as the SAIL Gateway. SAIL has established an application process to be followed by anyone who would like to access data via SAIL at https://saildatabank.com/data/apply-to-work-with-the-data/.

### Ethical approval and consent to participate

Approval for the use of anonymised data in this study, provisioned within the Secure Anonymised Information Linkage (SAIL) Databank, was granted by an independent Information Governance Review Panel (IGRP) under project 1689. The IGRP has a membership comprised of senior representatives from the British Medical Association (BMA), the National Research Ethics Service (NRES), Public Health Wales and Digital Health and Care Wales (DHCW). The usage of additional data was granted by each respective data owner. The SAIL Databank is compliant with General Data Protection Regulations (GDPR) and the UK Data Protection Act.

## Abbreviations

ALF: Anonymous Linking Field
CBT: Cognitive Behavioural Therapy
EDDS: Emergency Department Data Set
EHR: Electronic Health Record
eFI: electronic Frailty Index
GP: General Practitioner
HRU: Health Resource Utilisation
ICD-10: International Classification of Diseases, 10th Revision
ICD-11: International Classification of Diseases, 11th Revision
IQR: Interquartile Range
LSOA: Lower-layer Super Output Area
NHS: National Health Service
NICE: National Institute for Health and Care Excellence
ONS: Office for National Statistics
OPDW: Outpatient Database for Wales
OPRD: Outpatient Referral Dataset
PEDW: Patient Episode Database for Wales
RALF: Residential Anonymous Linking Field
SAIL: Secure Anonymised Information Linkage Databank
SD: Standard Deviation
WDSD: Welsh Demographic Service Dataset
WIMD: Welsh Index of Multiple Deprivation
WLGP: Welsh Longitudinal General Practice dataset

## Acknowledgements

The authors would like to thank Emma Davies, Owen Hughes and Libby Humphris for their Çme, experÇse, and contribuÇons during stakeholder meeÇngs in guiding the focus of the research and interpretaÇon of findings.

## Appendices

## Appendix 1 Read codes considered to be referring to persistent pain, developed through discussion with project team members and stakeholders

| READ_CODE | PREF_TERM_60 |
| --- | --- |
| F29y5 | Chronic post-traumatic headache |
| N03x9 | Arthritis associated with other disease, hip |
| N0631 | Climacteric arthritis of the shoulder region |
| N05zR | Osteoarthritis NOS, of other tarsal joint |
| N05z1 | Osteoarthritis NOS, of shoulder region |
| 8HVk. | Private referral to pain management service |
| C108C | Insulin dependent diabetes mellitus with polyneuropathy |
| N0524 | Localised, secondary osteoarthritis of the hand |
| N0544 | Oligoarticular osteoarthritis, unspecified, of hand |
| N05zG | Osteoarthritis NOS, of PIP joint of finger |
| C373F | Familial amyloid polyneuropathy type II |
| Ryu10 | [X]Pain localised to other parts of lower abdomen |
| N0662 | Unspecified monoarthritis of the upper arm |
| N0433 | Monarticular juvenile rheumatoid arthritis |
| N05zA | Osteoarthritis NOS, of sternoclavicular joint |
| N05z4 | Osteoarthritis NOS, of the hand |
| N03xF | Arthritis associated with other disease, talonavicular joint |
| N0452 | Juvenile arthritis in psoriasis |
| N0543 | Oligoarticular osteoarthritis, unspecified, of forearm |
| N0230 | Gouty arthritis of unspecified site |
| N03x8 | Arthritis associated with other disease, DIP joint of finger |
| N05z8 | Osteoarthritis NOS, other specified site |
| N242z | Neuralgia, neuritis or radiculitis NOS |
| N03xB | Arthritis associated with other disease, knee |
| N051E | Localised, primary osteoarthritis of toe |
| N0510 | Localised, primary osteoarthritis of unspecified site |
| F364. | Idiopathic progressive polyneuropathy |
| N0634 | Climacteric arthritis of the hand |
| N0545 | Oligoarticular osteoarthritis, unspecified, of pelvis/thigh |
| Nyu01 | [X]Arthritis+polyarthritis due/other specfd bacterial agents |
| F301. | Other specified trigeminal neuralgia |
| N05zF | Osteoarthritis NOS, of MCP joint |
| N01z2 | Infective arthritis NOS, of the upper arm |
| N01z7 | Infective arthritis NOS, of the ankle and foot |
| N0454 | Juvenile arthritis in ulcerative colitis |
| Nyu00 | [X]Other streptococcal arthritis and polyarthritis |
| N040J | Rheumatoid arthritis of other tarsal joint |
| F3360 | Phantom limb syndrome with pain |
| J6710 | Alcohol-induced chronic pancreatitis |
| N0514 | Localised, primary osteoarthritis of the hand |
| 1D19. | Pain in lymph nodes after alcohol consumption |
| N01zR | Infective arthritis NOS, of 1st MTP joint |
| N05zM | Osteoarthritis NOS, of tibio-fibular joint |
| N054z | Osteoarthritis of more than one site, unspecified, NOS |
| N0525 | Localised, secondary osteoarthritis of pelvic region/thigh |
| N063z | Climacteric arthritis NOS |
| N0628 | Allergic arthritis of other specified site |
| N051z | Localised, primary osteoarthritis NOS |
| N0432 | Pauciarticular juvenile rheumatoid arthritis |
| N01zP | Infective arthritis NOS, of talonavicular joint |
| 9mM3. | Rheumatoid arthritis monitoring verbal invitation |
| N0532 | Localised osteoarthritis, unspecified, of the upper arm |
| N0668 | Unspecified monoarthritis of other specified site |
| F371. | Polyneuropathy in collagen vascular disease |
| F3720 | Acute painful diabetic neuropathy |
| N200. | Giant cell arteritis with polymyalgia rheumatica |
| N03xK | Arthritis associated with other disease, IP joint of toe |
| N0104 | Pyogenic arthritis of the hand |
| 7P203 | Delivery of rehabilitation for rheumatoid arthritis |
| N040A | Rheumatoid arthritis of DIP joint of finger |
| A5312 | Postherpetic trigeminal neuralgia |
| N0103 | Pyogenic arthritis of the forearm |
| 9mM1. | Rheumatoid arthritis monitoring invitation second letter |
| N01z3 | Infective arthritis NOS, of the forearm |
| 38GJ1 | EuroQol five dimension five level pain discomfort score |
| N062z | Allergic arthritis NOS |
| F373. | Polyneuropathy in malignant disease |
| N051F | Localised, primary osteoarthritis of elbow |
| N0400 | Rheumatoid arthritis of cervical spine |
| Fyu5B | [X]Drug-induced headache, not elsewhere classified |
| N03xD | Arthritis associated with other disease, ankle |
| L1642 | Peripheral neuritis in pregnancy - delivered with p/n comp |
| Fyu74 | [X]Polyneuropathy in neoplastic disease CE |
| N051D | Localised, primary osteoarthritis of the wrist |
| E0111 | Korsakov's alcoholic psychosis with peripheral neuritis |
| N0237 | Gouty arthritis of the ankle and foot |
| F371z | Polyneuropathy in collagen vascular disease NOS |
| N0531 | Localised osteoarthritis, unspecified, of shoulder region |
| N010y | Pyogenic arthritis of other specified sites |
| N0639 | Climacteric arthritis of multiple sites |
| FyuJ1 | [X]Retrobulbar neuritis in diseases classified elsewhere |
| Eu454 | [X]Persistent somatoform pain disorder |
| N03xC | Arthritis associated with other disease, tibio-fibular joint |
| N01zE | Infective arthritis NOS, of MCP joint |
| 38Vs. | Rheumatoid Arthritis Impact of Disease questionnaire |
| N040G | Rheumatoid arthritis of subtalar joint |
| N241z | Myalgia or myositis NOS |
| N040P | Seronegative rheumatoid arthritis |
| N01z4 | Infective arthritis NOS, of the hand |
| N05z6 | Osteoarthritis NOS, of the lower leg |
| 7P212 | Delivery of rehabilitation for pain syndromes |
| R090y | [D]Other specified abdominal pain |
| N05zK | Osteoarthritis NOS, of sacro-iliac joint |
| N0548 | Oligoarticular osteoarthritis, unspecified, other spec sites |
| L1640 | Peripheral neuritis in pregnancy unspecified |
| N01zT | Infective arthritis NOS, of IP joint of toe |
| J0464 | Temporomandibular joint-pain-dysfunction syndrome |
| N242. | Neuralgia, neuritis and radiculitis unspecified |
| Nyu1G | [X]Seropositive rheumatoid arthritis, unspecified |
| N023x | Gouty arthritis of multiple sites |
| N040E | Rheumatoid arthritis of tibio-fibular joint |
| N0521 | Localised, secondary osteoarthritis of the shoulder region |
| N04X. | Seropositive rheumatoid arthritis, unspecified |
| N05zE | Osteoarthritis NOS, of wrist |
| SP073 | Internal prosthetic device causing pain |
| N01zK | Infective arthritis NOS, of knee |
| C373K | Familial amyloid polyneuropathy, Iowa type |
| 1M02. | Shoulder joint painful on movement |
| N05z2 | Osteoarthritis NOS, of the upper arm |
| N0523 | Localised, secondary osteoarthritis of the forearm |
| N0236 | Gouty arthritis of the lower leg |
| N0513 | Localised, primary osteoarthritis of the forearm |
| SP07C | Pain due to shoulder joint prosthesis |
| N01z6 | Infective arthritis NOS, of the lower leg |
| N0546 | Oligoarticular osteoarthritis, unspecified, of lower leg |
| 9mM0. | Rheumatoid arthritis monitoring invitation first letter |
| L164z | Peripheral neuritis in pregnancy NOS |
| N0530 | Localised osteoarthritis, unspecified, of unspecified site |
| F35z. | Mononeuritis of unspecified site NOS |
| 38DZ. | Disease activity score in rheumatoid arthritis |
| N0409 | Rheumatoid arthritis of PIP joint of finger |
| N010x | Pyogenic arthritis of multiple sites |
| 43c6. | Rheumatoid arthritis screening test |
| ZV134 | [V]Personal history of arthritis |
| N0401 | Other rheumatoid arthritis of spine |
| N0518 | Localised, primary osteoarthritis of other specified site |
| Fyu5A | [X]Vascular headache, not elsewhere classified |
| F3745 | Polyneuropathy in hypoglycaemia |
| N0623 | Allergic arthritis of the forearm |
| N05zJ | Osteoarthritis NOS, of hip |
| N0626 | Allergic arthritis of the lower leg |
| N051. | Localised, primary osteoarthritis |
| 66HB0 | Rheumatoid arthritis annual review |
| N0541 | Oligoarticular osteoarthritis, unspecified, of shoulder |
| 1M8.. | Diabetic peripheral neuropathic pain |
| N0108 | Staphylococcal arthritis and polyarthritis |
| 43b9. | Rheumatoid arthritis particle agglutination test |
| F3749 | Polyneuropathy in sarcoidosis |
| N0665 | Unspecified monoarthritis of the pelvic region and thigh |
| N023y | Gouty arthritis of other specified site |
| N01zD | Infective arthritis NOS, of wrist |
| N0231 | Gouty arthritis of the shoulder region |
| Nyu16 | [X]Juvenile arthritis in other diseases CE |
| N0233 | Gouty arthritis of the forearm |
| N0404 | Rheumatoid arthritis of acromioclavicular joint |
| N0516 | Localised, primary osteoarthritis of the lower leg |
| C10EC | Type 1 diabetes mellitus with polyneuropathy |
| N0234 | Gouty arthritis of the hand |
| N05zB | Osteoarthritis NOS, of acromioclavicular joint |
| F3450 | Diabetic mononeuritis multiplex |
| N043. | Juvenile rheumatoid arthritis - Still's disease |
| Fyu76 | [X]Polyneuropathy in nutritional deficiency CE |
| N01z8 | Infective arthritis NOS, of shoulder |
| F347. | Complex regional pain syndrome type II |
| 38VH. | PSEQ - Pain Self-Efficacy Questionnaire |
| N05zU | Osteoarthritis NOS, of IP joint of toe |
| N0534 | Localised osteoarthritis, unspecified, of the hand |
| F3744 | Polyneuropathy in herpes zoster |
| N0625 | Allergic arthritis of the pelvic region and thigh |
| 9hQ1. | Excepted osteoarthritis quality indicators: informed dissent |
| N05z5 | Osteoarthritis NOS, pelvic region/thigh |
| N03xH | Arthritis associated with other disease, 1st MTP joint |
| N03x7 | Arthritis associated with other disease, PIP joint of finger |
| N03xA | Arthritis associated with other disease, sacro-iliac joint |
| N05zQ | Osteoarthritis NOS, of talonavicular joint |
| L1644 | Peripheral neuritis in pregnancy with postnatal complication |
| N0406 | Rheumatoid arthritis of distal radio-ulnar joint |
| N0620 | Allergic arthritis of unspecified site |
| F262A | Frequent episodic tension-type headache |
| N0106 | Pyogenic arthritis of the lower leg |
| F34.. | Mononeuritis of upper limb and mononeuritis multiplex |
| N0527 | Localised, secondary osteoarthritis of the ankle and foot |
| N0630 | Climacteric arthritis of unspecified site |
| N051G | Osteoarthritis of spinal facet joint |
| N0540 | Oligoarticular osteoarthritis, unspec, of unspecified sites |
| N0633 | Climacteric arthritis of the forearm |
| N0637 | Climacteric arthritis of the ankle and foot |
| F2629 | Infrequent episodic tension-type headache |
| N05zP | Osteoarthritis NOS, of subtalar joint |
| Fyu7B | [X]Inflammatory polyneuropathy, unspecified |
| C10FB | Type 2 diabetes mellitus with polyneuropathy |
| N0627 | Allergic arthritis of the ankle and foot |
| N01zQ | Infective arthritis NOS, of other tarsal joint |
| N05z7 | Osteoarthritis NOS, of ankle and foot |
| N0456 | Pauciarticular onset juvenile chronic arthritis |
| N0502 | Generalised osteoarthritis of multiple sites |
| R090P | [D]Functional abdominal pain syndrome |
| F370z | Acute infective polyneuritis NOS |
| N0526 | Localised, secondary osteoarthritis of the lower leg |
| N053. | Localised osteoarthritis, unspecified |
| N05zT | Osteoarthritis NOS, of lesser MTP joint |
| N0667 | Unspecified monoarthritis of the ankle and foot |
| N01zy | Infective arthritis NOS, of other specified site |
| N05.. | Osteoarthritis and allied disorders |
| 9NNh. | Under care of pain management specialist |
| Ryu11 | [X]Other and unspecified abdominal pain |
| N0664 | Unspecified monoarthritis of the hand |
| N040L | Rheumatoid arthritis of lesser MTP joint |
| N05zS | Osteoarthritis NOS, of 1st MTP joint |
| N01zJ | Infective arthritis NOS, of sacro-iliac joint |
| 38VG. | PCS - Pain Catastrophising Scale |
| SP07A | Pain due to hip joint prosthesis |
| F3711 | Polyneuropathy in polyarteritis nodosa |
| N0549 | Oligoarticular osteoarthritis, unspecified, multiple sites |
| N0235 | Gouty arthritis of the pelvic region and thigh |
| C109B | Non-insulin dependent diabetes mellitus with polyneuropathy |
| N050. | Generalised osteoarthritis – OA |
| N1441 | Lumbosacral neuritis, unspecified |
| 14CH. | History of chronic pancreatitis |
| N040M | Rheumatoid arthritis of IP joint of toe |
| Fyu73 | [X]Polyneuropathy in infectious+parasitic diseases CE |
| A94y4 | Syphilitic retrobulbar neuritis |
| N0663 | Unspecified monoarthritis of the forearm |
| Nyu10 | [X]Rheumatoid arthritis+involvement/other organs or systems |
| N0638 | Climacteric arthritis of other specified site |
| N0636 | Climacteric arthritis of the lower leg |
| N03x3 | Arthritis associated with other disease, elbow |
| N052z | Localised, secondary osteoarthritis NOS |
| N0547 | Oligoarticular osteoarthritis, unspecified, of ankle/foot |
| F300. | Post-herpetic trigeminal neuralgia |
| N11D0 | Osteoarthritis of cervical spine |
| N0403 | Rheumatoid arthritis of sternoclavicular joint |
| N0101 | Pyogenic arthritis of the shoulder region |
| Fyu77 | [X]Polyneuropathy/systemic connective tissue disorders CE |
| N05zC | Osteoarthritis NOS, of elbow |
| N03x5 | Arthritis associated with other disease, wrist |
| Nyu12 | [X]Other specified rheumatoid arthritis |
| N241. | Myalgia and myositis unspecified |
| F35y. | Unspecified mononeuritis lower limb |
| N0453 | Juvenile arthritis in Crohn's disease |
| N054. | Oligoarticular osteoarthritis, unspecified |
| F3712 | Polyneuropathy in rheumatoid arthritis |
| N0451 | Juvenile seronegative polyarthritis |
| N0624 | Allergic arthritis of the hand |
| N0501 | Generalised osteoarthritis of the hand |
| N03x2 | Arthritis associated with other disease, acromioclavic joint |
| Fyu54 | [X]Other specified headache syndromes |
| N0635 | Climacteric arthritis of the pelvic region and thigh |
| N11D1 | Osteoarthritis of thoracic spine |
| F37X. | Inflammatory polyneuropathy, unspecified |
| N01zx | Infective arthritis NOS, of multiple sites |
| N0500 | Generalised osteoarthritis of unspecified site |
| N144. | Thoracic and lumbosacral neuritis |
| N0621 | Allergic arthritis of the shoulder region |
| N065. | Unspecified polyarthropathy or polyarthritis |
| N0533 | Localised osteoarthritis, unspecified, of the forearm |
| N33C. | Complex regional pain syndrome type I |
| N0536 | Localised osteoarthritis, unspecified, of the lower leg |
| N0107 | Pyogenic arthritis of the ankle and foot |
| F3721 | Chronic painful diabetic neuropathy |
| Nyu11 | [X]Other seropositive rheumatoid arthritis |
| N0661 | Unspecified monoarthritis of the shoulder region |
| N0660 | Unspecified monoarthritis of unspecified site |
| N01z1 | Infective arthritis NOS, of the shoulder region |
| N01zH | Infective arthritis NOS, of hip |
| 38Vk. | Arthritis Research UK musculoskeletal health questionnaire |
| NyuA3 | [X]Other soft tissue disorders related/use,overuse+pressure |
| Fyu75 | [X]Polyneuropathy/other endocrine+metabolic diseases CE |
| Fyu7C | [X] Polyneuropathy, unspecified |
| N0515 | Localised, primary osteoarthritis of the pelvic region/thigh |
| N01zF | Infective arthritis NOS, of PIP joint of finger |
| N05zH | Osteoarthritis NOS, of DIP joint of finger |
| N01zM | Infective arthritis NOS, of ankle |
| N0109 | Pneumococcal arthritis and polyarthritis |
| 9Ok5. | Cancer pain and symptom management |
| N05z9 | Osteoarthritis NOS, of shoulder |
| N040C | Rheumatoid arthritis of sacro-iliac joint |
| N0537 | Localised osteoarthritis, unspecified, of the ankle and foot |
| Fyu79 | [X]Polyneuropathy in other diseases classified elsewhere |
| N05z0 | Osteoarthritis NOS, of unspecified site |
| Fyu78 | [X]Polyneuropathy/other musculoskeletal disorders CE |
| N144z | Thoracic and lumbosacral neuritis NOS |
| N0522 | Localised, secondary osteoarthritis of the upper arm |
| N043z | Juvenile rheumatoid arthritis NOS |
| N0622 | Allergic arthritis of the upper arm |
| N0632 | Climacteric arthritis of the upper arm |
| SP07B | Pain due to knee joint prosthesis |
| N0629 | Allergic arthritis of multiple sites |
| N04.. | Rheumatoid arthritis and other inflammatory polyarthropathy |
| N065z | Unspecified polyarthropathy or polyarthrititis NOS |
| N03x0 | Arthritis associated with other disease, shoulder |
| N05z3 | Osteoarthritis NOS, of the forearm |
| N0511 | Localised, primary osteoarthritis of the shoulder region |
| N03x6 | Arthritis associated with other disease, MCP joint |
| Nyu02 | [X]Arthritis in other infectious and parasitic diseases CE |
| N03xG | Arthritis associated with other disease, other tarsal joint |
| N023z | Gouty arthritis NOS |
| F3964 | Myopathy due to rheumatoid arthritis |
| N03xE | Arthritis associated with other disease, subtalar joint |
| N047. | Seropositive erosive rheumatoid arthritis |
| N053z | Localised osteoarthritis, unspecified, NOS |
| F3710 | Polyneuropathy in disseminated lupus erythematosus |
| N0538 | Localised osteoarthritis, unspecified, of other spec site |
| F2X.. | Vascular headache, not elsewhere classified |
| N01zG | Infective arthritis NOS, of DIP joint of finger |
| N01zA | Infective arthritis NOS, of acromioclavicular joint |
| L384. | Obstetric spinal and epidural anaesthesia-induced headache |
| N052. | Localised, secondary osteoarthritis |
| N0431 | Acute polyarticular juvenile rheumatoid arthritis |
| N040S | Rheumatoid arthritis - multiple joint |
| N040H | Rheumatoid arthritis of talonavicular joint |
| N0100 | Pyogenic arthritis of unspecified site |
| N0232 | Gouty arthritis of the upper arm |
| FyuU4 | [X]Acoustic neuritis in infectious+parasitic diseases CE |
| 1M111 | Ischaemic foot pain when walking |
| 38Va. | Psoriatic Arthritis Impact of Disease 9 questionnaire |
| R090E | [D]Recurrent acute abdominal pain |
| 8H4c. | Referral to headache special interest general practitioner |
| N01zL | Infective arthritis NOS, of tibio-fibular joint |
| F3742 | Polyneuropathy in vitamin B deficiency |
| N0528 | Localised, secondary osteoarthritis of other specified site |
| N0535 | Localised osteoarthritis, unspecified, pelvic region/thigh |
| 9hQ.. | Exception reporting: osteoarthritis quality indicators |
| 38DZ0 | Disease activity score 28 joint in rheumatoid arthritis |
| N0542 | Oligoarticular osteoarthritis, unspecified, of upper arm |
| F2W.. | Drug-induced headache, not elsewhere classified |
| N0517 | Localised, primary osteoarthritis of the ankle and foot |
| N0102 | Pyogenic arthritis of the upper arm |
| N0520 | Localised, secondary osteoarthritis of unspecified site |
| ZV177 | [V]Family history of arthritis |
| N0105 | Pyogenic arthritis of the pelvic region and thigh |
| N0402 | Rheumatoid arthritis of shoulder |
| N0666 | Unspecified monoarthritis of the lower leg |
| N01z9 | Infective arthritis NOS, of sternoclavicular joint |

## Appendix 2 ICD-11 and ICD-10 codes for persistent pain, developed through discussion with project team members and stakeholders

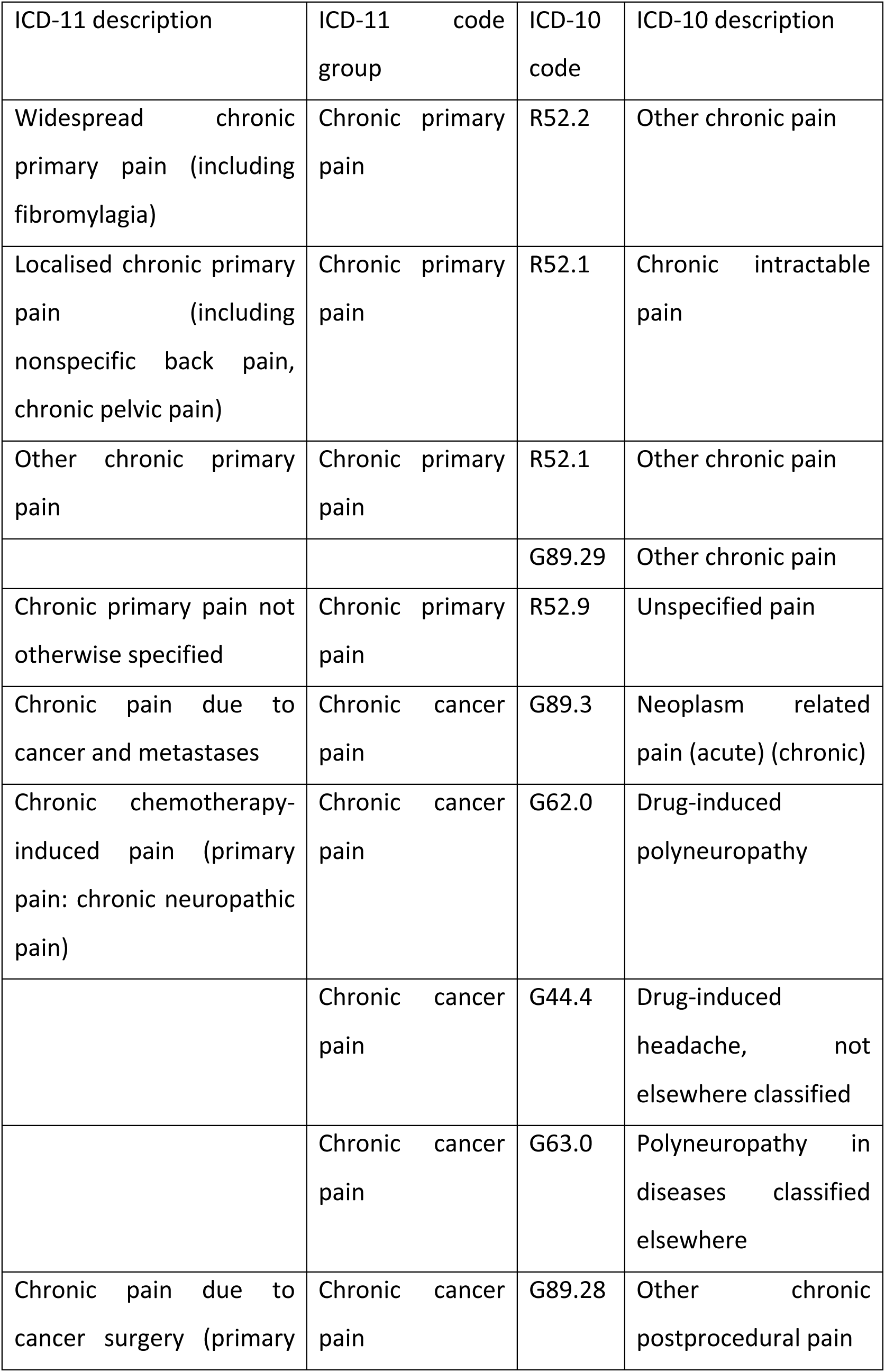
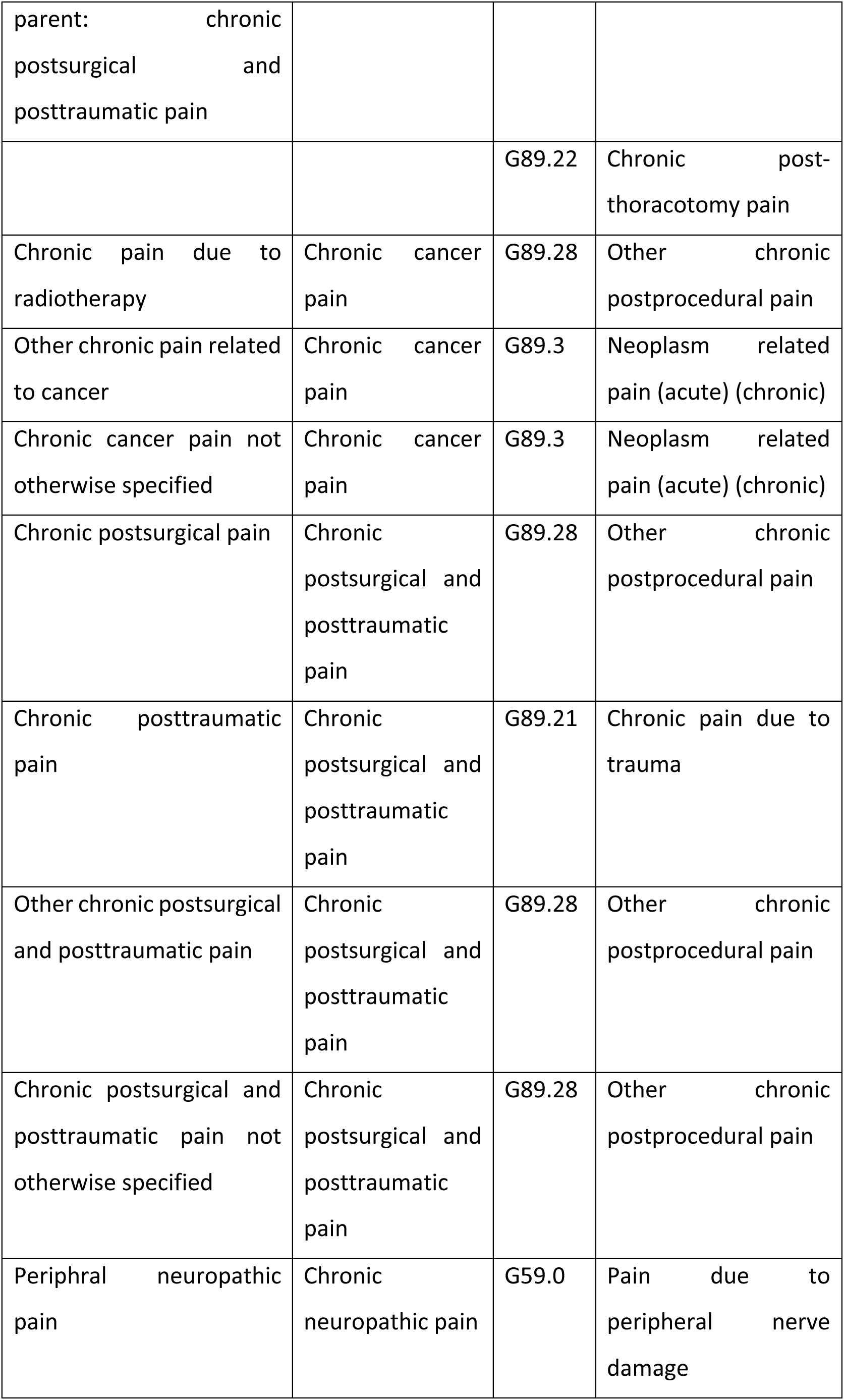
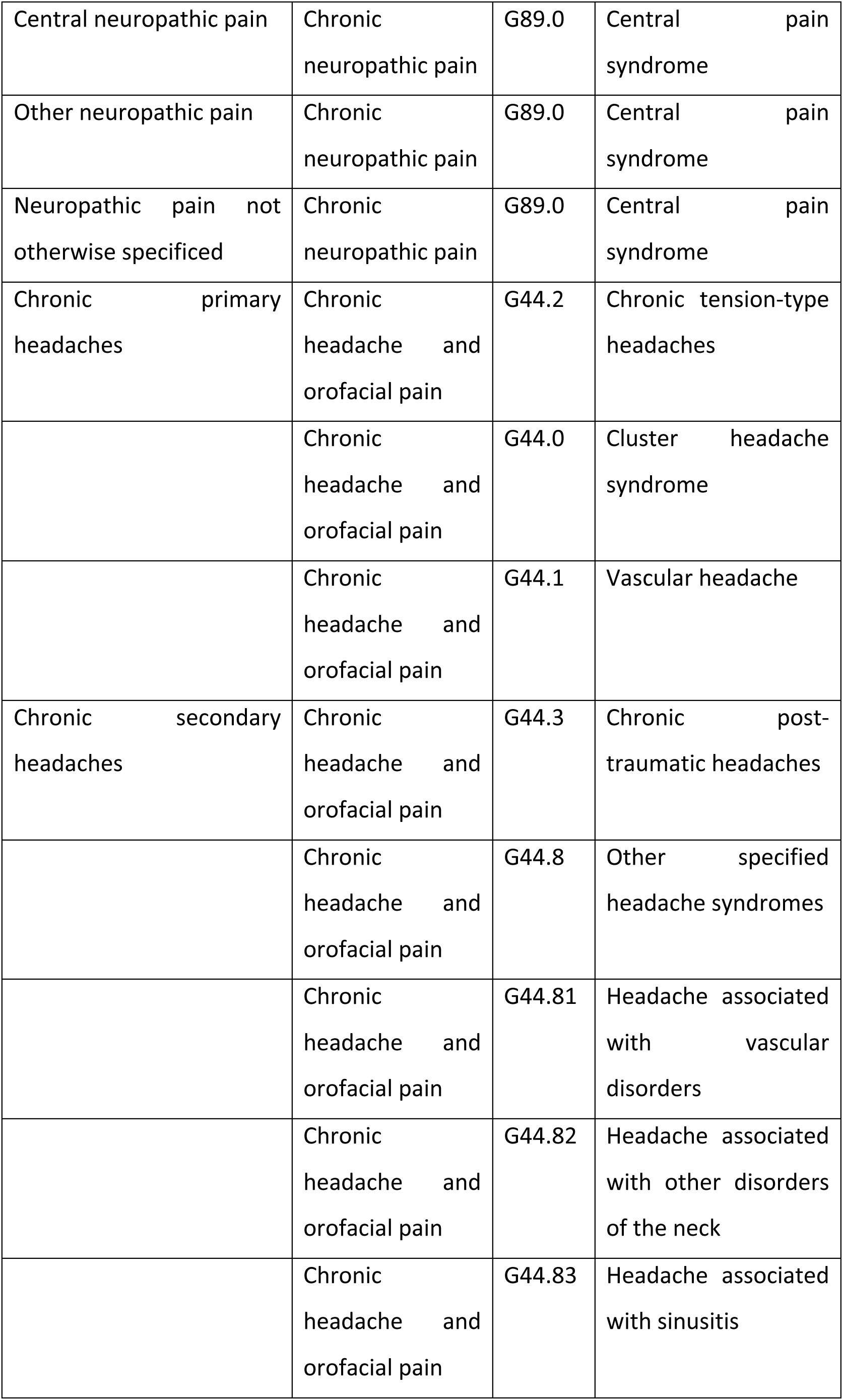
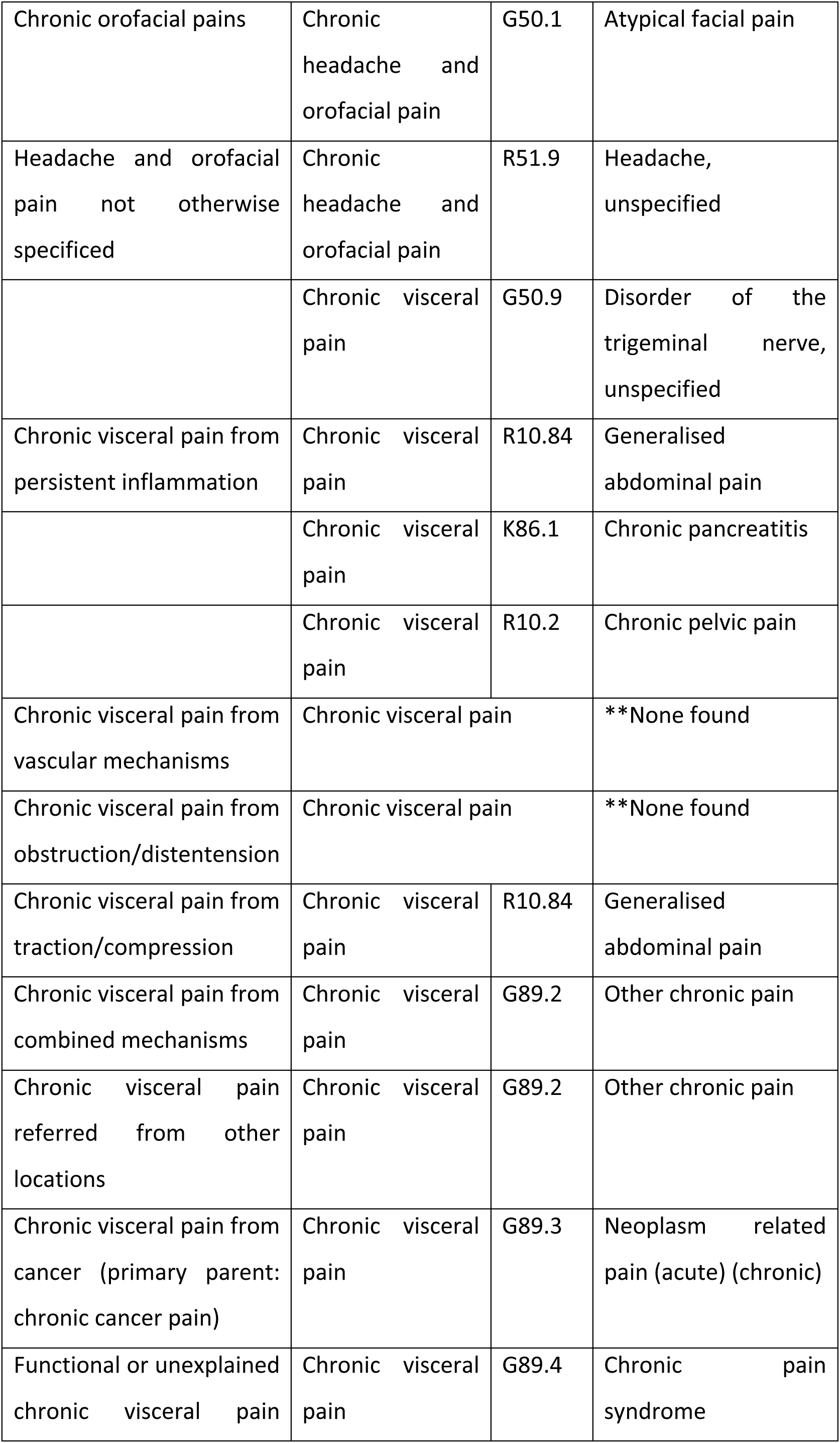
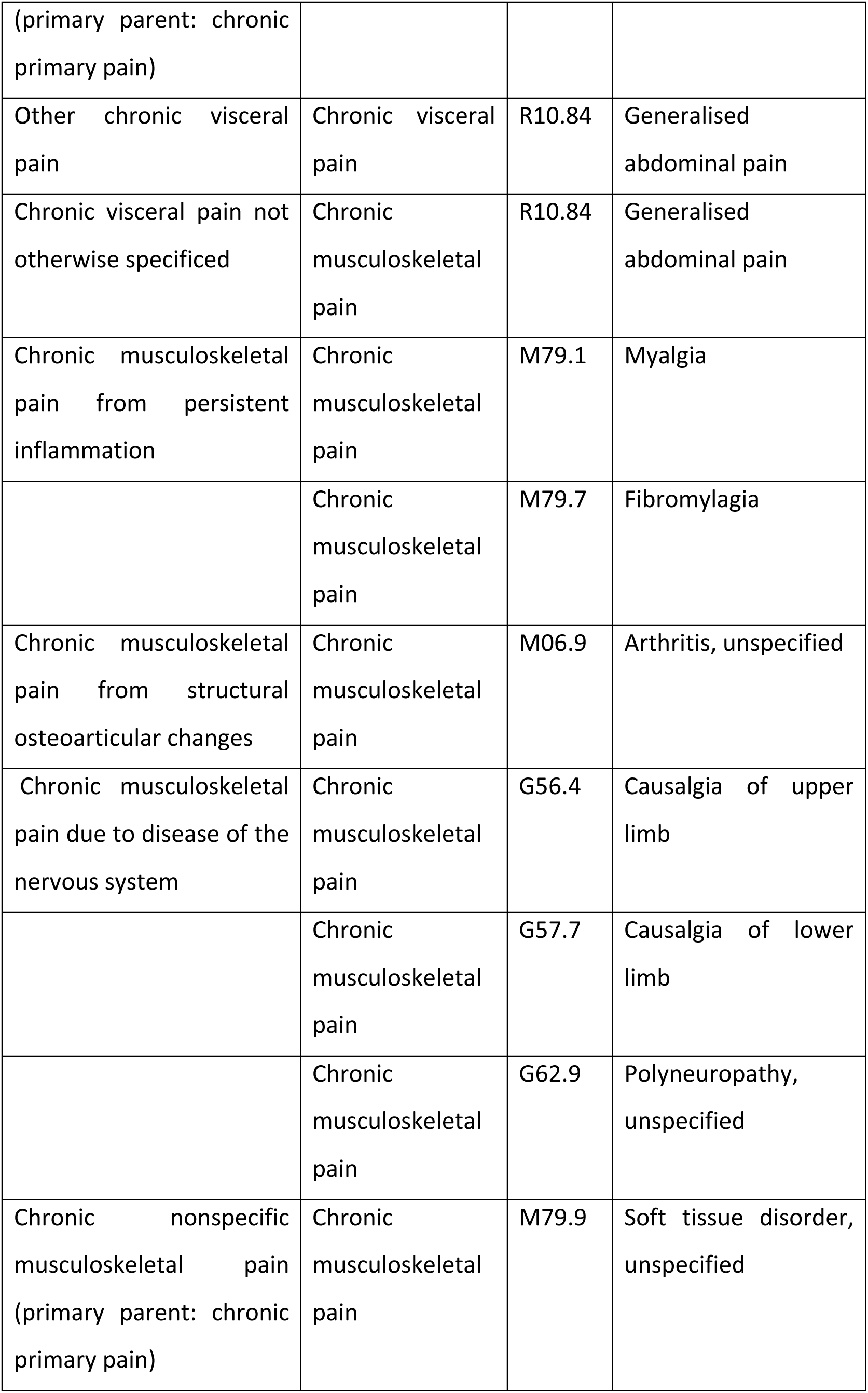
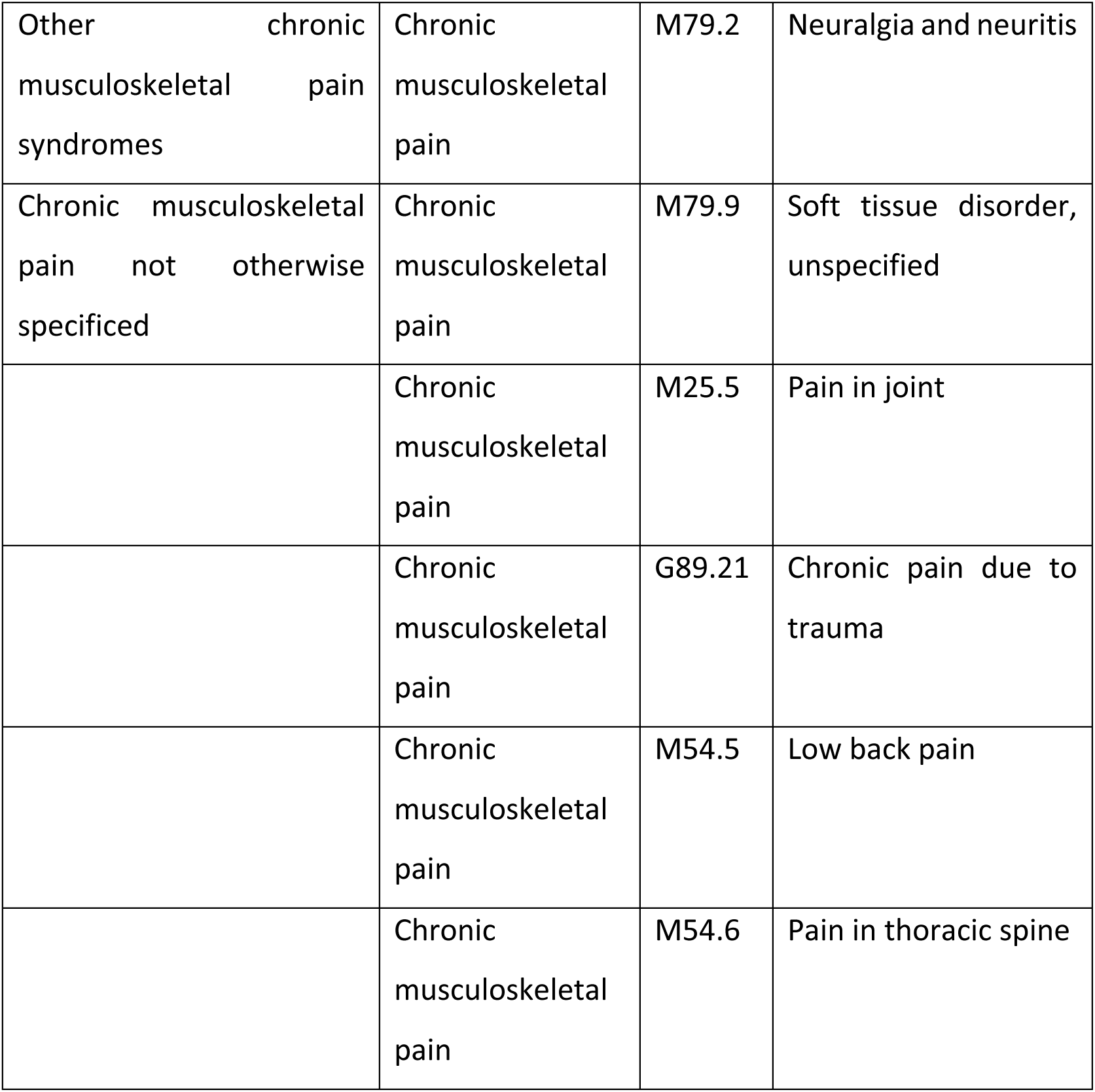

## Appendix 3 Medications commonly prescribed to individuals experiencing persistent pain, developed through discussion with project team members and stakeholders

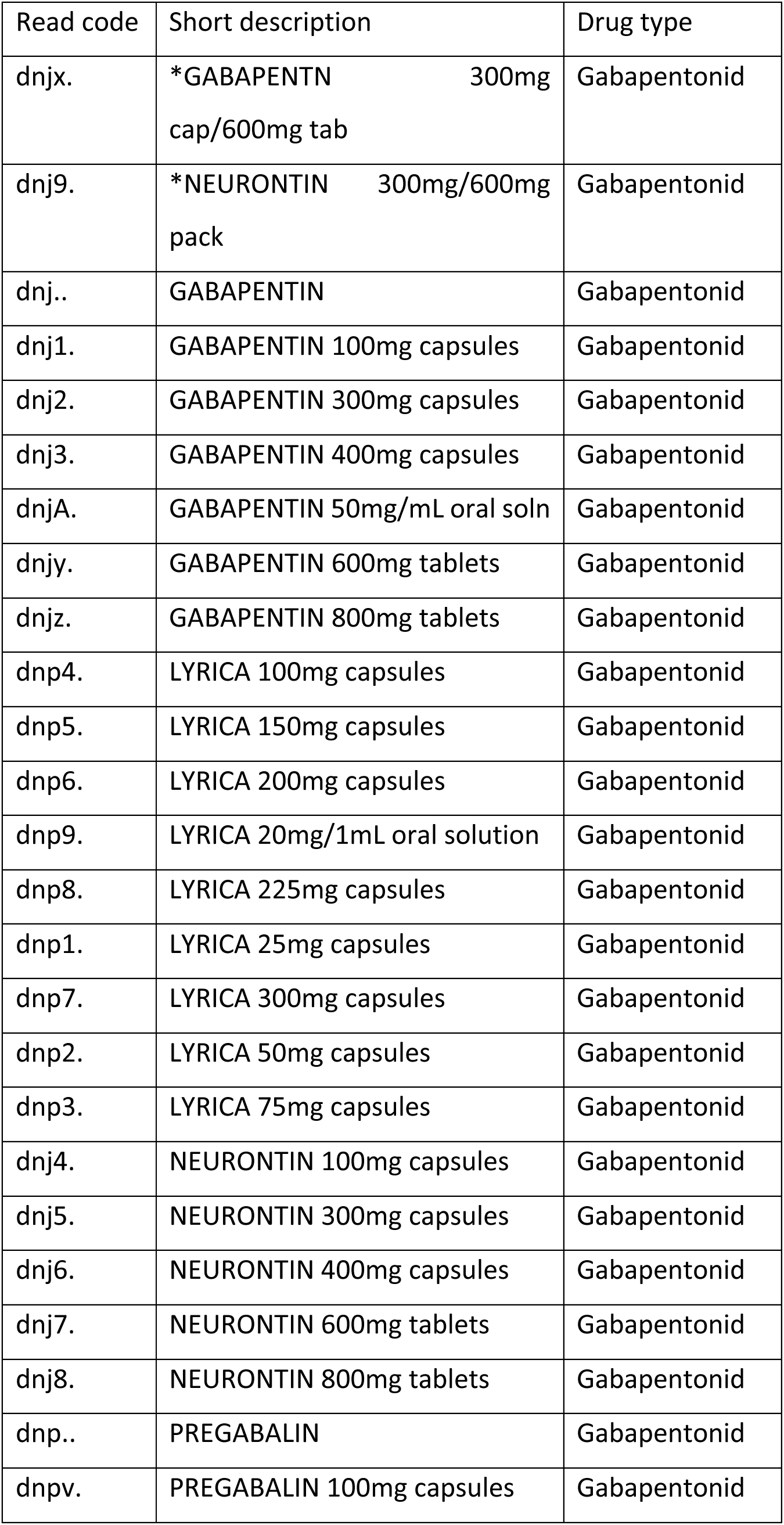
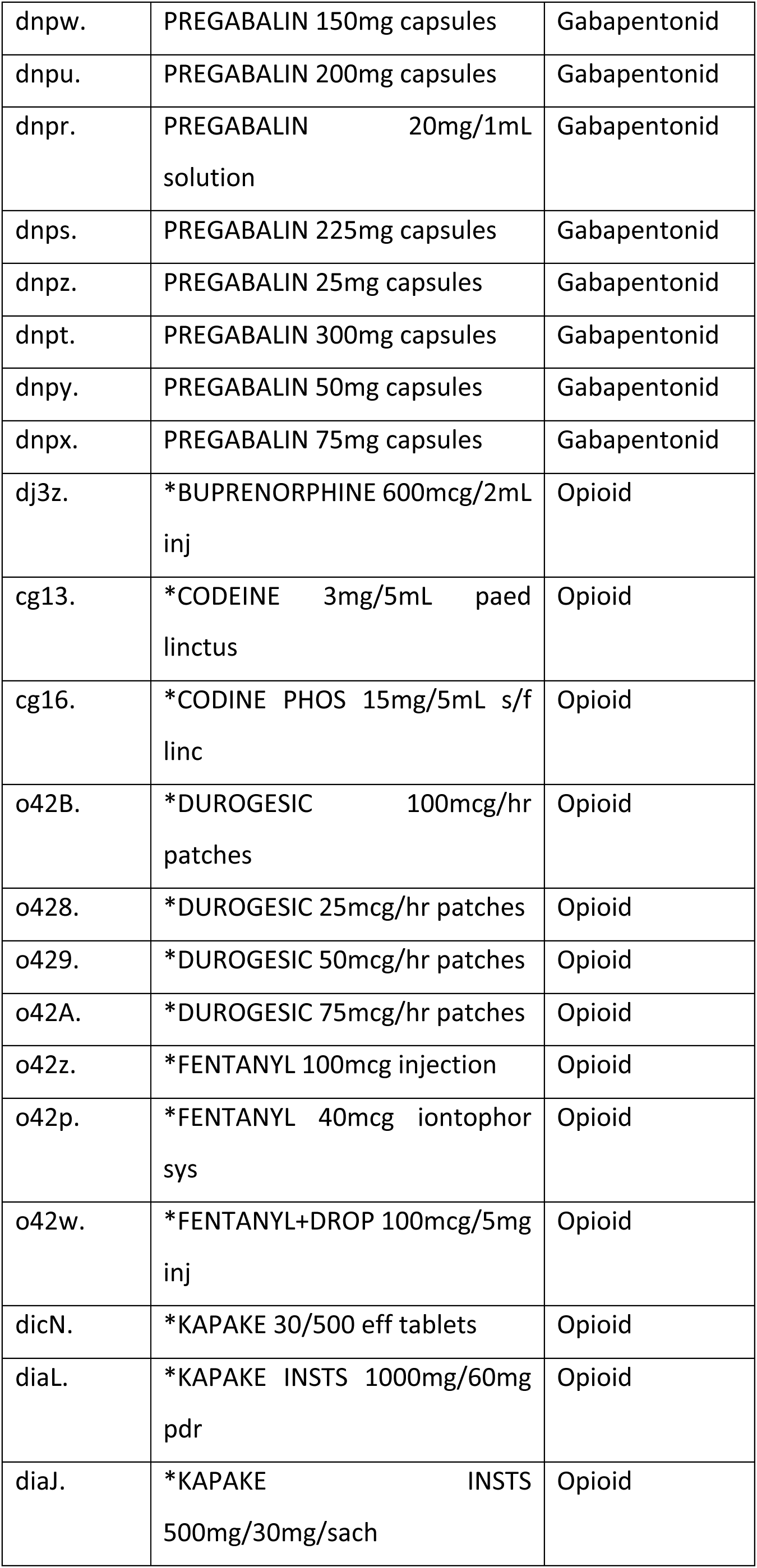
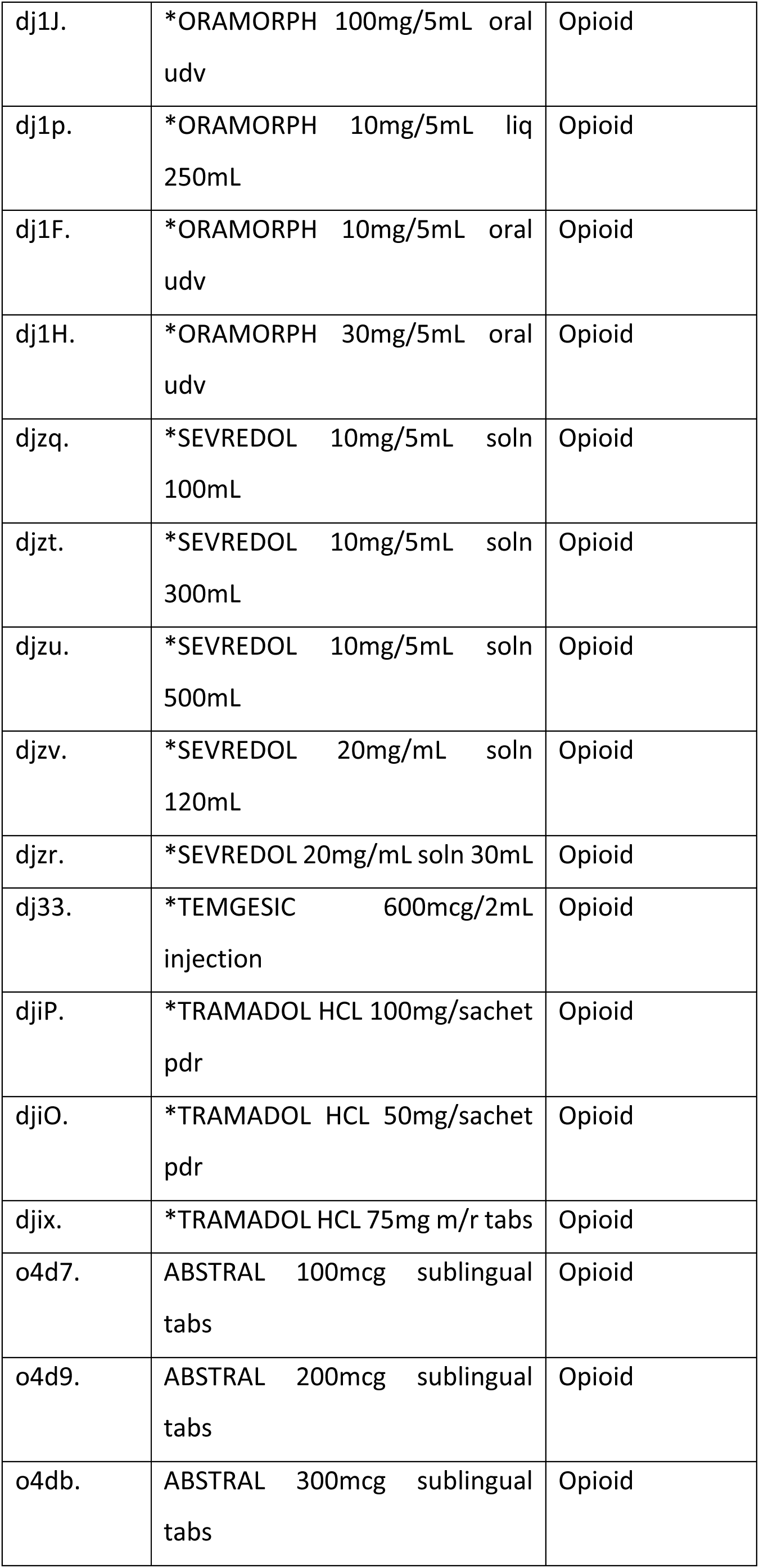
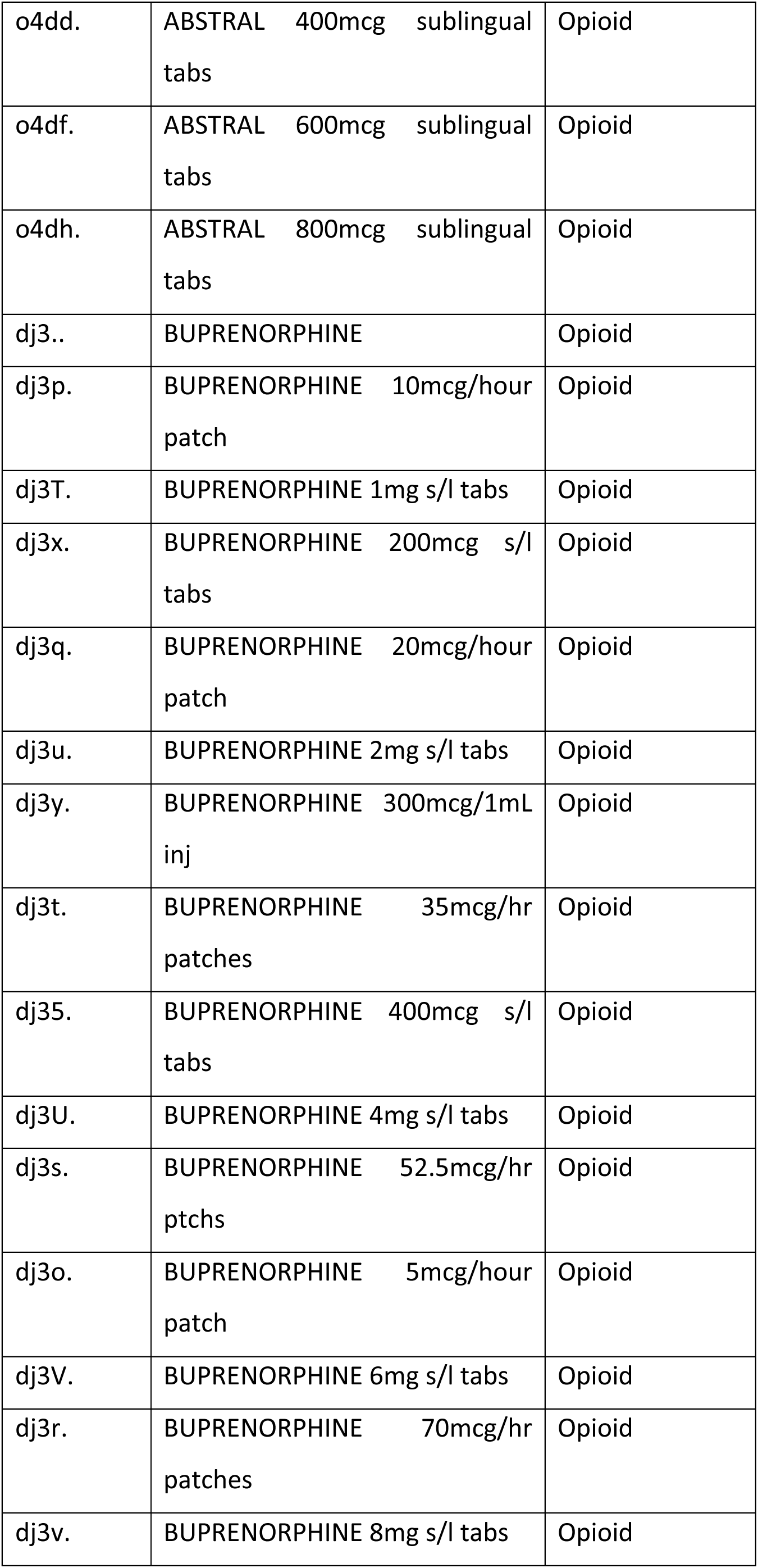
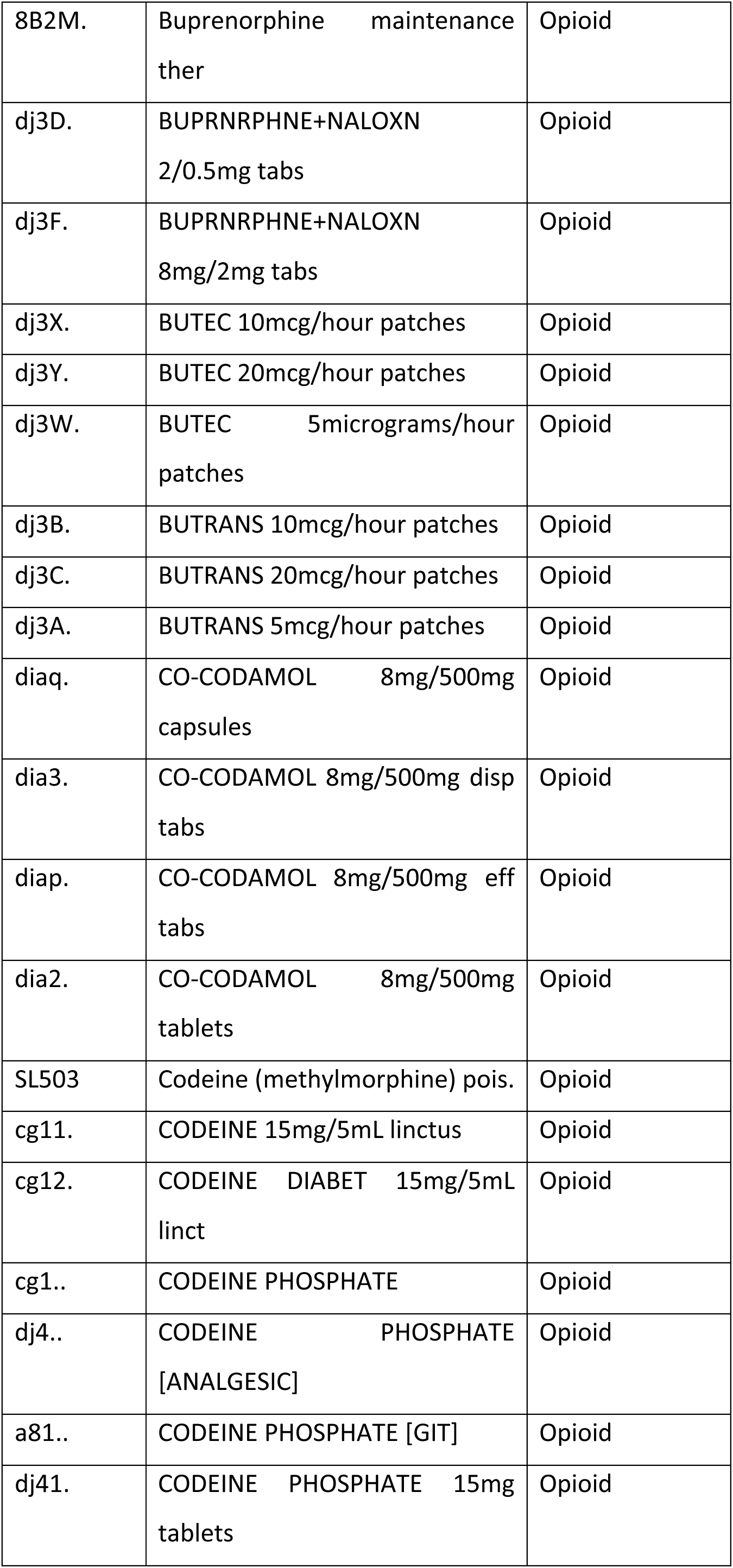
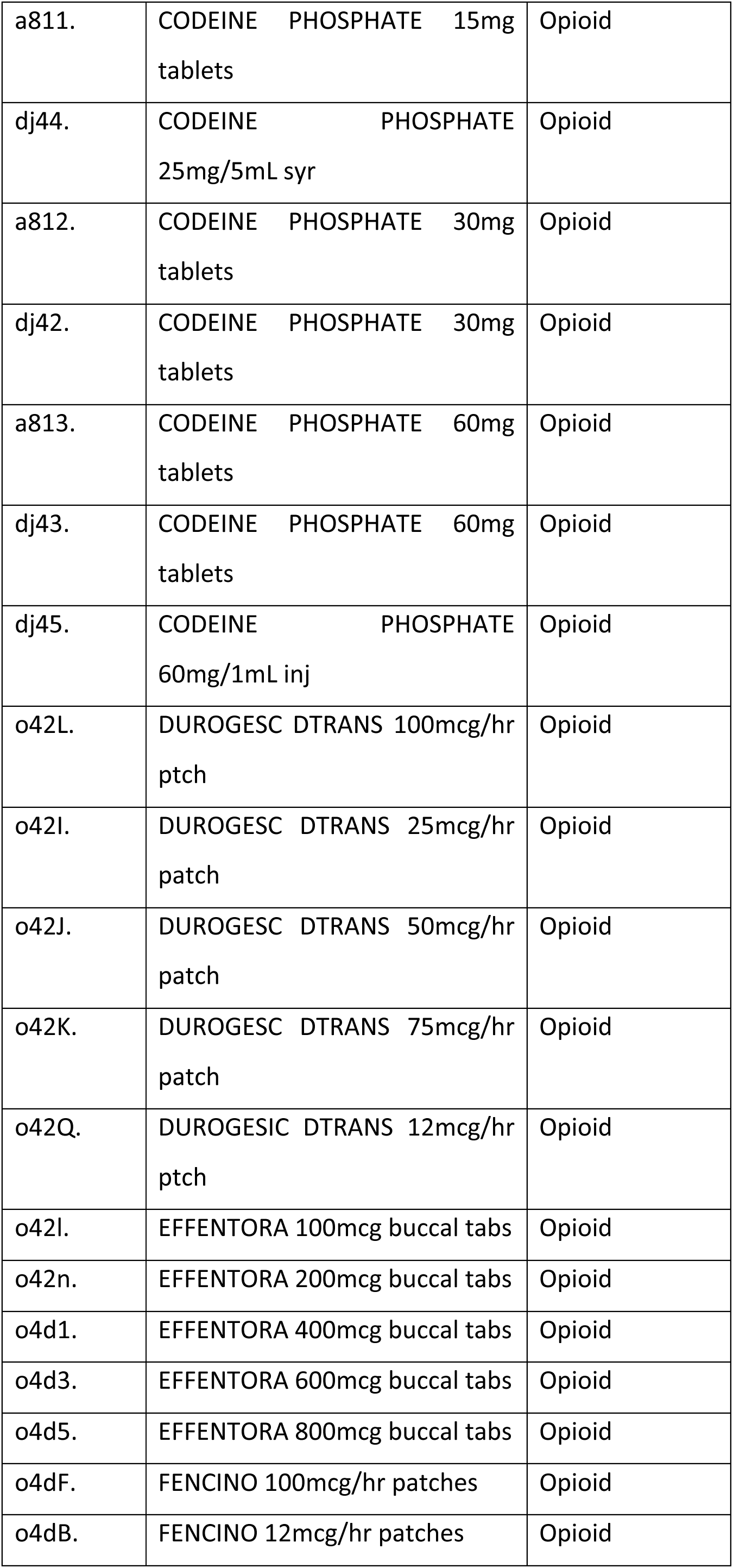
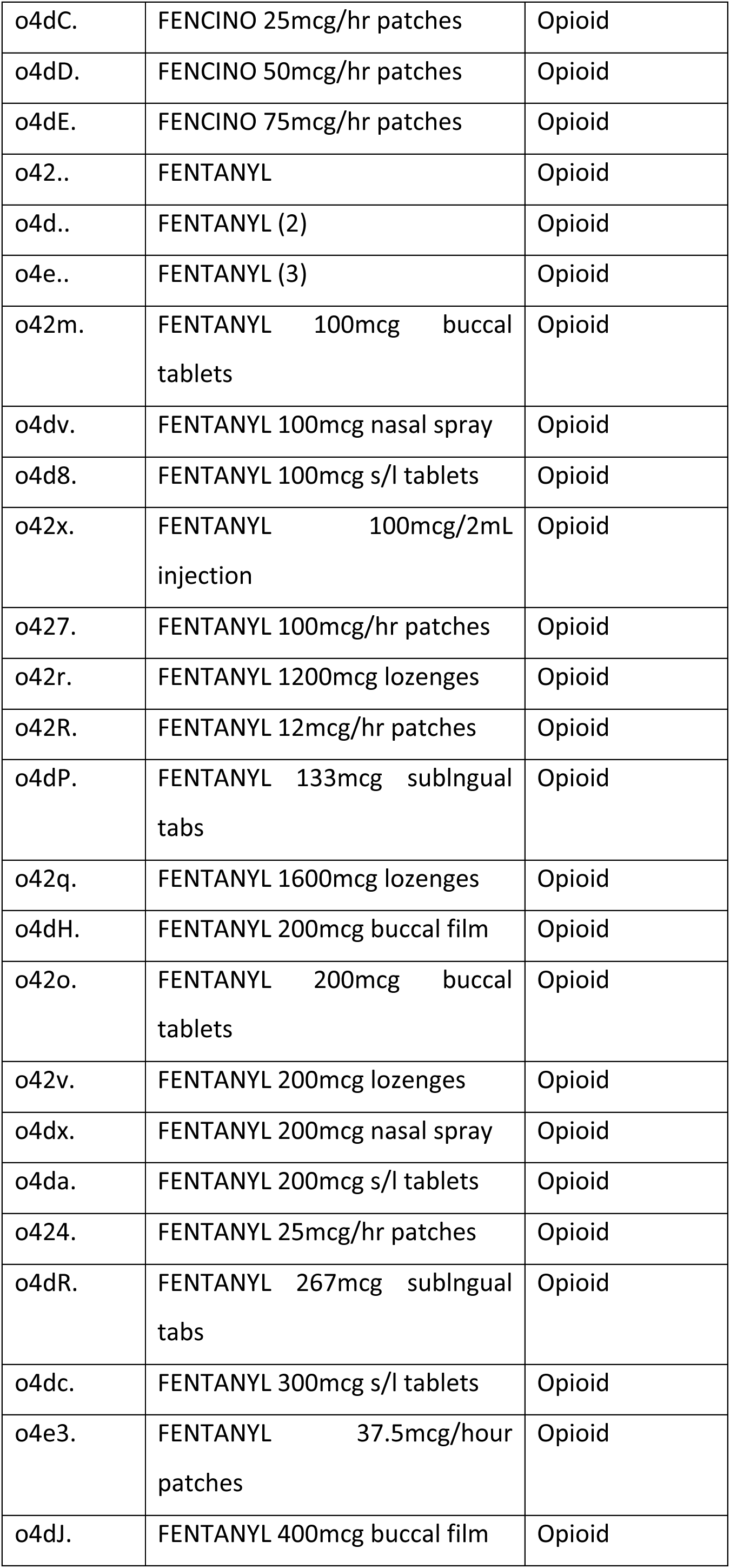
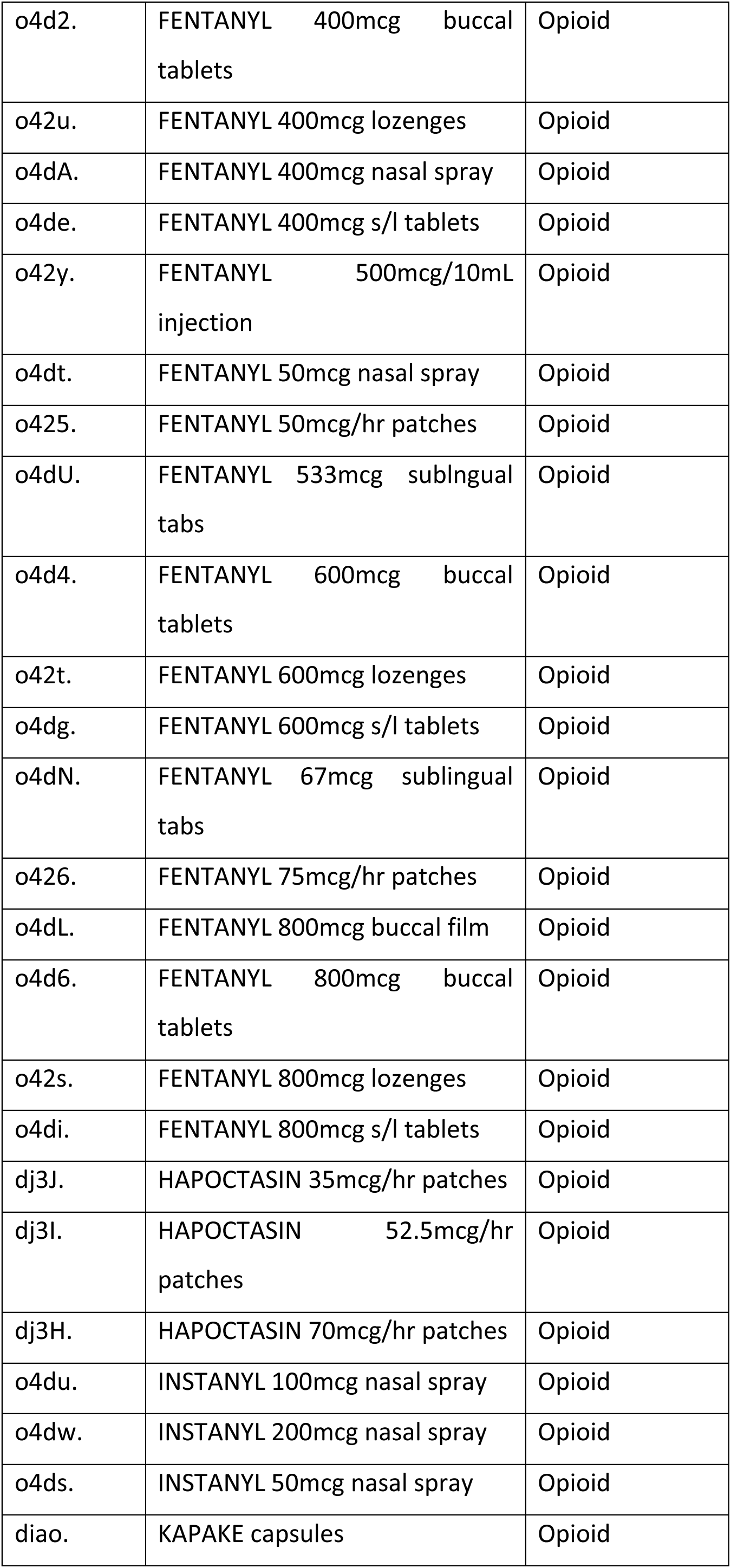
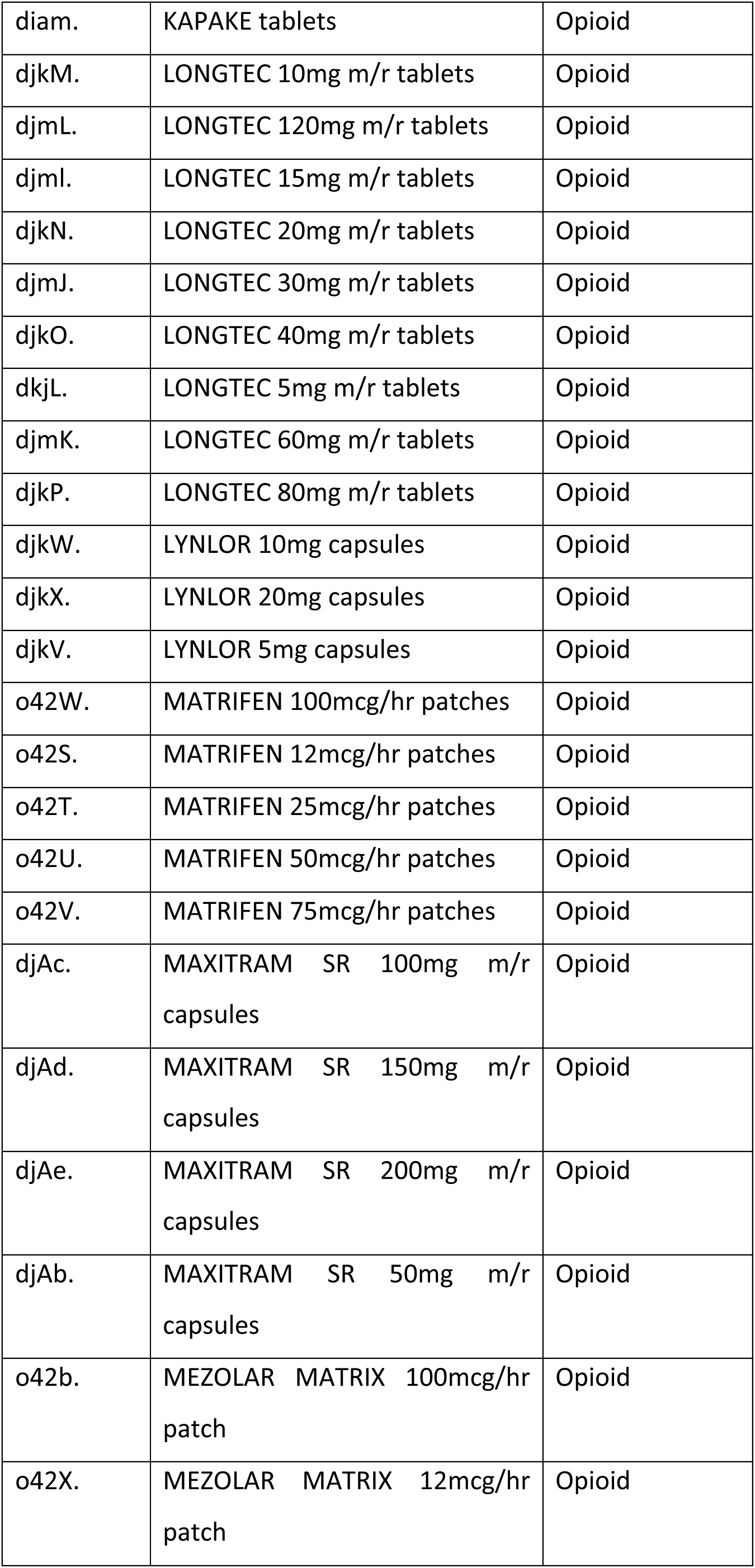
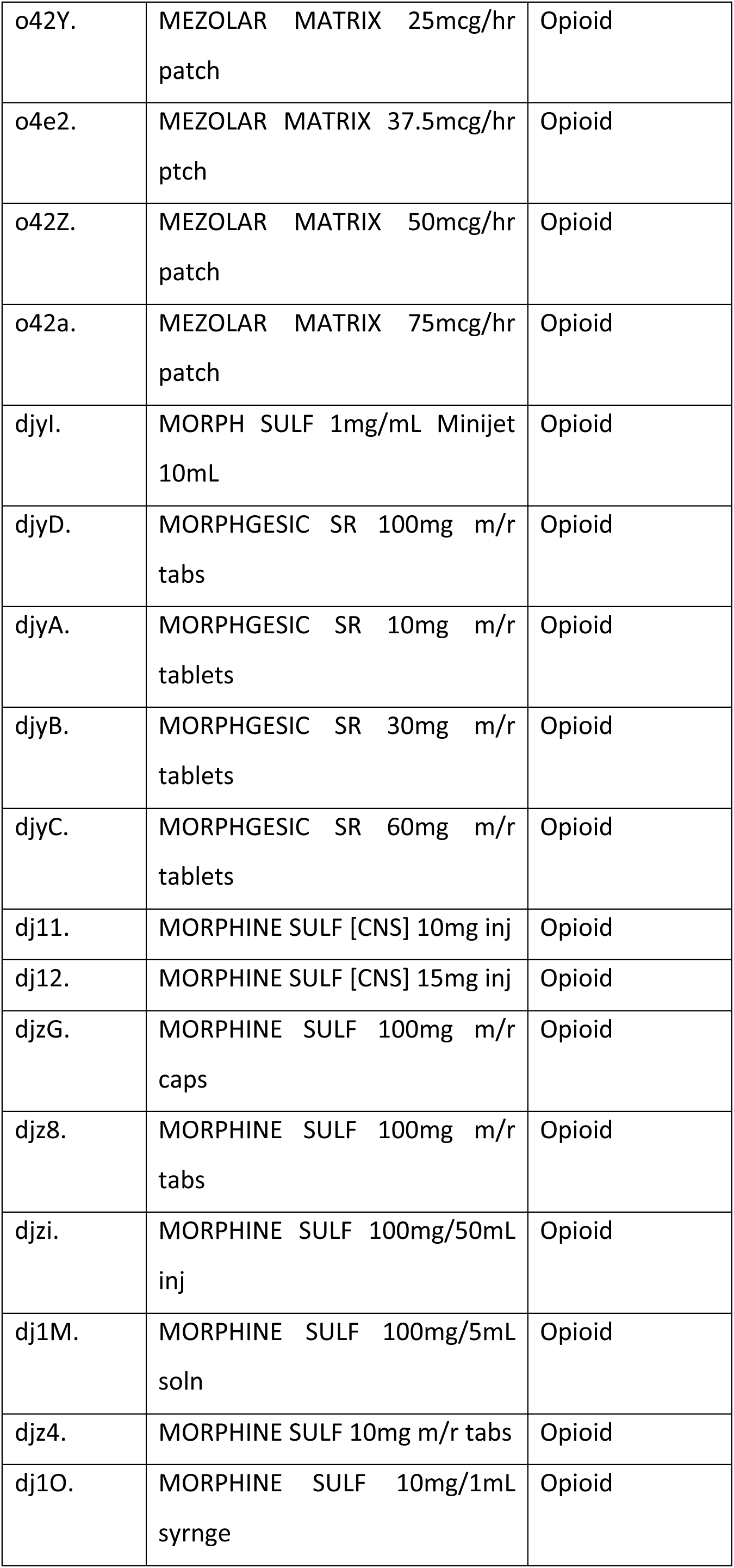
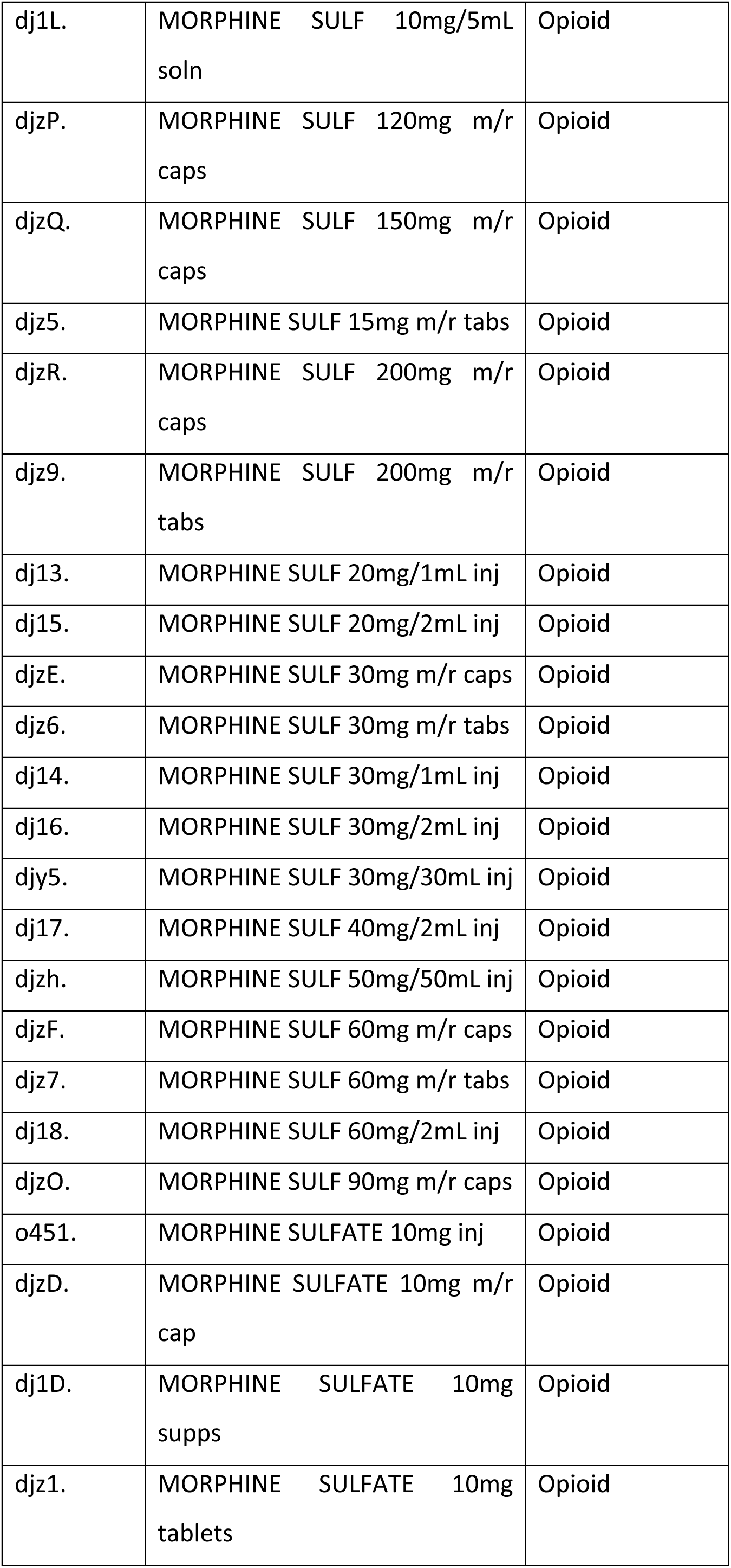
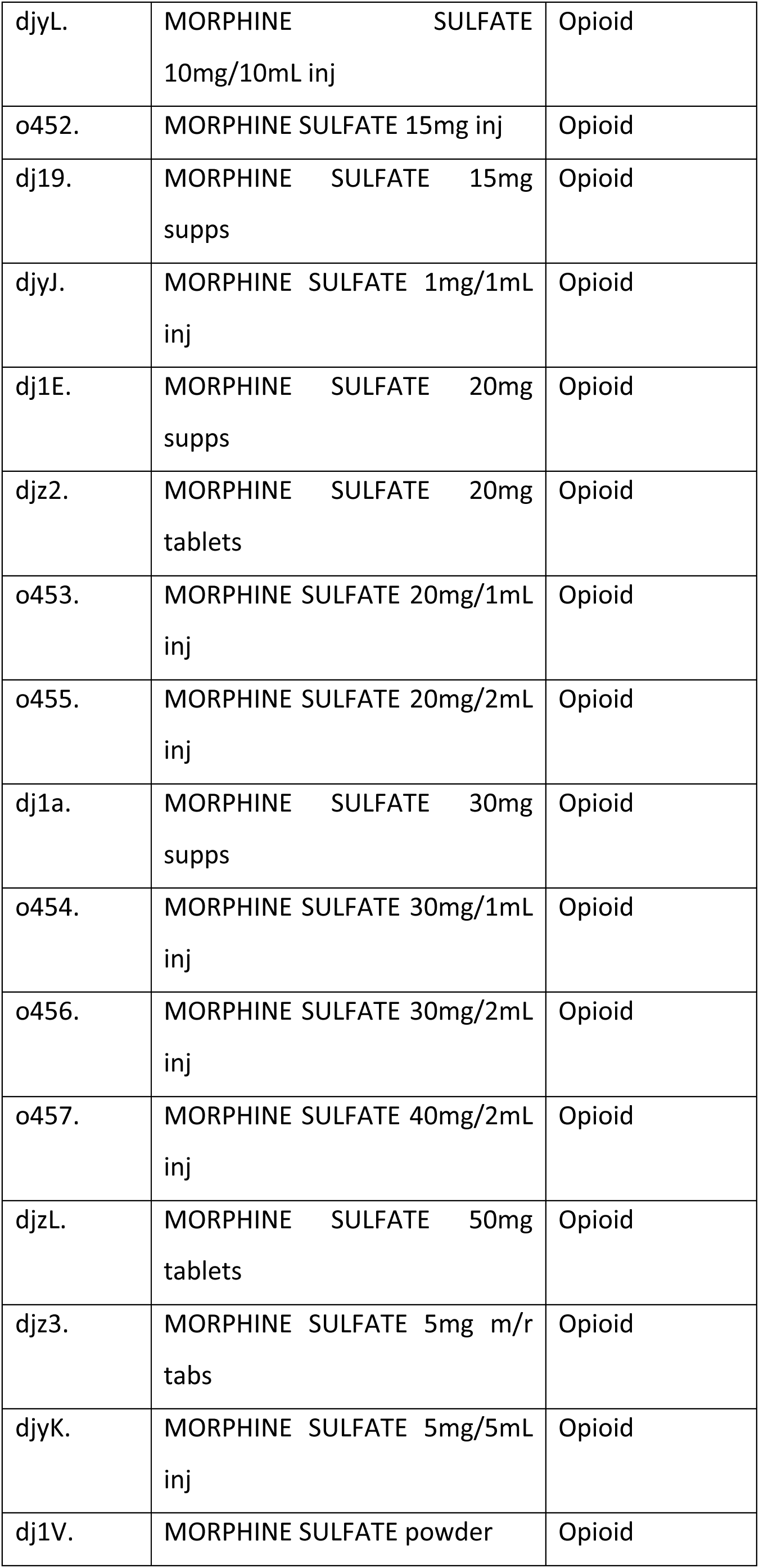
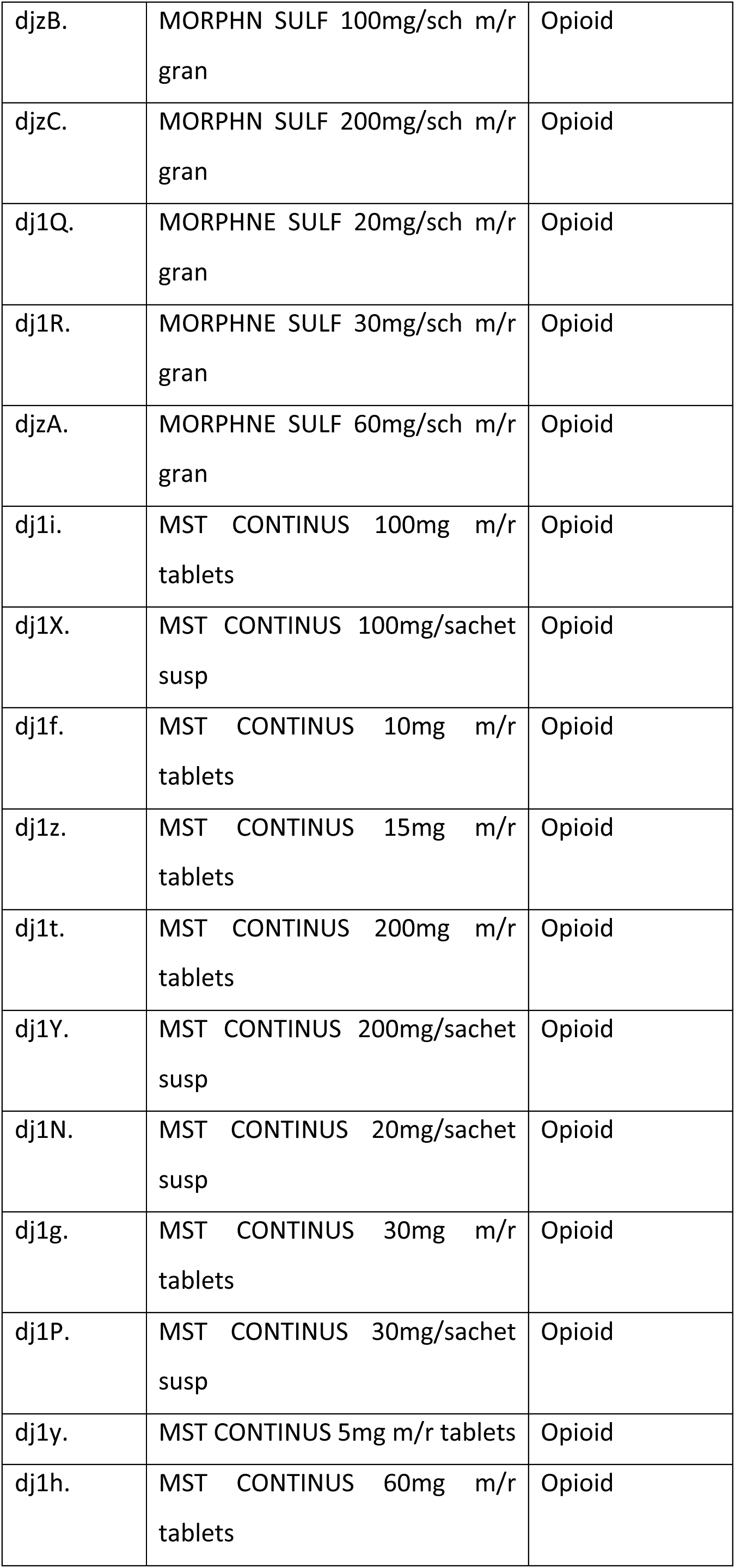
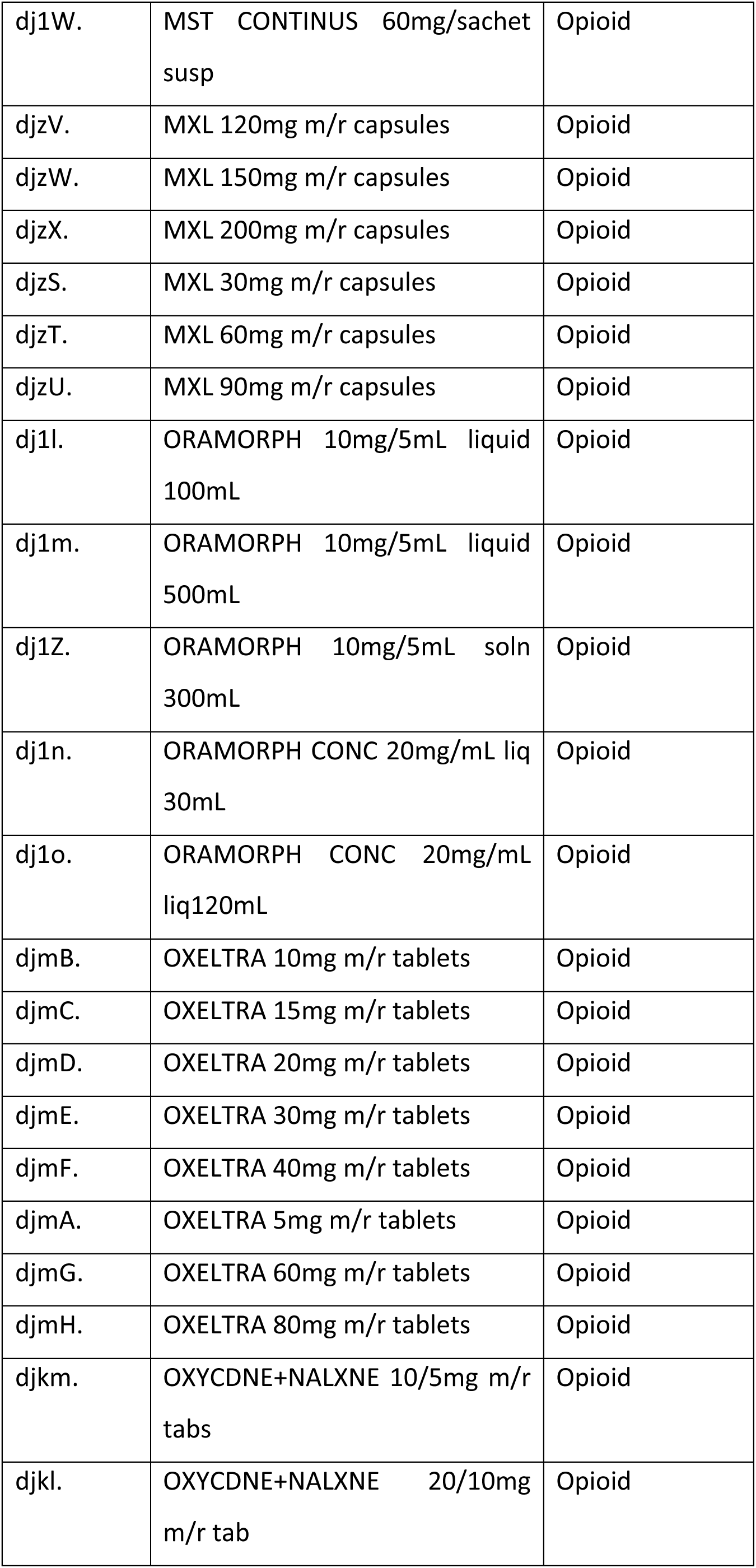
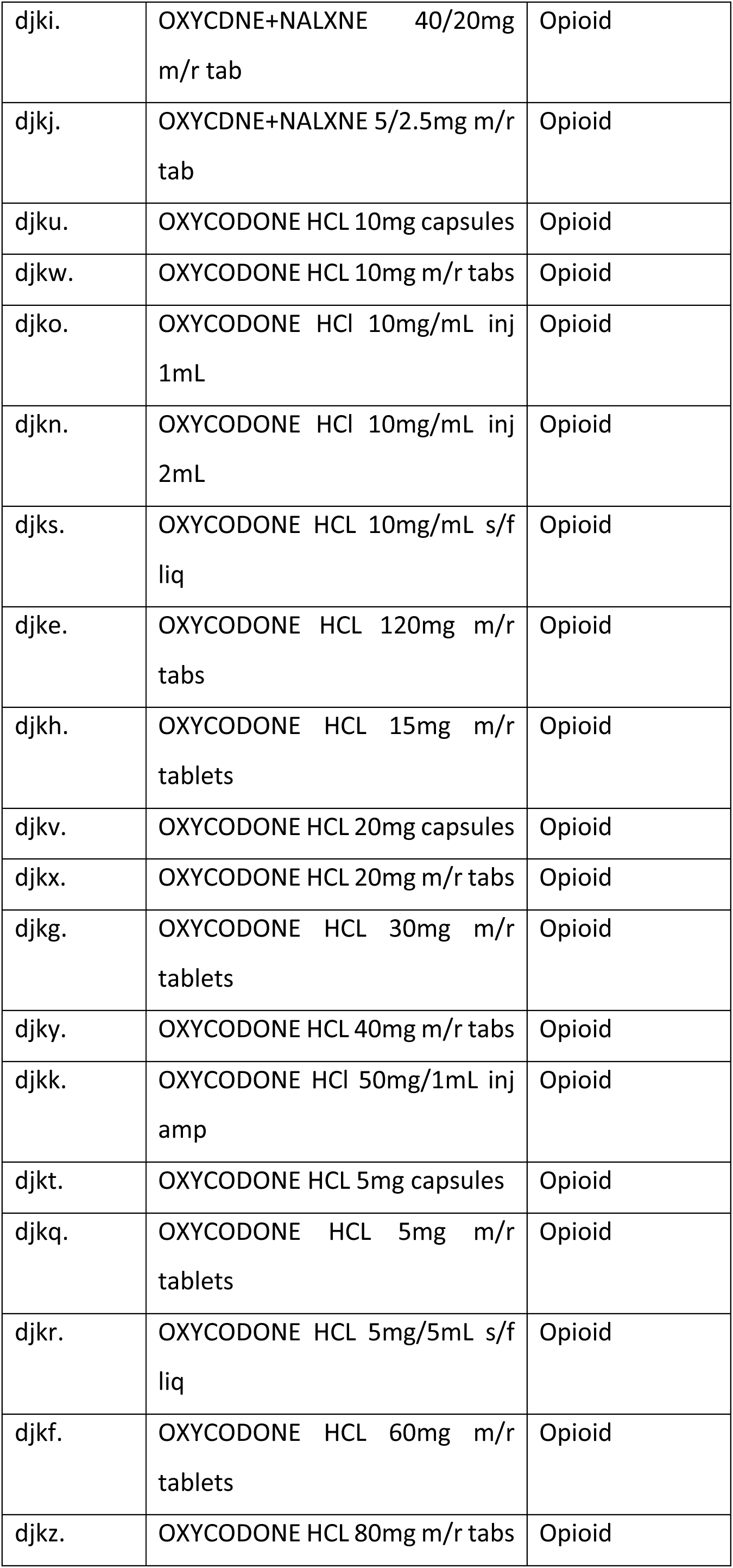
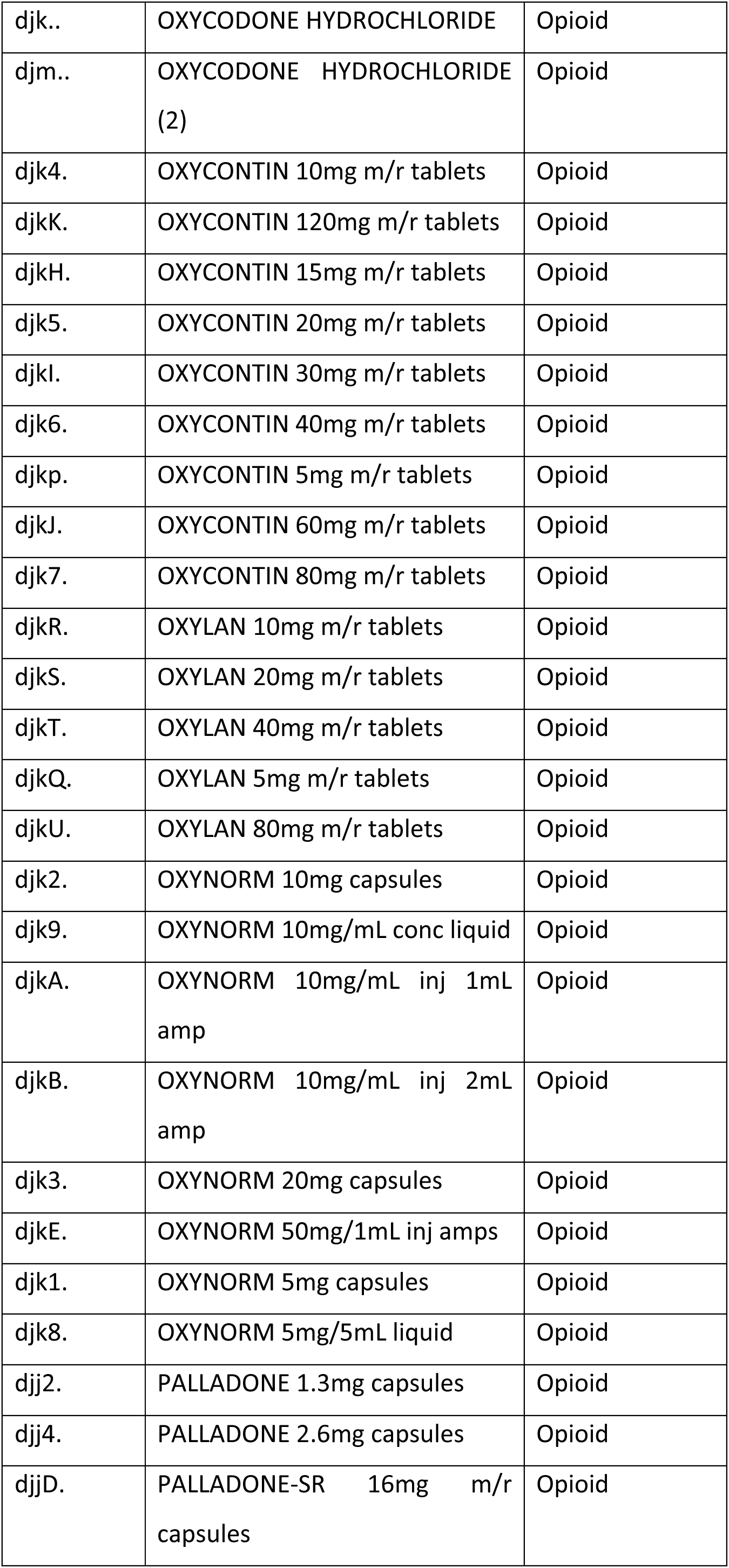
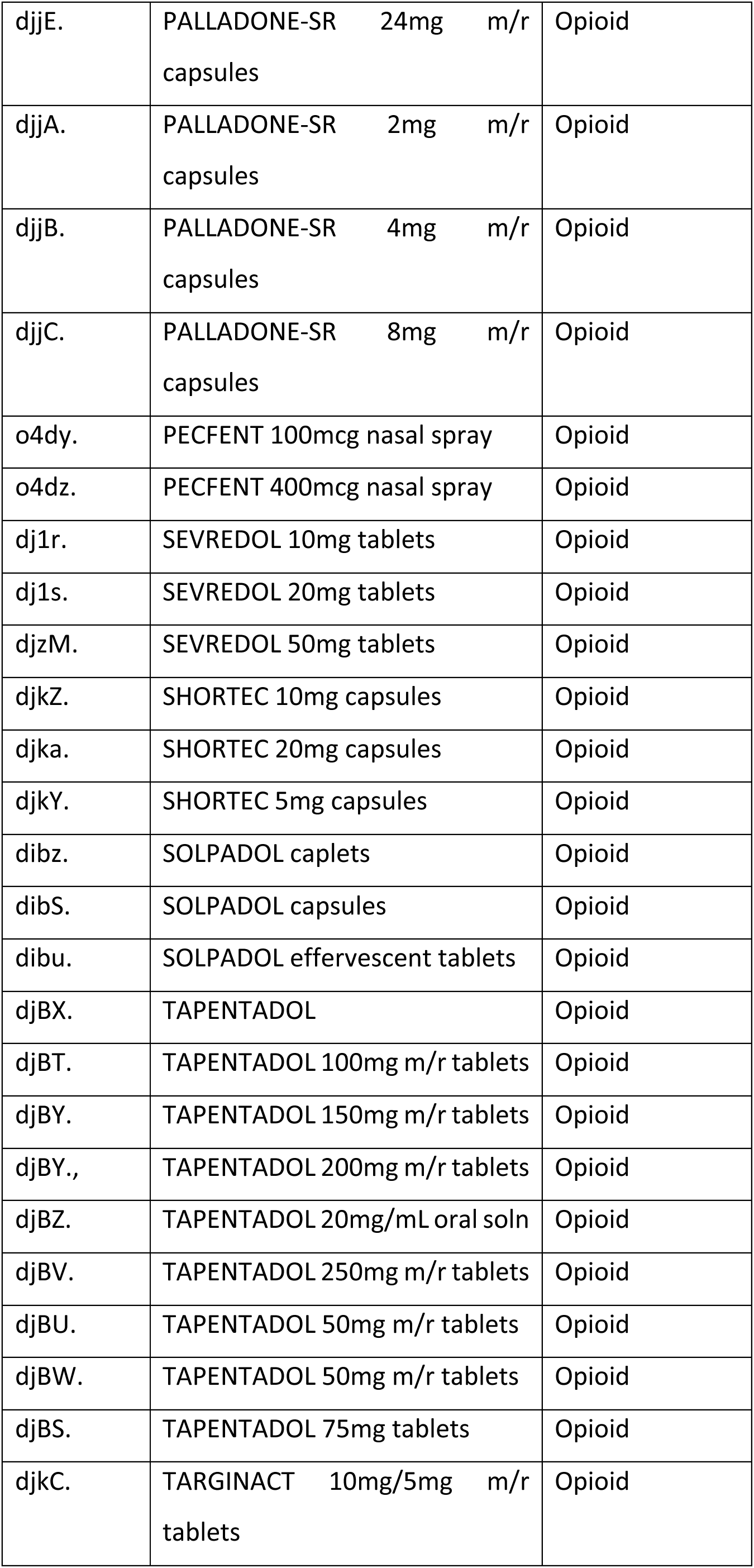
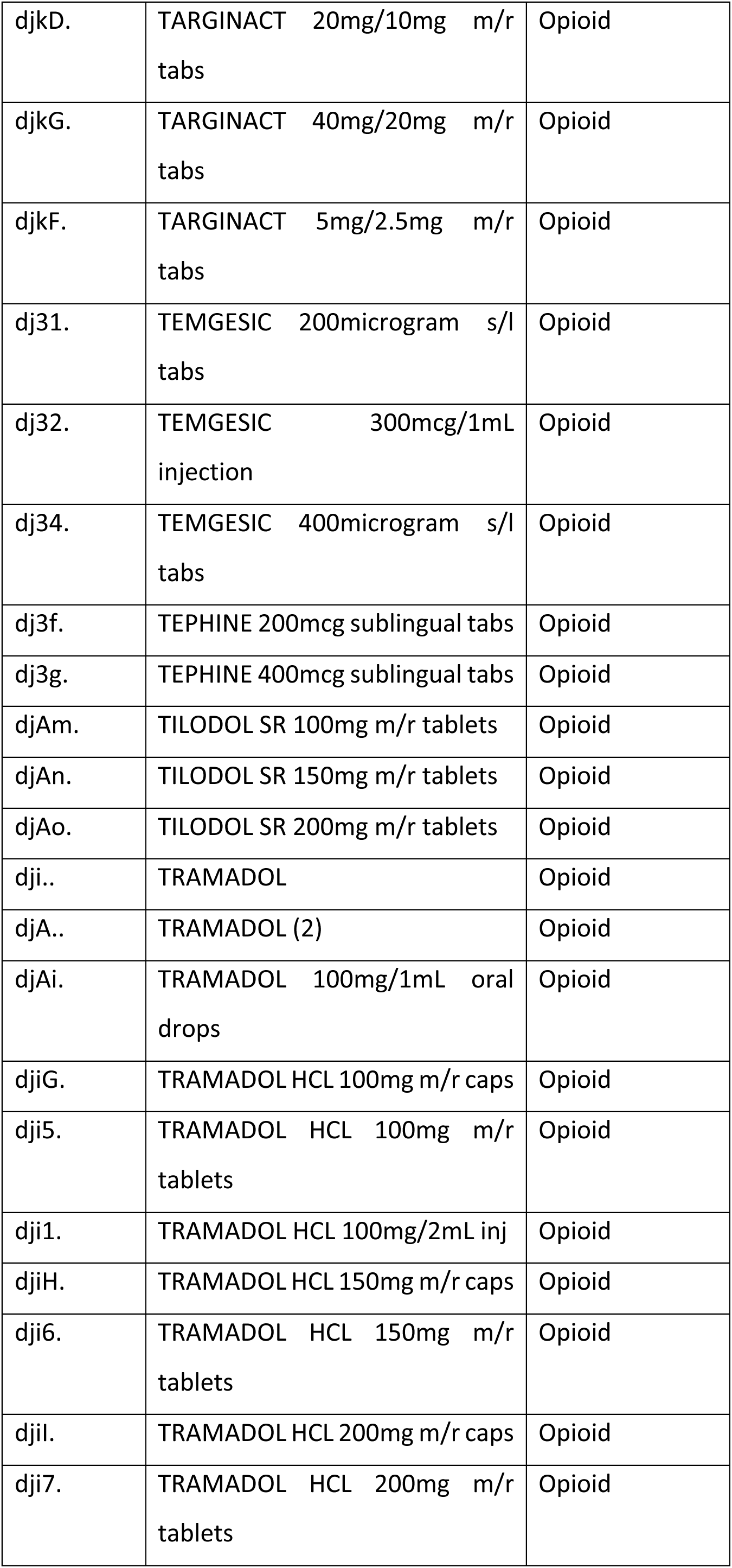
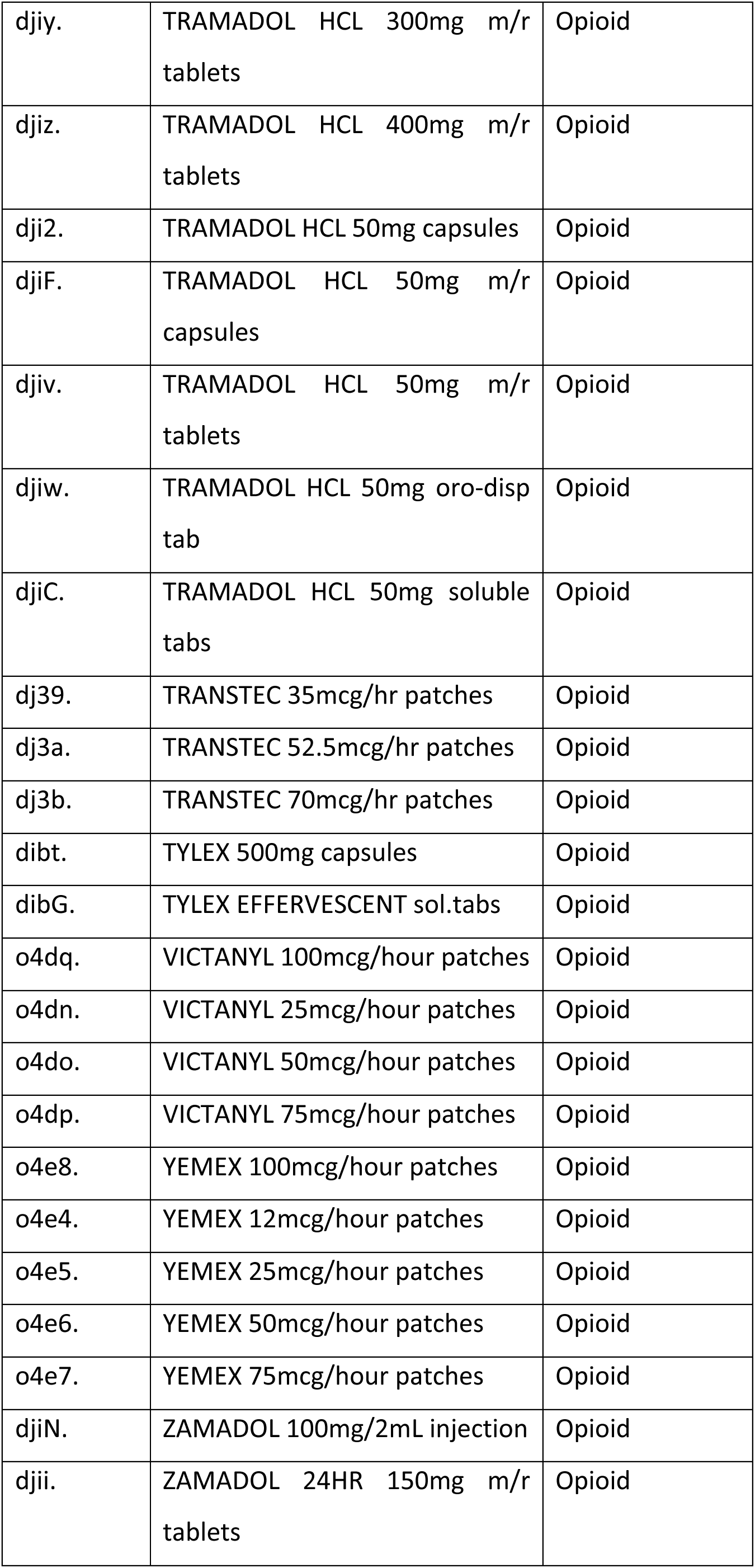
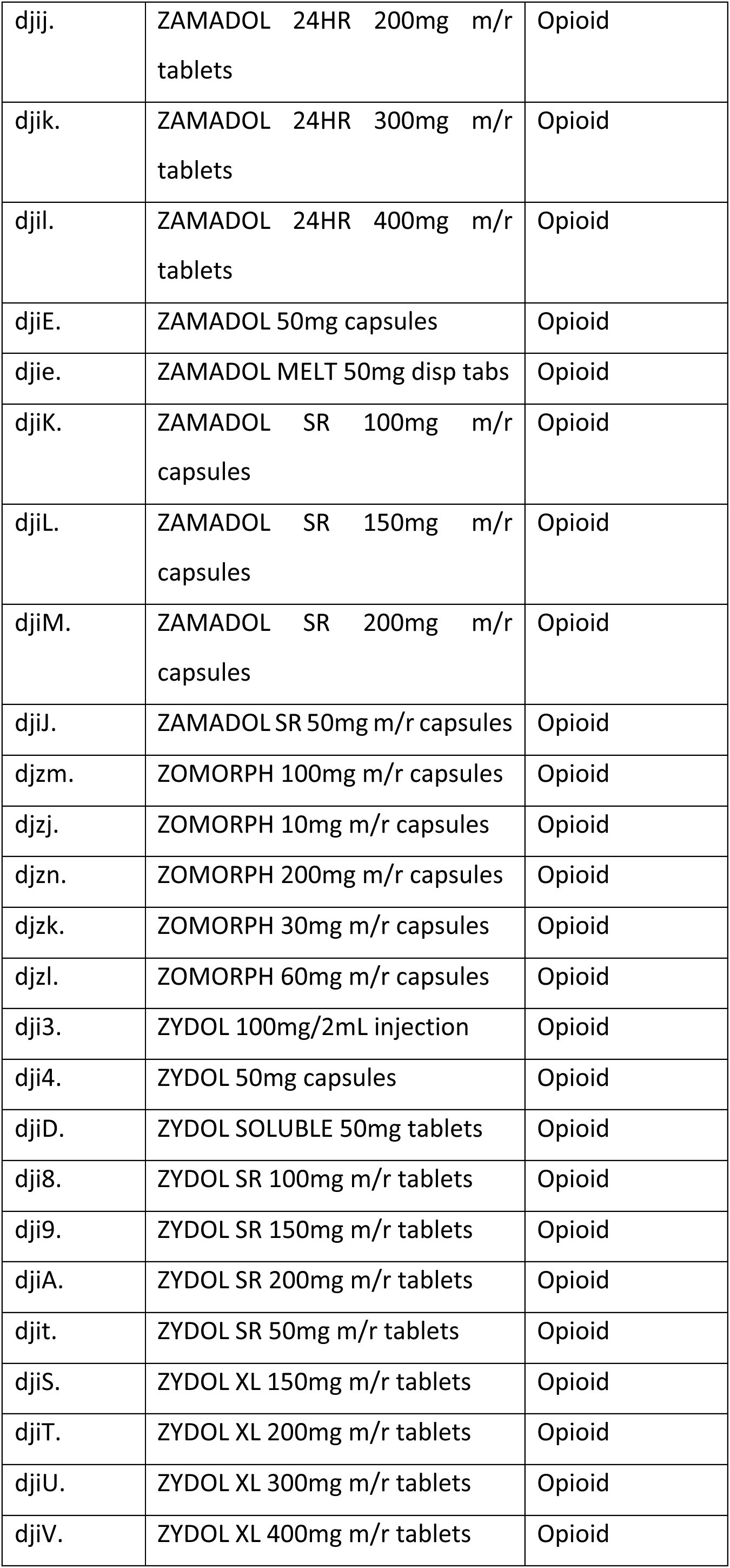

## Appendix 4 Graph and model coefficients for yearly change in cohort counts

**Figure 6.**
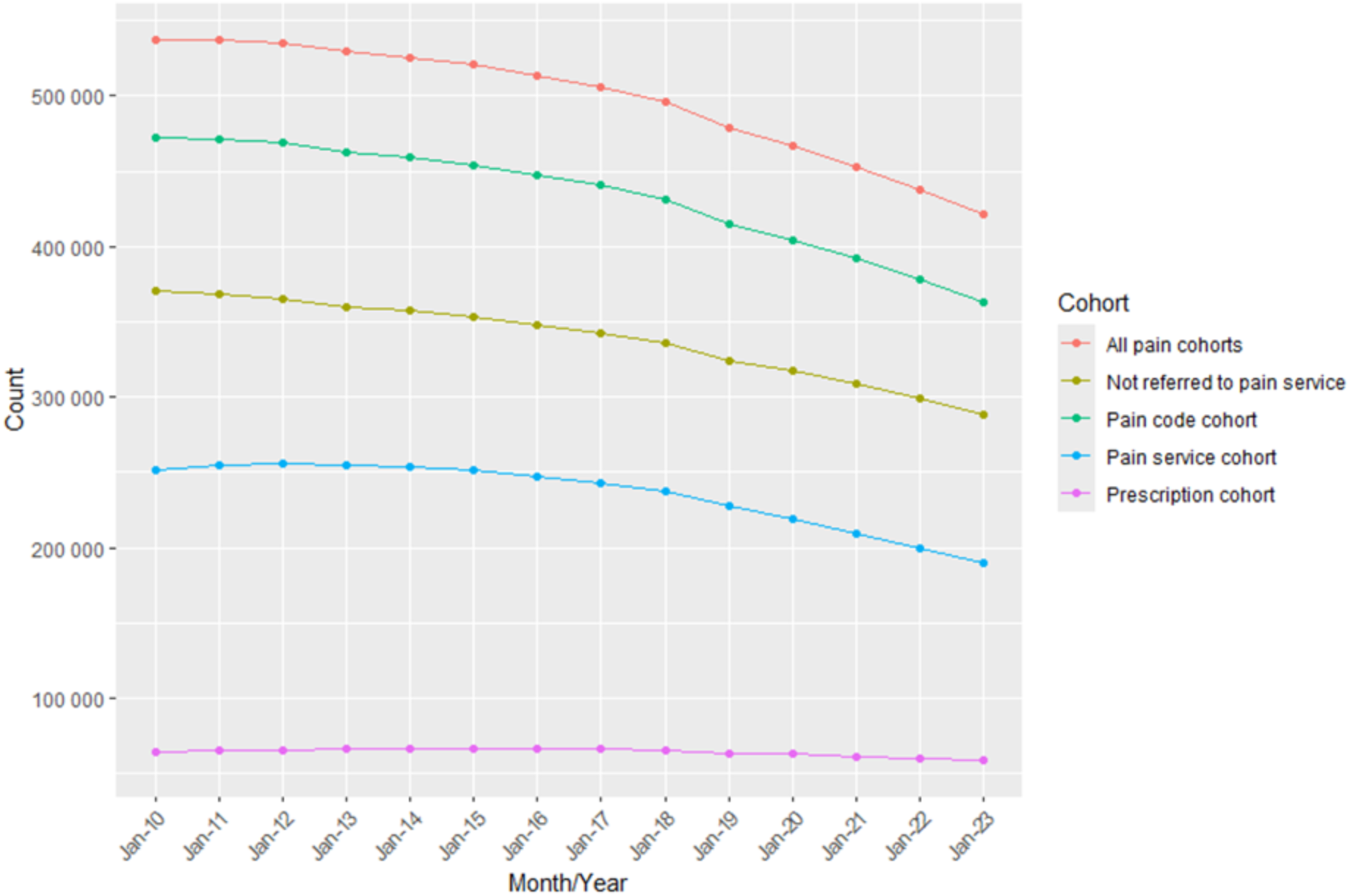
Graph of yearly cohort counts.

## Appendix 5 Coefficients for monthly trend in pain cohorts

1. All pain

RR: 0.9999328

CI: 0.9999254 to 0.9999401

p value: 4.267228e-73

Percentage change: -0.00672166

2. Diagnosis

RR: 0.9999315

CI: 0.9999260 to 0.9999370

p value: 1.814543e-133

Percentage change: -0.006850044

3. Prescription

RR: 0.9999238

CI: 0.9999106 to 0.9999369

p value: 7.454641e-31

Percentage change: -0.007624323

4. Pain service

RR: 0.9999622

CI: 0.9999541 to 0.99997019

p value: 1.061125e-20

Percentage change: -0.003783013

5. Non pain service RR: 0.9999284

CI: 0.9999211 to 0.9999357

p value: 1.196295e-83

Percentage change: -0.00715806

## Appendix 6 Monthly healthcare utilisation counts for each cohort

**Table 6.** Healthcare utilisation for comparator cohort.

| Month | GP events | Admissions | Emergency Admissions | Emergency attendances | Outpatient attendances |
| --- | --- | --- | --- | --- | --- |
| 2010.01 | 1,722,287 | 36,618 | 14,325 | 34,310 | 153,801 |
| 2011.01 | 1,799,707 | 37,633 | 15,063 | 39,491 | 164,682 |
| 2012.01 | 1,911,077 | 40,383 | 15,499 | 43,756 | 178,419 |
| 2013.01 | 1,995,663 | 39,237 | 14,803 | 42,898 | 174,548 |
| 2014.01 | 2,026,435 | 40,606 | 15,358 | 43,151 | 175,988 |
| 2015.01 | 1,979,787 | 40,570 | 15,655 | 42,076 | 173,608 |
| 2016.01 | 2,009,148 | 42,693 | 16,448 | 47,307 | 175,550 |
| 2017.01 | 2,171,240 | 44,047 | 17,072 | 46,957 | 193,577 |
| 2018.01 | 2,310,648 | 46,039 | 17,998 | 48,702 | 196,976 |
| 2019.01 | 2,350,501 | 48,060 | 19,033 | 50,371 | 201,085 |
| 2020.01 | 2,426,863 | 48,485 | 18,148 | 49,728 | 206,459 |
| 2021.01 | 2,381,711 | 31,445 | 12,525 | 32,142 | 144,951 |
| 2022.01 | 2,677,656 | 38,424 | 15,179 | 46,301 | 196,704 |
| 2023.01 | 2,663,791 | 44,281 | 17,099 | 48,730 | 231,751 |

**Table 7.** Healthcare utilisation for combined pain cohort.

| Month | GP events | Admissions | Emergency Admissions | Emergency attendances | Outpatient attendances |
| --- | --- | --- | --- | --- | --- |
| 2010.01 | 472,422 | 9,308 | 3,363 | 5,332 | 46,726 |
| 2011.01 | 610,828 | 13,063 | 4,818 | 7,520 | 62,017 |
| 2012.01 | 719,535 | 15,937 | 5,595 | 9,246 | 73,263 |
| 2013.01 | 793,875 | 16,262 | 5,806 | 10,243 | 77,389 |
| 2014.01 | 887,833 | 18,925 | 6,840 | 11,157 | 83,504 |
| 2015.01 | 896,273 | 18,756 | 7,051 | 11,608 | 84,767 |
| 2016.01 | 918,621 | 20,242 | 7,403 | 13,229 | 85,922 |
| 2017.01 | 1,014,046 | 21,425 | 8,046 | 13,445 | 96,028 |
| 2018.01 | 1,082,446 | 22,926 | 8,609 | 14,367 | 99,310 |
| 2019.01 | 1,107,413 | 23,694 | 8,909 | 14,700 | 103,635 |
| 2020.01 | 1,131,887 | 23,734 | 8,659 | 15,138 | 105,333 |
| 2021.01 | 1,079,355 | 15,515 | 6,679 | 11,321 | 67,118 |
| 2022.01 | 1,052,425 | 18,298 | 7,181 | 13,358 | 91,562 |
| 2023.01 | 1,126,039 | 20,792 | 7,846 | 13,985 | 107,308 |

**Table 8.** Healthcare utilisation for diagnosis cohort.

| Month | GP events | Admissions | Emergency Admissions | Emergency attendances | Outpatient attendances |
| --- | --- | --- | --- | --- | --- |
| 2010.01 | 409,654 | 8,207 | 2,979 | 4,718 | 41,061 |
| 2011.01 | 477,345 | 9,656 | 3,495 | 5,737 | 48,419 |
| 2012.01 | 539,500 | 11,330 | 3,850 | 6,671 | 55,025 |
| 2013.01 | 577,037 | 11,090 | 3,846 | 7,114 | 56,516 |
| 2014.01 | 636,137 | 12,879 | 4,517 | 7,640 | 59,856 |
| 2015.01 | 632,565 | 12,538 | 4,612 | 7,693 | 60,156 |
| 2016.01 | 640,189 | 13,502 | 4,811 | 8,799 | 60,446 |
| 2017.01 | 699,869 | 14,299 | 5,226 | 8,868 | 66,974 |
| 2018.01 | 740,372 | 15,261 | 5,549 | 9,373 | 68,660 |
| 2019.01 | 754,919 | 15,715 | 5,662 | 9,500 | 71,362 |
| 2020.01 | 769,524 | 15,899 | 5,604 | 9,901 | 72,690 |
| 2021.01 | 729,255 | 9,917 | 4,256 | 7,074 | 44,671 |
| 2022.01 | 701,632 | 12,007 | 4,658 | 8,541 | 61,685 |
| 2023.01 | 753,991 | 13,800 | 5,093 | 8,892 | 72,450 |

**Table 9.** Healthcare utilisation for prescription cohort.

| Month | GP events | Admissions | Emergency Admissions | Emergency attendances | Outpatient attendances |
| --- | --- | --- | --- | --- | --- |
| 2010.01 | 117,151 | 1,406 | 475 | 757 | 7,214 |
| 2011.01 | 167,011 | 3,497 | 1,247 | 1,850 | 16,594 |
| 2012.01 | 183,107 | 3,946 | 1,402 | 2,175 | 18,119 |
| 2013.01 | 191,116 | 3,780 | 1,291 | 2,230 | 18,492 |
| 2014.01 | 202,784 | 4,328 | 1,558 | 2,408 | 18,808 |
| 2015.01 | 191,078 | 4,097 | 1,427 | 2,324 | 18,006 |
| 2016.01 | 187,490 | 4,189 | 1,491 | 2,614 | 17,508 |
| 2017.01 | 193,125 | 4,046 | 1,436 | 2,413 | 18,395 |
| 2018.01 | 192,271 | 4,130 | 1,543 | 2,550 | 17,689 |
| 2019.01 | 182,921 | 4,037 | 1,507 | 2,341 | 17,328 |
| 2020.01 | 166,186 | 3,549 | 1,331 | 2,198 | 58,223 |
| 2021.01 | 146,523 | 2,325 | 948 | 1,649 | 9,902 |
| 2022.01 | 127,681 | 2,574 | 994 | 1,706 | 11,798 |
| 2023.01 | 101,939 | 2,206 | 875 | 1,384 | 10,364 |

**Table 10.** Healthcare utilisation for pain service cohort.

| Month | GP events | Admissions | Emergency Admissions | Emergency attendances | Outpatient attendances |
| --- | --- | --- | --- | --- | --- |
| 2010.01 | 45,479 | 906 | 294 | 612 | 5,081 |
| 2011.01 | 72,897 | 1,740 | 589 | 991 | 8,745 |
| 2012.01 | 95,997 | 2,310 | 758 | 1,299 | 11,631 |
| 2013.01 | 115,465 | 2,526 | 872 | 1,712 | 13,386 |
| 2014.01 | 133,257 | 3,072 | 1,042 | 1,875 | 14,617 |
| 2015.01 | 138,189 | 3,009 | 1,057 | 1,890 | 15,307 |
| 2016.01 | 145,259 | 3,391 | 1,221 | 2,284 | 15,684 |
| 2017.01 | 162,859 | 3,588 | 1,346 | 2,388 | 17,326 |
| 2018.01 | 176,778 | 3,819 | 1,383 | 2,448 | 18,640 |
| 2019.01 | 182,288 | 4,098 | 1,580 | 2,691 | 19,608 |
| 2020.01 | 186,075 | 3,815 | 1,436 | 2,678 | 19,387 |
| 2021.01 | 172,597 | 2,476 | 1,129 | 2,119 | 12,014 |
| 2022.01 | 175,037 | 2,981 | 1,242 | 2,448 | 16,917 |
| 2023.01 | 184,550 | 3,463 | 1,320 | 2,410 | 19,296 |

**Table 11.** Healthcare utilisation for non-pain service cohort.

| Month | GP events | Admissions | Emergency Admissions | Emergency attendances | Outpatient attendances |
| --- | --- | --- | --- | --- | --- |
| 2010.01 | 426,943 | 8,402 | 3,069 | 4,720 | 41,645 |
| 2011.01 | 537,931 | 11,323 | 4,229 | 6,529 | 53,272 |
| 2012.01 | 623,538 | 13,627 | 4,837 | 7,947 | 61,632 |
| 2013.01 | 678,410 | 13,736 | 4,934 | 8,531 | 64,003 |
| 2014.01 | 754,576 | 15,853 | 5,798 | 9,282 | 68,887 |
| 2015.01 | 758,084 | 15,747 | 5,994 | 9,718 | 69,460 |
| 2016.01 | 773,362 | 16,851 | 6,182 | 10,945 | 70,238 |
| 2017.01 | 851,187 | 17,837 | 6,700 | 11,057 | 78,702 |
| 2018.01 | 905,668 | 19,107 | 7,226 | 11,919 | 80,670 |
| 2019.01 | 925,125 | 19,596 | 7,329 | 12,009 | 84,027 |
| 2020.01 | 945,812 | 19,919 | 7,223 | 12,460 | 85,946 |
| 2021.01 | 906,758 | 13,039 | 5,550 | 9,202 | 55,104 |
| 2022.01 | 877,388 | 15,317 | 5,939 | 10,910 | 74,645 |
| 2023.01 | 941,489 | 17,329 | 6,526 | 11,575 | 88,012 |

## Appendix 7 Coefficients for monthly change in healthcare utilisation for pain cohort

1. GP events

RR: 1.000052

CI: 1.0000501 to 1.0000531

p value: 0

Percentage change: 0.005160515

2. PEDW

RR: 1.000012

CI: 1.00000973 to 1.0000148

p value: 4.190824e-21

Percentage change: 0.001228515

3. PEDW emergency

RR: 0.9999997

CI: 0.9999955 to 1.0000039

p value: 8.903084e-01

Percentage change: -2.981367e-05

4. EDDS

RR: 1.000006

CI: 1.0000026 to 1.0000091

p value: 4.136162e-04

Percentage change: 0.0005864294

5. OPDW

RR: 0.9999824

CI: 0.9999812 to 0.9999836

p value: 4.686760e-183

Percentage change: -0.001761389

## Appendix 8 Regression coefficients for the difference in healthcare utilisation between pain and comparator cohort

1. GP events

RR: 1.625988

CI: 1.6246580 to 1.6273187

p value: 0.0000000

Percentage difference: 62.59882

2. PEDW

RR: 0.9719469

CI: 0.9705392 to 0.9733565

p value: 0.000000e+00

Percentage difference: -2.805308

3. PEDW emergency admissions RR: 1.008847

CI: 1.006414 to 1.011286

p value: 8.691537e-13

Percentage difference: 0.8847125

4. EDDS

RR: 1.019355

CI: 1.017587 to 1.021126

p value: 6.703982e-104

Percentage difference: 1.935532

5. OPDW

RR: 1.027361

CI: 1.026648 1.028075

p value: 0.000000e+00

Percentage difference: 2.736142

## Appendix 9 Regression coefficients for the difference in healthcare utilisation between pain service and non-pain service cohort

1. GP events

RR: 1.327872

p value: 0

Percentage difference: 32.78718

95% CI: 1.325428 to 1.330321

2. PEDW admissions

RR: 0.955908

p value: 0

Percentage difference: -4.409203

95% CI: 0.95297949 to 0.9588435

3. PEDW emergency admissions RR: 1.010012

p value: 1.166564e-04

Percentage difference: 1.001153

95% CI: 1.004904 to 1.015140

4. EDDS emergency attendances RR: 1.02017

p value: 1.437087e-26

Percentage difference: 2.017157

95% CI: 1.016434 to 1.023920

5. OPDW outpatient attendances RR: 1.023332

p value: 1.482671e-230

Percentage difference: 2.333179

95% CI: 1.021906 to 1.024759

## Appendix 10 Decomposition plots

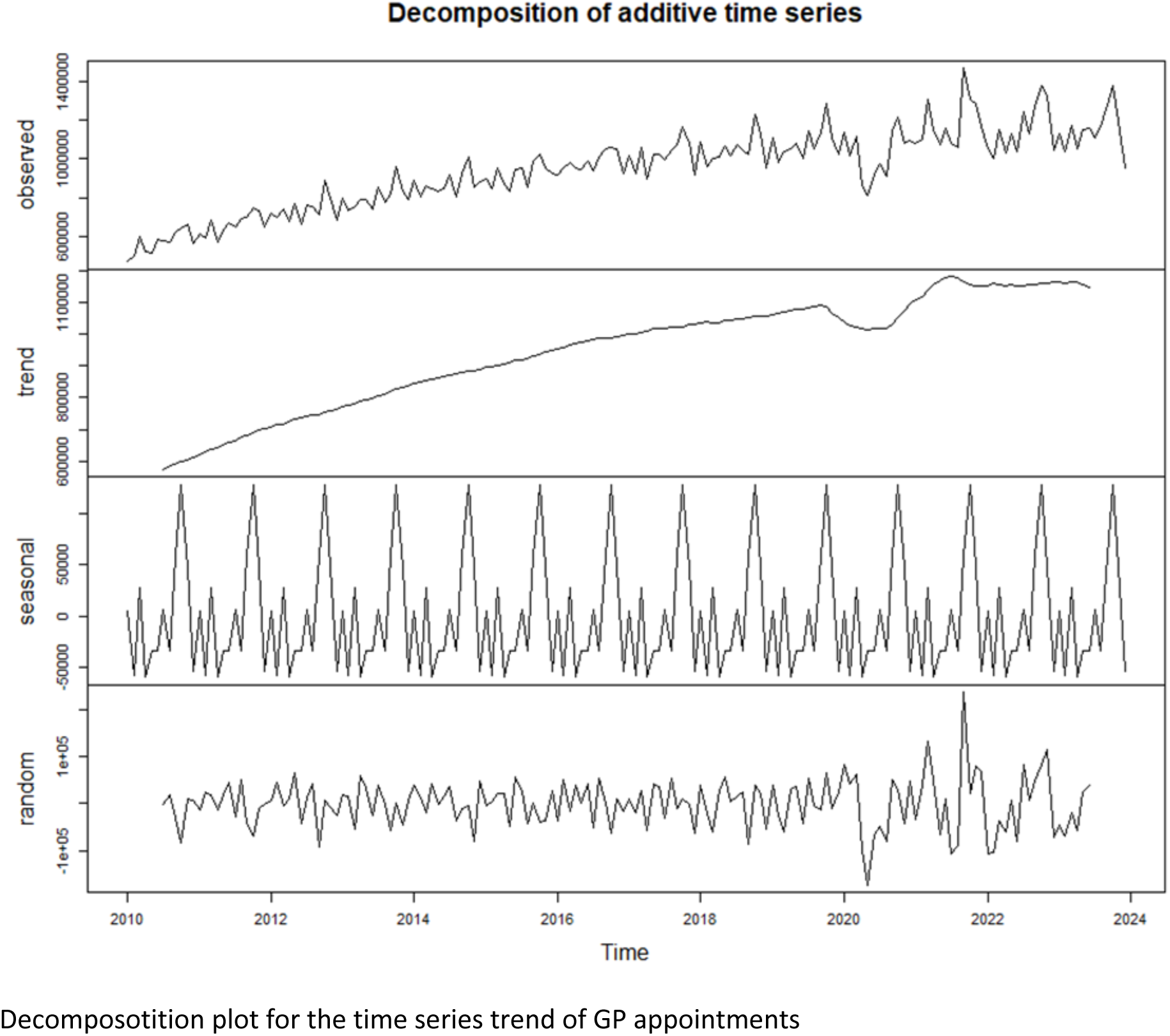
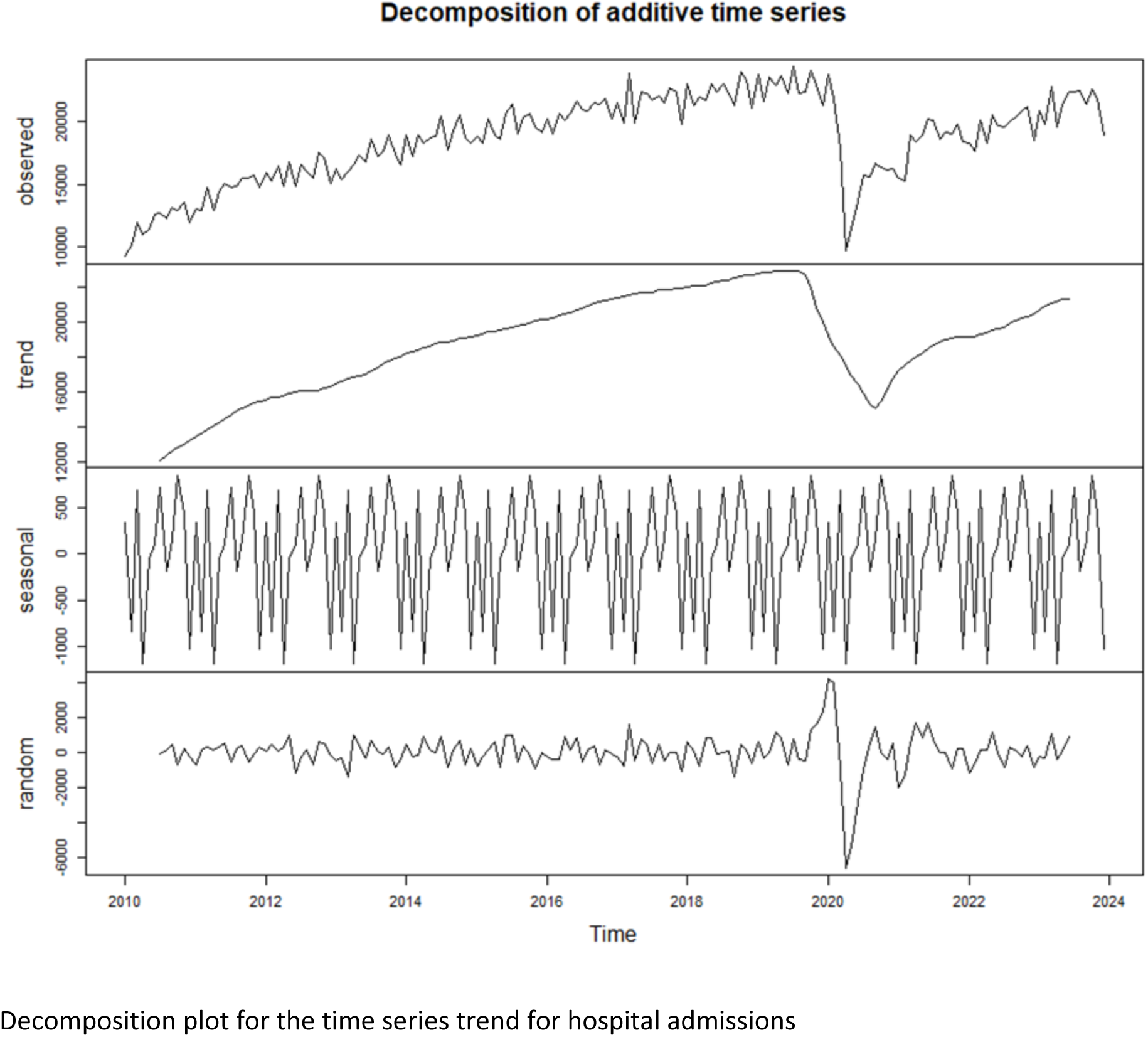
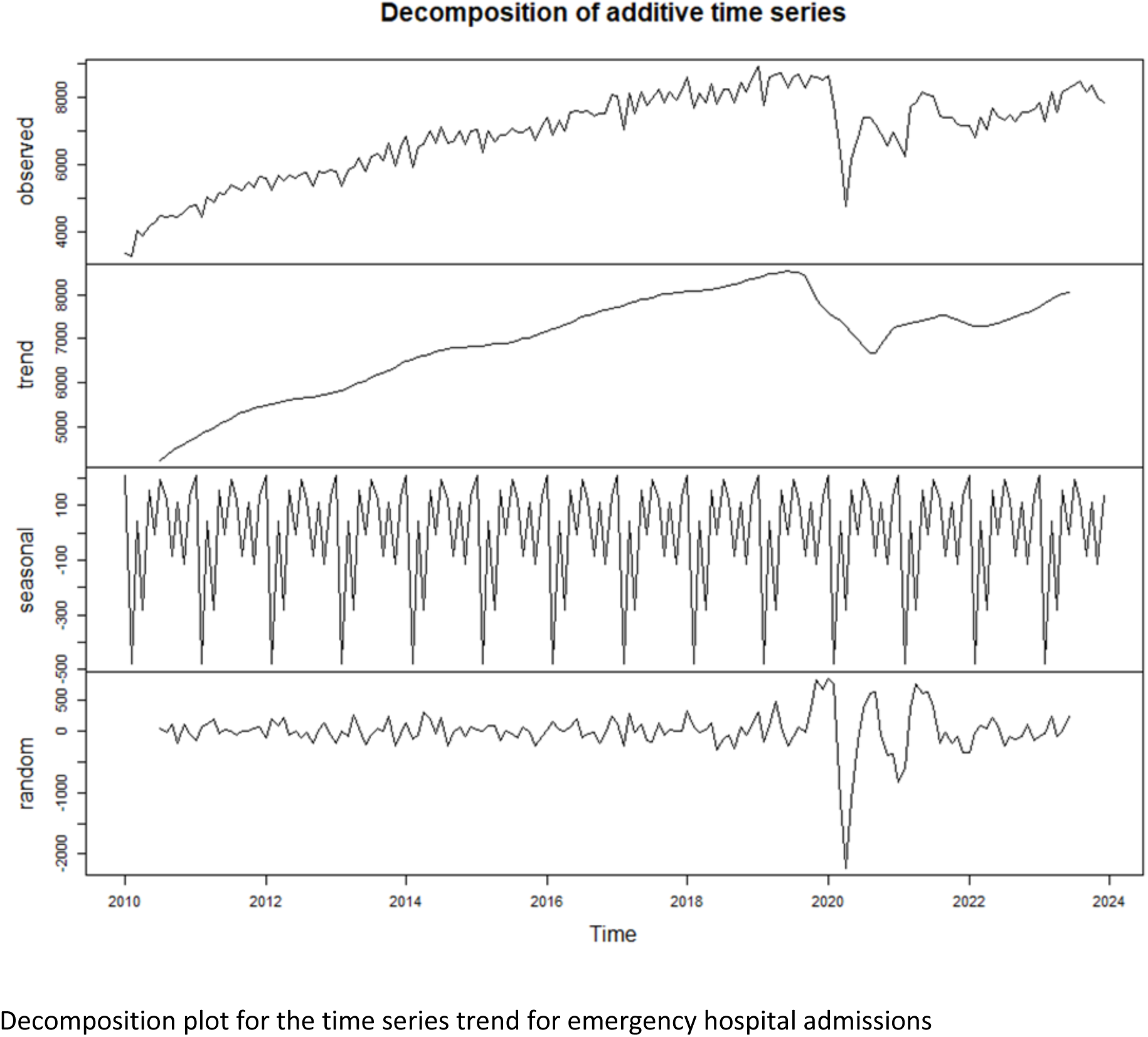
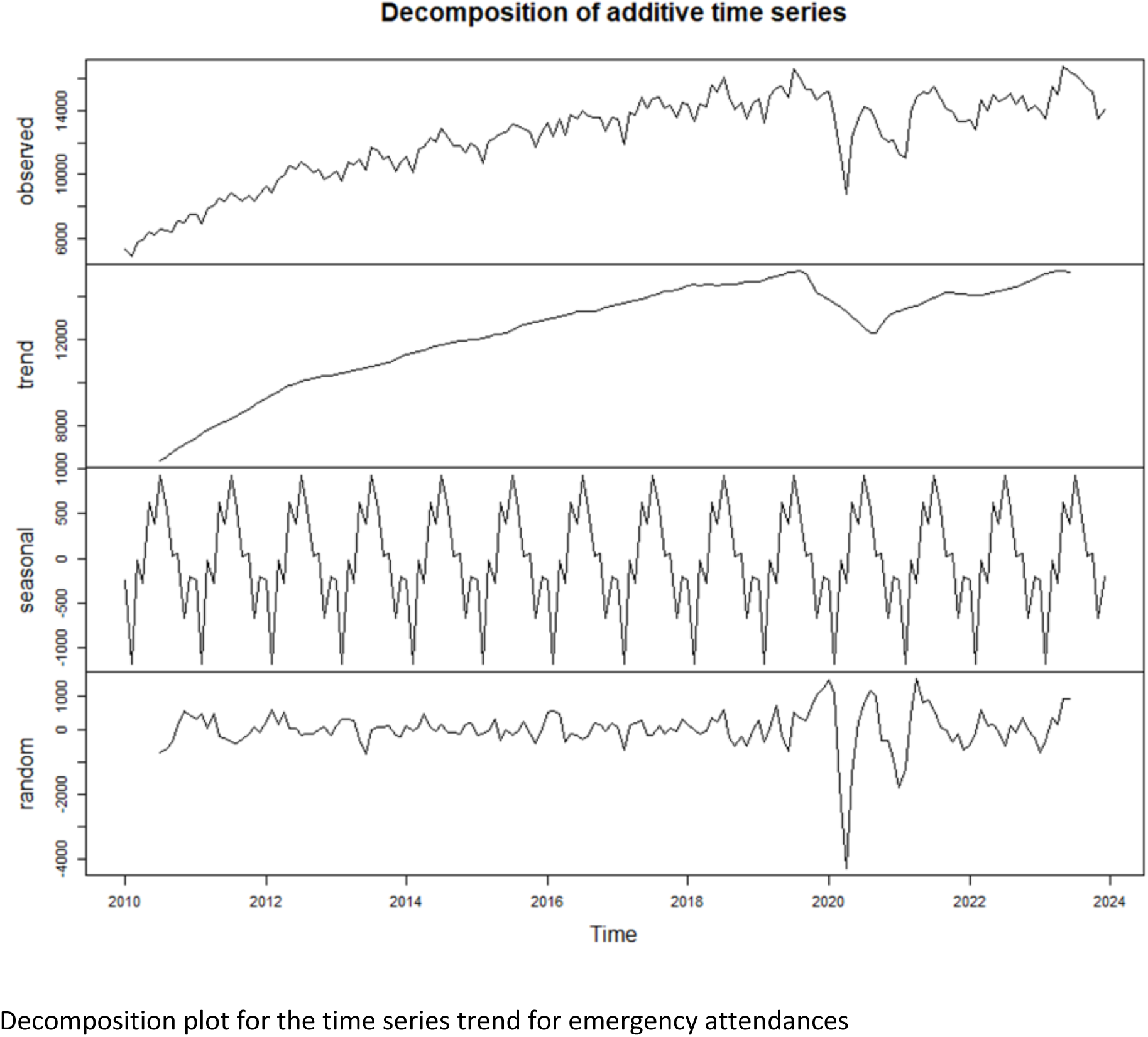
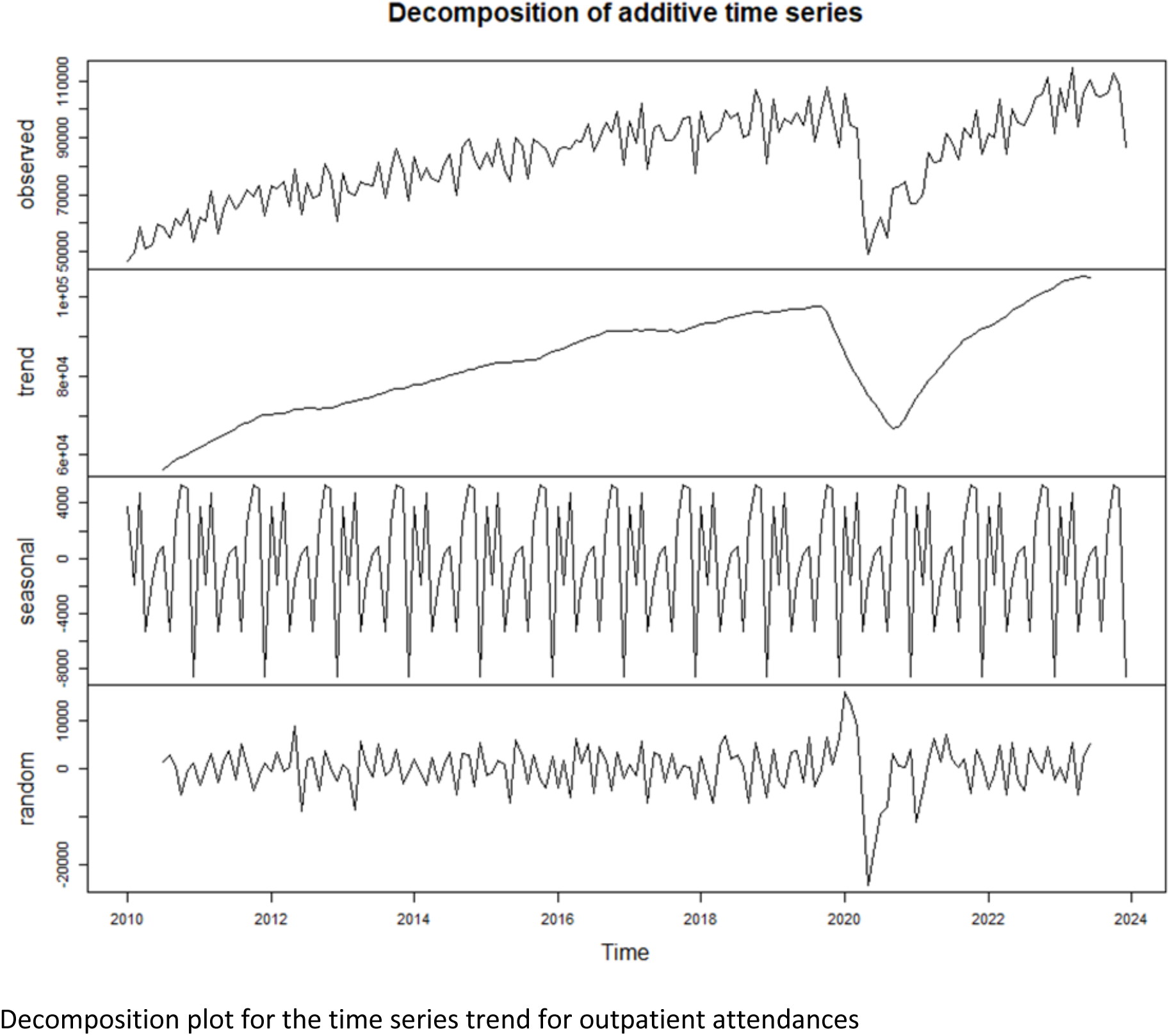

